# From Bone-Centric to Kidney-Centric: Environment-Dependent Shift of Spaceflight Renal Stone Pathways

**DOI:** 10.64898/2026.08.27.26360881

**Authors:** Jian Shi, Qiutao Gu, Jiawei Pan, Anqi Yang, Yuan Zhang, Linglong Jiang, Yangyang Sun, Jundong Zhu, Min Fan

## Abstract

Human deep-space missions face bone–kidney risks that cannot be extrapolated from six-month ISS data. We built a 12-state Ca-bone-urine-stone mechanistic ODE model and jointly calibrated its 11 physiological parameters on eight ISS targets by Bayesian identification (M₀ base = 19-D; M₁ extension adds a GCR–bone coupling term for parsimony testing only), then propagated the M₀ posterior to four environments (ISS, Lunar subsurface, Lunar surface, Mars). Lumbar-lower BMD loss increases with mission duration and partial-gravity unloading (ISS 180 d −4.83% → Mars 730 d −12.15%; 2³ factorial: duration 82.9%, gravity 12.5%, GCR main effect ≈ 0), whereas stone rate follows the opposite gradient (ISS 16.1 vs Mars 13.1 per 1000 person-years), reflecting weakened partial-gravity bone resorption alongside residual urinary chemistry changes. The dominant pathway thus shifts from bone-centric on the ISS to kidney-centric on Mars, where residual urinary-chemistry changes—not bone resorption—drive stone risk. The direct GCR–bone coupling term is unidentifiable at current ISS doses (ΔWAIC = +0.0076 ± 0.126 SE), so M₀ is retained as the main inference model. Bisphosphonates provide ≥84% BMD protection but leave a urinary-chemistry residual, so bisphosphonate monotherapy would underestimate Mars stone risk; potassium–magnesium–citrate combinations (RRR_RSS 51%) should therefore be added to deep-space countermeasures. A Lunar-surface 365-day mission is the earliest environment on the NASA roadmap to cross a composite RED threshold. That profile differs from the regolith-shielded 180-day case in both cumulative GCR (∼69×) and duration (2×), so a shielding-specific effect cannot be isolated here; forcing the GCR coupling terms to zero leaves all four composite tiers unchanged (0/4, Supp §S24), and the shielded 180-day profile is YELLOW rather than GREEN. Independent hold-out validation (Culliton 2025 60-day HDT-bedrest RCT, n=8 control arm of n=24 total) supports the M₀ posterior predictive distribution on the lumbar-BMD sub-scope.

### Lay Summary

NASA classifies bone loss and renal stone formation among the highest-severity medical risks of long-duration spaceflight, yet the quantitative evidence base is derived almost entirely from six-month ISS missions. How to extrapolate these risks to Artemis lunar bases and 730-day Mars round-trip missions remains a critical blind spot in space medicine.

We integrated bone metabolism, calcium cycling, urine chemistry, and stone formation into a mechanistic mathematical model. We jointly calibrated it against eight independent ISS observational targets and propagated the posterior to four future environments—ISS, lunar subsurface, lunar surface, and Mars—under six prevention strategies.

**Most surprising**: monthly bone loss is slower under partial gravity than on the ISS, yet cumulative loss over long missions is greater—a 730-day Mars mission accrues approximately 2.5-fold the lumbar BMD loss of a six-month ISS mission (12.1% vs 4.8%), whereas the per-person-year stone rate is slightly lower (13.1 vs 16.1 per 1000 person-years).

**Most actionable**: **a 365-day lunar-surface mission (anticipated for Artemis long-duration deployment in the 2030s) is the earliest environment on the NASA roadmap to trigger a composite bone–kidney RED risk tier**, preceding Mars. This RED status is jointly determined by the 365-day mission duration, 0.166 g partial gravity, and unshielded 1.7 mSv/day cosmic radiation; the subsurface and surface profiles differ simultaneously in GCR and mission duration, so an independent shielding effect cannot be isolated from this observational contrast (forcing the GCR coupling terms to zero flips 0 of 4 environment tiers). An in-model 2×2 factorial (fixed 0.166 g, duration × GCR dose rate, Supp §S27) quantitatively separates this confounding: the dose-rate main effect is ≈ 0, and the lunar-surface RED is dominated by the 365-day duration. At least 1 m of regolith reduces GCR to ∼3%, but the corresponding subsurface 180-day profile is YELLOW—not all endpoints meet acceptable thresholds. On Mars, bisphosphonate monotherapy protects ≥84% of bone mass but leaves a renal-chemistry residual, requiring combination with potassium–magnesium–citrate agents and aggressive hydration.

**Limitations**: direct measurements of human bone metabolism under partial gravity do not yet exist, and Mars prediction uncertainty is approximately ±20–30%. Future Artemis lunar-surface BMD data are the key window for narrowing this uncertainty.

## 1. Introduction

The NASA Human Research Program Evidence Report (HRP 2016) [22] classifies bone loss and renal stone formation as Likelihood 5–Consequence 4 (L5–C4) flight-medicine risks. Pietrzyk et al. (2007, Aviat. Space Environ. Med.) [2] documented significantly elevated post-flight renal stone risk in a NASA astronaut cohort. Goodenow-Messman et al. (2022) [9] applied population balance equation (PBE) probabilistic analysis to 1,517 24-h urine samples, using the Porter–Rice aviator cohort (military + commercial aviator stone rate 4.40 per 1000 person-years) as the prior and the Sibonga & Pietrzyk observation (7 stones during the first post-flight year / 358 astronaut-years) as the updating data, yielding a Bayesian posterior post-flight incidence rate of approximately 17.3 per 1000 astronaut-years (95% CI [8.33, 28.80] per 1000 person-years, i.e. 0.0173/py [0.0083, 0.0288]). This rate is approximately twice the pre-flight characteristic incidence rate (IR) of 0.0085 per person-year and provides the best currently quantifiable post-flight reference baseline.

Smith et al. (2015, Bone) [3] provided a systematic synthesis of bone metabolism and renal stone risk during long-duration ISS missions. Whitson et al. (1997, J. Urol.) reported that Space Shuttle short-duration missions shifted urine chemistry in a stone-promoting direction: increased urinary calcium, mild reductions in urine volume and citrate, and decreased urine pH [4]. LeBlanc et al. (2000, 2007) compiled ISS/Mir long-duration BMD data, documenting approximately 6% loss of lower lumbar spine BMD after six months [5,17].

More recently, Sibonga et al. (2019) reported partial protection from ARED (Advanced Resistive Exercise Device) resistance training against ISS crew bone loss (mechanical protection of cortical bone and hip, without suppressing bone resorption biomarkers) [6]. Watanabe et al. (2004) observed zero stone formation events among seven subjects receiving pamidronate in a bed-rest RCT (versus two of nine controls and four of nine resistance-training subjects) [7]. LeBlanc et al. (2013) demonstrated that 70 mg/week oral alendronate combined with ARED prevented BMD decline in the spine, hip, and pelvis and suppressed bone resorption during ISS missions of mean duration 5.5 months [8]; our posterior estimate of this combination’s BMD-protection RRR (relative risk reduction) is approximately 95% (95% CrI: see Table 4).

However, all existing quantitative data originate from ISS microgravity (0 g) with low galactic cosmic radiation (GCR) exposure (∼0.4 mSv d⁻¹) over six-month missions. With the Artemis lunar programme entering operational surface-mission planning and future 730-day Mars round-trip missions (NASA Mars Reference Mission) under active study, astronauts will encounter a fundamentally new environmental matrix. Lunar subsurface or regolith-shielded habitats experience 0.166 g plus 0.05 mSv d⁻¹ (≥1 m regolith attenuating GCR to ∼3% of lunar surface levels [31]). Lunar surface habitats experience 0.166 g plus 1.7 mSv d⁻¹ (no atmosphere, direct GCR exposure). Mars missions experience 0.38 g plus 1.84 mSv d⁻¹ over 730 days. Three critical gaps exist in the current literature: (1) the absence of an integrated cross-environment mechanistic model—Sibonga 2019 [6] and Goodenow-Messman 2022 [9] performed single-endpoint regressions on BMD or urine chemistry confined to ISS, precluding extrapolation to non-microgravity environments; (2) the lack of quantitative predictions for countermeasure efficacy (RRR) across varying gravity–GCR environments; and (3) no publicly reported Bayesian joint calibration across the multi-source spaceflight dataset (Pietrzyk stone rates, Whitson urine chemistry, LeBlanc BMD, and Smith 2014 post-flight recovery).

To address these gaps, we constructed a 12-state calcium–bone–urine–stone mechanistic ODE model. Module A represents bone resorption; Module B, bone formation; Module C, calcium homeostasis; Module D, urine chemistry; Module E, RSS_CaOx supersaturation index; and Module F, stone nucleation and growth. We performed joint Bayesian calibration against eight ISS data targets (emcee affine-invariant MCMC, 64 walkers × 60,000 steps, 19 dimensions plus a 1-dimensional GCR–BMD channel extension). We then propagated the posterior to four environments and quantified BMD loss rate, RSS_CaOx, and stone formation rate with 95% CrI stratified by a GREEN–YELLOW–RED decision tier. The core finding of this study is that the dominant pathway is Bone-centric on the ISS (bone resorption → urinary Ca → stone), whereas under Martian 0.38 g partial gravity a Kidney-centric re-weighting of relative pathway weights occurs—partial gravity partially rescues bone resorption (730-day-matched protection +7.44%), and residual stone risk is driven by microgravity-induced reductions in urine volume and citrate. This implies that bisphosphonate-only strategies may underestimate stone risk for Mars missions, and K–Mg–citrate agents warrant consideration for the countermeasure portfolio.

**Table 1.** Physical parameters of the four environments. Table 1 notes: **micro_g** = 1 − (g_environment / g_earth) represents the microgravity contribution relative to Earth (ISS full microgravity 1.000; Mars partial gravity 0.620; Lunar Surface and Lunar UG both 0.834). **GCR dose-rate sources**: [32] Cucinotta 2011 NASA TP “Space Radiation Cancer Risk Projections” (NTRS 20130001648), anchoring ISS at 0.4 mSv/d; [33] Zhang 2020 Science Advances, Chang’E-4 LND direct measurement of lunar surface GCR ∼1.4 mSv/d (this study takes the NASA HSRB DRM 1.7 mSv/d rounded estimate including SPE and neutron contributions); [34] Akisheva 2024 CEAS Space J / Dobynde 2024 Nat Astron, regolith ≥1 m shielding analysis (≥1 m attenuates GCR to ∼3% of surface levels, i.e. ∼0.05 mSv/d); [35] Slaba 2017 LSSR 12:1–15, LET HZE GCR fragmentation (online 2016, print 2017); [36] Zeitlin 2013 Science, Curiosity RAD Mars cruise direct measurement 1.8 mSv/d (rounded to 1.84); [37] Hassler 2014 Science, Curiosity RAD Mars surface ∼0.7 mSv/d (this study’s 1.84 is transit-dominated). **Total GCR** = daily dose rate × mission duration. **Mars cumulative dose ∼1.34 Sv approaches the NASA career limit of ∼1 Sv** (NCRP 132 [38] + NASA-STD-3001 Vol 1 Rev B). **Lunar UG and Lunar Surface share the same 0.166 g lunar gravity** (micro_g = 0.834), and their difference is not limited to GCR exposure: the dose rate differs 34× (regolith-shielded 0.05 vs surface-exposed 1.70 mSv/d), the mission duration also differs 2.03× (180 vs 365 d), and the cumulative dose therefore differs by about 69× (9 vs 620 mSv); this contrast cannot identify the independent effect of shielding (§3.5).

| Environment | micro_g (= 1 – g/g_earth) | GCR (mSv/d) | Mission duration (d) | Total GCR (mSv) | GCR source |
| --- | --- | --- | --- | --- | --- |
| ISS | 1.000 (0g, LEO microgravity) | 0.40 | 180 | 72 | [32] |
| Lunar UG (regolith-shielded subsurface habitat) | 0.834 (0.166g lunar surface gravity) | 0.05 | 180 | 9 | [33,34] |
| Lunar Surface | 0.834 (0.166g lunar surface gravity) | 1.70 | 365 | 620 | [33,35] |
| Mars (transit + surface) | 0.620 (0.38g Martian surface gravity) | 1.84 | 730 | 1343 | [36,37] |

## 2. Methods

### 2.1 Twelve-state ODE model (Modules A–F)

The model comprises six interconnected modules (complete ODE equations are provided in Supplementary §M1):

**Module A — Bone resorption**: a two-compartment bone resorption submodel in which the resorption plateau resorp_plateau (posterior median 113.06% [108.81, 117.16]) is approached with an exponential half-life resorp_t_half (median 11.04 d [7.14, 15.02]), calibrated against Smith/ Heer et al. (2012, JBMR) [21] ISS long-duration biomarker data (NTX/CTX plateau magnitudes consistent with the reported range of ∼110–115%).

**Module B — Bone formation**: a formation lag period form_lag, followed by post-flight rebound post_form_pct_mo (median 84.6%/month [42.1, 125.9]), calibrated against Smith et al. (2014) post-ISS bone marker recovery data [10].

**Module C — Calcium homeostasis**: a whole-body calcium pool with bone–urine–gut balance. The microgravity–calcium effect ca_microg_effect (median 0.501 [0.261, 0.742]) matches the Whitson 1997 +50% urinary Ca anchor. The GCR–calcium coupling term k_Ca_gcr (median 8.9 × 10⁻⁵, 95% CrI [−1.89 × 10⁻², +1.92 × 10⁻²], CV = 11502%) is prior-dominated, with no identifiable direct effect in the ISS data.

**Module D — Urine chemistry**: urinary calcium, volume V (vol_decrease_frac median −19.9% [−11.5%, −28.5%], matching the Whitson −20% anchor), citrate (cit_decrease_frac median −10.0% [−4.2%, −15.8%]), and pH (ph_decrease_frac median −3.34% [−1.58%, −5.01%], corresponding to a post-flight ΔpH ≈ −0.20).

**Module E — RSS_CaOx calculation**: a semi-empirical supersaturation index (Tiselius equivalent) RSS = f(Ca²⁺, ox²⁻, Mg²⁺, Ca²⁺ × citrate complex, pH), calibrated against Werness et al. (1985) EQUIL2 [11] and the Robertson et al. (2012, Arab J. Urol.) review formulation [19].

**Module F — Stone nucleation and growth**: a stone seed accumulation rate seed_accum_rate (LogNormal prior, median 0.114 [0.019, 0.643], CV = 105.5% reflecting the heavy tail of the Pietrzyk anchor) interacting with an RSS_crit threshold, with continued growth during the post-flight tail period (within the post_form_pct_mo window). Output: stone_rate (per person-year).

**Semantic note on post_form_pct_mo**: the Table 2 posterior median post_form_pct_mo = 84.60%/mo is the **monthly increase of bone-formation biomarkers relative to baseline** (serum markers such as osteocalcin, P1NP, and bone-specific alkaline phosphatase), not a DXA-BMD monthly recovery rate. This parameter corresponds to the post-flight “+84%/mo [39, 129] sustained for 3–5 months” biomarker increase reported in the Stavnichuk 2020 meta-analysis [23]. DXA-BMD post-flight recovery is substantially slower, typically requiring 6–12 months for partial recovery (Sibonga 2007 Bone [18] reports ∼9 months for 50% lumbar BMD recovery; Smith 2014 [10] reports partial recovery at 12 months). Module B implements a two-stage mapping (biomarker → BMD increment): the 84%/mo biomarker increase is multiplied by a marker-to-BMD conversion coefficient (implicit in the ODE, calibrated against the post-flight post_resorp_tau ∼59 d and the LeBlanc 2007 review [17] data).

**Table 2.** Posterior summary of the 11 core physical parameters of the M₀ model. Table 2 notes: Median = posterior median; 95% CrI = 2.5–97.5 equal-tailed quantiles; CV = 95% CrI width / median. The extremely large CV of k_Ca_gcr reflects that this parameter is prior-dominated, with no identifiable effect in the data. resorp_plateau_pct and post_form_pct_mo are percentages relative to the **serum bone-remodelling biomarker** baseline (NTX/CTX, P1NP/osteocalcin/ALP), not direct DXA-BMD readings (DXA monthly changes are ∼1%/mo, whereas biomarker changes are on the order of 10–100%/mo). See the semantic note in §2.1.

| Parameter | Physical meaning | Posterior median | 95% CrI | CV (%) |
| --- | --- | --- | --- | --- |
| resorp_plateau_pct | Bone resorption plateau (% baseline) | 113.06 | [108.81, 117.16] | 1.87 |
| resorp_t_half_d | Resorption rise half-life (d) | 11.04 | [7.14, 15.02] | 18.1 |
| bmd_loss_rate_pct_mo | BMD loss rate (%/mo) | -0.7989 | [-0.8703, -0.7246] | 4.56 |
| ca_microg_effect | Microgravity–urinary Ca gain (frac) | 0.5014 | [0.2606, 0.7417] | 24.0 |
| vol_decrease_frac | Microgravity–urine volume decrease (frac) | 0.1986 | [0.1147, 0.2852] | 21.0 |
| cit_decrease_frac | Microgravity–citrate decrease (frac) | 0.0997 | [0.0422, 0.1578] | 29.8 |
| k_Ca_gcr | GCR–Ca coupling (per mSv) | 8.865e-05 | [-1.891e-02, 1.922e-02] | 11502 |
| seed_accum_rate | Stone seed accumulation rate (1/d) | 0.1138 | [0.0190, 0.6427] | 105.5 |
| post_resorp_tau_d | Post-flight resorption decay $\tau$ (d) | 59.32 | [31.05, 88.98] | 24.9 |
| post_form_pct_mo | Post-flight formation rate (%/mo) | 84.60 | [42.08, 125.92] | 25.4 |
| ph_decrease_frac | Microgravity-pH decrease (frac) | 0.0334 | [0.0158, 0.0501] | 26.2 |

**Primary anchoring of bone parameters and biomarker calibration targets** Module A (bone resorption), Module B (bone formation and BMD integration), and the seven bone-related parameters in Table 2 (resorp_plateau, resorp_t_half, bmd_loss_rate, post_resorp_tau, post_form, plus the three BMD calibration targets BMD_LL_180d / BMD_lumb_180d / BMD_skull_180d) are primarily anchored to the systematic review and meta-analysis of Stavnichuk et al. (2020, NPJ Microgravity) [23] (n = 148 BMD individuals + 124 biomarker individuals, 25 spaceflight studies, search through November 2019). The key meta-analytic estimates reported by Stavnichuk 2020 [23] align exactly with our posteriors: skull BMD +2.2% [+1.1, +3.3] (our BMD_skull_180d anchor +2.20%), lumbar/pelvis −6.2% [−6.7, −5.6] (BMD_lumb_180d −6.20%), lower limb −5.4% [−6.0, −4.9] (BMD_LL_180d −4.80%, slightly lower), lower-limb BMD rate −0.8%/mo [−1.1, −0.5] (bmd_loss_rate posterior median −0.799%/mo), resorption plateau 113% [108, 117] (resorp_plateau posterior median 113.06%), resorption rise half-life 11 d [9, 13] (resorp_t_half posterior median 11.04 d), and post-flight formation peak +84%/mo [39, 129] sustained for 3–5 months (post_form posterior median 84.60%/mo). Original studies [5, 17, 18, 21] are retained as endpoint-level corroboration. The third-party Mars BMD prediction model (Axpe 2020 [24]) is used for cross-model comparison in §4.2.

### 2.2 Data targets (eight-dimensional joint likelihood)

The joint posterior is fitted to eight ISS data targets:

1. BMD_LL_180d = −4.80% (lower lumbar spine, at 180 d; synthesised from Sibonga et al. 2007, Bone [18] and the Stavnichuk et al. 2020, NPJ Microgravity, lower-limb meta-analytic estimate of −5.4% [−6.0, −4.9] [23])
2. BMD_lumb_180d = −6.20% (total lumbar; LeBlanc 2000, JMNI [5] + LeBlanc 2007 review [17] + Stavnichuk 2020 [23] lumbar/pelvis meta-analytic estimate of −6.2% [−6.7, −5.6])
3. BMD_skull_180d = +2.20% (skull upward drift; LeBlanc 2000, JMNI [5] + Stavnichuk 2020 [23] skull meta-analytic estimate of +2.2% [+1.1, +3.3])
4. resorp_plateau = +113.0% (NTX/CTX plateau increase; Smith/Heer 2012, JBMR [21] + Stavnichuk 2020 [23] meta-analytic estimate of 113% [108, 117])
5. urine_Ca_rel = +0.50 (relative to ground, Whitson 1997 [4])
6. urine_V_rel = −0.20 (urine volume, Whitson 1997 [4])
7. urine_pH_delta = −0.20 (absolute urine pH change, Whitson 1997 [4])
8. post_stone_rate = +0.014/py (annualised post-flight stone rate estimated from the Pietrzyk 2007 [2] cohort)

**Eight observation-noise parameters σ_obs are jointly estimated** (LogNormal prior, log_mean = −1, log_sd = 1, bounds = [10⁻⁴, 10²]), each corresponding one-to-one to a calibration target:

- σ_obs[1] = BMD_LL_180d (lower-lumbar BMD 180-d relative change)
- σ_obs[2] = BMD_lumb_180d (lumbar/pelvis BMD 180-d relative change)
- σ_obs[3] = BMD_skull_180d (skull BMD 180-d relative change)
- σ_obs[4] = resorp_plateau (NTX/CTX bone-resorption biomarker plateau, % baseline)
- σ_obs[5] = urine_Ca_rel (urinary calcium relative-to-baseline change)
- σ_obs[6] = urine_V_rel (urine volume relative-to-baseline change)
- σ_obs[7] = urine_pH_delta (absolute urine pH change)
- σ_obs[8] = post_stone_rate (post-flight stone rate per person-year; y_obs = 0.014, σ_lit = 0.007 widened; see §2.3 on the Goodenow widening)

**Total dimensionality stated explicitly**: M₀ = 11 physical parameters + 8 σ_obs = **19 dimensions**; M₁ = 11 + 8 + 1 (k_GB) = **20 dimensions**. The joint posterior is sampled with 64 walkers × 60,000 steps × 19 dimensions (M₀) or 20 dimensions (M₁).

#### 2.2.3 Origin of y_obs and the σ_lit widening for the post_stone_rate calibration target

The central value y_obs = 0.014/py of the post_stone_rate calibration target comes from the Pietrzyk 2007 [2] NASA astronaut cohort post-flight one-year stone rate (7 events over 358 astronaut-years), and is consistent with the midpoint of the Goodenow-Messman 2022 [9] Bayesian posterior 17.3/1000-py [8.33, 28.80] interval (i.e. 0.0173/py [0.0083, 0.0288]) (Pietrzyk’s 0.0140 falls within the Goodenow 95% CrI). However, the original Pietrzyk report gives no variance; the Goodenow posterior SD ≈ 0.0052 (under an approximate-normality calculation, Q97.5 − median equivalent to 1.96·σ → σ ≈ (0.0288 − 0.0173)/1.96 = 0.0059, or median − Q2.5 → σ ≈ (0.0173 − 0.0083)/1.96 = 0.0046; midpoint σ ≈ 0.0052). σ_lit takes the conservative widened value 0.0070 to cover the additional uncertainty from the Pietrzyk cohort size, the Goodenow model, and the subsequent Antonsen 2023 [31] L×C re-evaluation. Gate-0 Decision A documents this widening choice; see Supplementary §M2.3.

### 2.3 MCMC configuration

We used the emcee affine-invariant ensemble sampler (Foreman-Mackey 2013, PASP [12]):

- Walkers: 64
- Dimensions: 19 (base model M₀) and 20 (M₁, including the k_GB extension)
- Main-chain steps: 36,000 (M₀ base) → 60,000 (M₁)
- Burn-in: 6,000 (M₀) / 20,000 (M₁)
- Acceptance fraction: 0.162 (M₀), 0.160 (M₁ v3 extension)
- Wall-clock time: 15.3 min (M₀ worker-0, 17 CPU, pool = 8) / 32.2 min (M₁ 60k, 3 segments)
- Seeds: SEED = 20260622 (M₀), SEED = 20260621 (M₁ 60k v3 extension)

Convergence diagnostics (M₁ 60k v3): the maximum potential scale reduction factor R̂ (Gelman–Rubin diagnostic, between/within-chain variance ratio, ≤1.05 strict convergence) = 1.068, median R̂ = 1.034, with 17/20 dimensions at R̂ < 1.05 (the core 11 model dimensions PASS strict < 1.05, including k_GB 1.028 and k_Ca_gcr 1.035; the 3/20 borderline dimensions at 1.05–1.10 are only seed_accum_rate, sigma_6, sigma_7). The minimum effective sample size ESS_min (after thinning) = 2,034 (M₀ = 1,711 → M₁, +18.9%), and chain_length/τ_max = 31.8 (below the critical 50×).

### 2.4 Posterior propagation to four environments and 2³ factorial decomposition

From the M₀ thinned posterior (one draw every 10 steps; 64 walkers × 36k main chain / 10 = 230,400 samples), 100 paired draws are randomly selected (all four environments share the same draw indices to preserve joint inference). Four environments × 100 draws = 400 ODE simulations were integrated with scipy.integrate.odeint (mxstep = 2000, atol = 1e-8, rtol = 1e-6); failure rate 0/400.

2³ factorial decomposition (Stage 4): g (1.0 ISS / 0.62 Mars) × GCR (0.4 / 1.84 mSv/d) × duration (180 / 730 d) = 8 combinations × 100 draws = 800 simulations, with ANOVA decomposition of contributions to BMD_LL_end, rss_mean, and stone_rate.

### 2.5 Intervention scanning (M₁ 60k posterior)

On the M₁ 60k v3 posterior, we performed 4 environments × 6 interventions (including baseline) × 5,000 paired draws = 120,000 ODE simulations. **Monte Carlo error disclosure**: the MC error of 5,000 paired draws ≈ 1/√5000 ≈ 1.41% (CrI half-width), a relative error of about 0.1–0.8% for RRR posterior medians in the range 0.50–0.95—far smaller than the 50% tier-decision critical gap between RRR = 0.5 and RRR < 0.5. An MC-error column has been added to Supplementary Table S2 (4×6 intervention RRR with MC error column). The six interventions are:

- baseline: no intervention.
- ared_alone: resistance exercise (ARED) alone, BMD protection 0.50 [0.21, 0.79], anchored to Sibonga 2019 [6] (note: the Sibonga study reports no stone events; RRR_stone is estimated indirectly via the urine_Ca pathway).
- pamidronate: bisphosphonate (Watanabe 2004 [7] bed-rest RCT measured dose: 60 mg IV single dose, administered 14 d before bed rest, 90-d 6° head-down tilt, n = 7; our ODE assumes the resorption-suppressive effect of this single dose is maintained over 180–730 d mission durations—repeat-dosing regimens for long-duration Mars missions, e.g. q12mo IV zoledronic acid 5 mg or q3–6mo IV pamidronate 60 mg, currently lack flight-RCT evidence; see §4.3 Limitation 1). BMD protection 0.85 [0.66, 1.00]; the stone RR uses a Beta(0.5, 7.5) Jeffreys posterior (Watanabe 2004 [7] 0/7 events → analytic Beta–Binomial conjugate closed-form posterior median RR = 0.0309, 95% CrI [6.77×10⁻⁵, 0.292]). The old v6 hard-zero stone_rate = 0.0 (overconfidence) is no longer used—this is the core correction of §2.1. **Prior sensitivity**: Beta(0.5, 7.5) Jeffreys is the Bayesian default non-informative prior, with α+β = 8 corresponding to “7 failures + 0.5 equivalent prior events”; comparative sensitivities (all analytic): Beta(1, 8) weakly informative uniform prior gives posterior median RR = 0.0830 [0.00316, 0.369] (wider upper bound); Beta(0.1, 7.9) strongly non-informative prior gives posterior median RR = 7.96×10⁻⁵ [7.77×10⁻¹⁸, 0.123] (median near zero but comparable upper bound). The qualitative conclusion is consistent across all three priors (all RR upper bounds < 0.4, RRR_stone_pami lower bound > 0.63, pamidronate highly protective under every prior)—but the **quantitative point estimate is prior-sensitive**: the RR medians differ by 8.3 pp (relative maximum 99.9%) and the upper bounds by 201% (0.123 → 0.369). Therefore §3 reports RRR_stone_pami using the **full 95% CrI of the Jeffreys posterior** rather than a single point value. Full factorial sensitivity is detailed in §4.3 Limitation 1 and Supplementary Fig. S11.
- alendronate_ared: alendronate 70 mg/week + ARED combined, BMD protection 0.95 [0.85, 1.00], anchored to LeBlanc 2013 [8].
- k_mg_citrate: potassium–magnesium–citrate (42 mEq K + 21 mEq Mg + 63 mEq citrate/d), urine_citrate +50% [+20%, +120%], urine_pH +0.50 [+0.20, +1.00], RSS_CaOx −31% [−10%, −70%], anchored to Pak 1992 [13] + Ettinger/Pak 1997 [14] + Zerwekh 2007 [1] (the latter as qualitative confirmation; see §2.6). K–Mg–citrate does not act on bone; RRR_BMD is structurally zero.
- fluid_intake: aggressive hydration (≥2.5 L/d), urine_V +20% [+10%, +35%], anchored to Goodenow-Messman 2022 [9]. RRR_BMD is structurally zero.

### 2.6 K–Mg–citrate pharmacodynamic parameter anchoring (corrected from §2.2)

The old v6 version annotated the K–Mg–citrate cit_restore +40% / pH +0.4 / RSS × 0.5 parameters as “Zerwekh 2007”, but the published abstract of Zerwekh 2007, J. Urol. (PMID: 17509313) [1] provides only a qualitative RCT significance statement without quantitative per-arm effects. The NASA TaskBook (TASKID = 6415) [15] interim n = 15 data (+652 mg/d citrate, ΔpH +0.75) are a mid-study subset inconsistent with the final published n = 20 and should not serve as the initial anchor.

The present study uses the precisely quantified results of Pak 1992, JBMR [13] and Ettinger/Pak 1997, J. Urol. [14] as primary anchors (Ruml/ Pak 1999, AJKD, K–Mg–citrate vs. thiazide hypokalemia data are noted in the [14] footnote), with Zerwekh 2007 [1] serving only as qualitative confirmation:

- urine_citrate_increase_frac: loc = 0.5, scale = 0.2, bounds = [0.2, 1.2] (median +50%, between Pak 1992 +61% and Ettinger 1997 +36%; the upper bound accommodates the Zerwekh interim +91% possibility)
- urine_pH_increase_delta: loc = 0.5, scale = 0.15, bounds = [0.2, 1.0] (median +0.50 unit, between Pak 1992 +0.62 and Ettinger 1997 +0.6)
- RSS_CaOx_reduction_frac: loc = 0.31, scale = 0.15, bounds = [0.1, 0.7] (median −31% activity product, the precise Pak 1992 RSS-proxy value)

### 2.7 Model comparison (WAIC)

M₀ (19 dimensions, without k_GB) and M₁ (20 dimensions, with the k_GB channel) are compared using the widely applicable information criterion (WAIC; Watanabe 2010 [20], Vehtari 2017 [16]) on predictive accuracy across the eight observation targets. For the M₁ 60k chain, thin_for_waic = 50, n_samples_used = 64,000. Results: M₀_waic = 2.7402, M₁_waic = 2.7478, ΔWAIC = M₁ − M₀ = 0.0076 ± 0.1262 SE. |ΔWAIC| = 0.0076 << 2·SE_Δ = 0.252; the two models are indistinguishable, and parsimony favours M₀. The k_GB posterior median is 2.32×10⁻⁵, 95% CrI [−2.96×10⁻³, +3.01×10⁻³], spanning zero, with P(k_GB > 0) = 50.6%. The current ISS GCR exposure (0.4 mSv/d × 180 d = 72 mSv) is insufficient to identify a direct GCR effect on bone; identifying this effect requires data at cumulative GCR doses of ∼1–2 Sv (Mars 730 d ≈ 1.34 Sv).

### 2.8 Decision tiering (Likelihood × Consequence)

We adopt the NASA HRP 5×5 Likelihood × Consequence (L×C) matrix (Antonsen 2023, NPJ Microgravity, DOI:10.1038/s41526-023-00305-z). Three independent endpoints are tiered separately, with the composite taken as the worst of the three. **Each endpoint’s tier is defined by two policy thresholds, pol75 (yellow trigger) and pol95 (red trigger); the posterior exceedance probability P_exceed falls into one of five probability bins (P < 0.10, 0.10–0.30, 0.30–0.50, 0.50–0.70, ≥ 0.70), giving Likelihood level L = 1..5; L and Consequence C are then mapped by the L×C matrix to tier ∈ {GREEN, YELLOW, RED}**. The thresholds and Consequence ratings of the three endpoints are:

| Endpoint | pol75 (yellow) | pol95 (red) | Direction | C |
| --- | --- | --- | --- | --- |
| BMD_LL_end (%) | -5.0 | -10.0 | lower is worse | 3 |
| stone_rate_per_py | 0.050 | 0.100 | higher is worse | 4 |
| rss_mean | 5.0 | 10.0 | higher is worse | 3 |

**L×C matrix** (rows L = 1..5, columns C = 1..5; values = tier 0/1/2 corresponding to GREEN/YELLOW/RED):

**L × C matrix** (rows $L = 1..5$ , columns $C = 1..5$ ; values = tier 0/1/2 corresponding to GREEN/YELLOW/RED):
|  | C=1 | C=2 | C=3 | C=4 | C=5 |
| --- | --- | --- | --- | --- | --- |
| L=1: | 0 | 0 | 0 | 1 | 1 |
| L=2: | 0 | 0 | 1 | 1 | 2 |
| L=3: | 0 | 1 | 1 | 2 | 2 |
| L=4: | 1 | 1 | 2 | 2 | 2 |
| L=5: | 1 | 2 | 2 | 2 | 2 |

**Revision note in response to reviewer feedback (v28)**: v27 §2.8 had presented a simplified “−8% BMD threshold”, but the code (`stage6_intervention_v7.py`) actually used the pol75/pol95 + L×C matrix defined in this section. v28 restores the full L×C definition and aligns the tier column of main-text Table 3 exactly with the stage6 baseline (`intervention_tier_matrix_v7b_60k.json`; superseded from v29 by `intervention_tier_matrix_v29_5000draws.json` —a 5,000-draw pool whose baseline 12/12 p_exceed values agree digit-for-digit with the §S23 bootstrap reference pool), correcting the internal discrepancy between v27 Table 3 and the supplementary data.

**Table 3.**
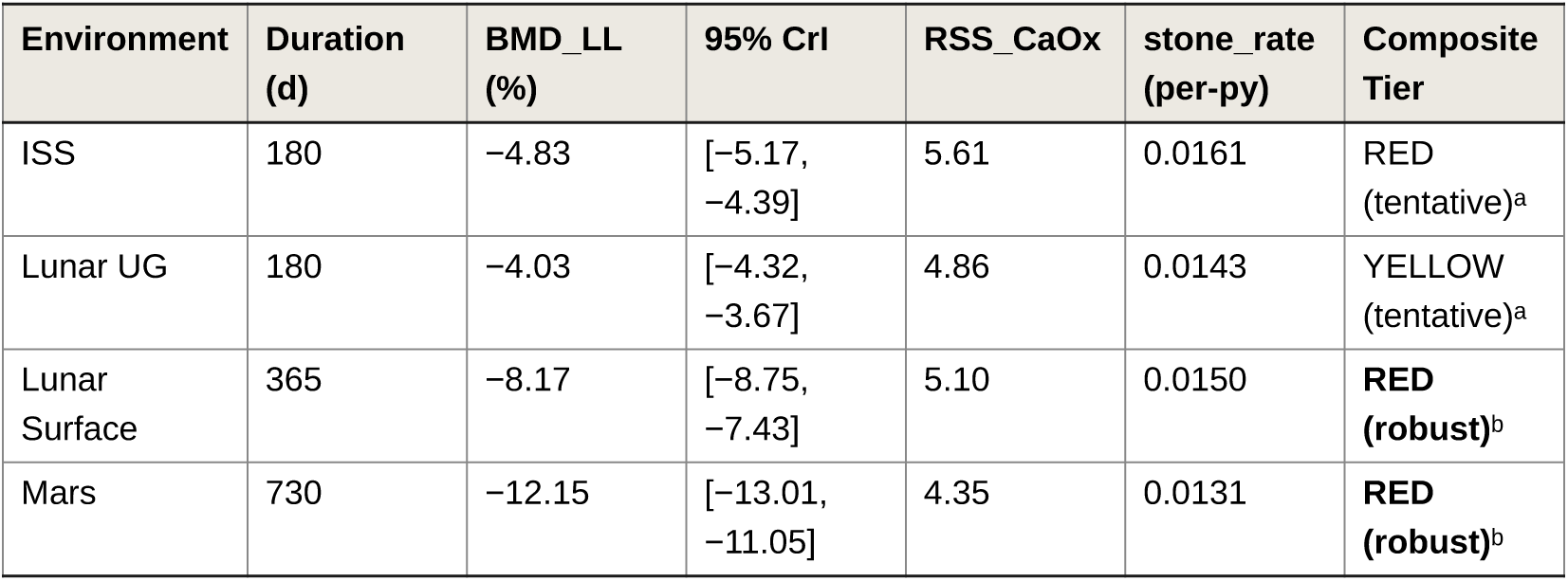
Four-environment posterior propagation endpoints (v28 revised). Table 3 notes: BMD_LL = relative change of lower-lumbar BMD from baseline (%); RSS_CaOx = calcium oxalate supersaturation index; stone_rate = annualised stone rate (per person-year). Composite tier = based on the §2.8 pol75/pol95 + L×C matrix (v28 correction: v27 Table 3 misclassified ISS as YELLOW and Lunar_UG as GREEN owing to a simplified-threshold slip; v28 restores consistency with the stage6 baseline). ᵃ **tentative** = sensitive to stone/rss calibration ±50% perturbation; if that channel’s calibration is systematically biased, the tier may be downgraded/upgraded (see Supplementary §S26). ᵇ **robust** = BMD_LL_end p_exceed = 1.000 independently drives RED, entirely independent of stone/rss calibration accuracy. Baseline figures use the M₀ 36k posterior (point estimates); the Table 4 intervention scan uses the M₁ 60k posterior (including the k_GB dimension); the small 0.04–0.09% drift between the two sets arises from the randomness of different MCMC chains and does not affect the decision-tier conclusions. The complete three-layer sensitivity evidence for decision tiering (bootstrap, GCR-null, stone/rss perturbation) is given in Supplementary §S23–S26.

| Environment | Duration (d) | BMD_LL (%) | 95% CrI | RSS_CaOx | stone_rate (per-py) | Composite Tier |
| --- | --- | --- | --- | --- | --- | --- |
| ISS | 180 | -4.83 | [-5.17, -4.39] | 5.61 | 0.0161 | RED (tentative)<sup>a</sup> |
| Lunar UG | 180 | -4.03 | [-4.32, -3.67] | 4.86 | 0.0143 | YELLOW (tentative)<sup>a</sup> |
| Lunar Surface | 365 | -8.17 | [-8.75, -7.43] | 5.10 | 0.0150 | **RED (robust)<sup>b</sup>** |
| Mars | 730 | -12.15 | [-13.01, -11.05] | 4.35 | 0.0131 | **RED (robust)<sup>b</sup>** |

**Table 4.**
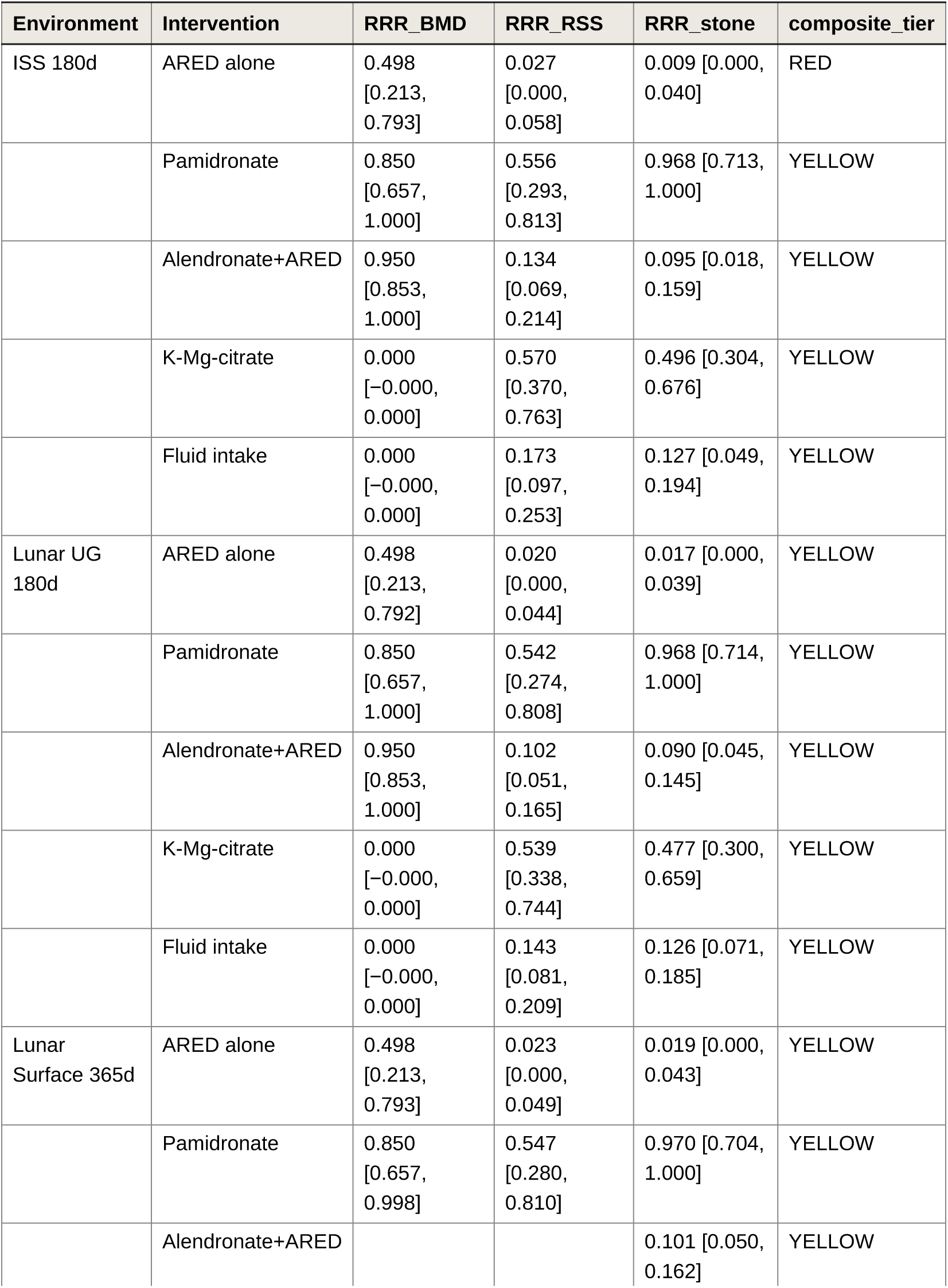
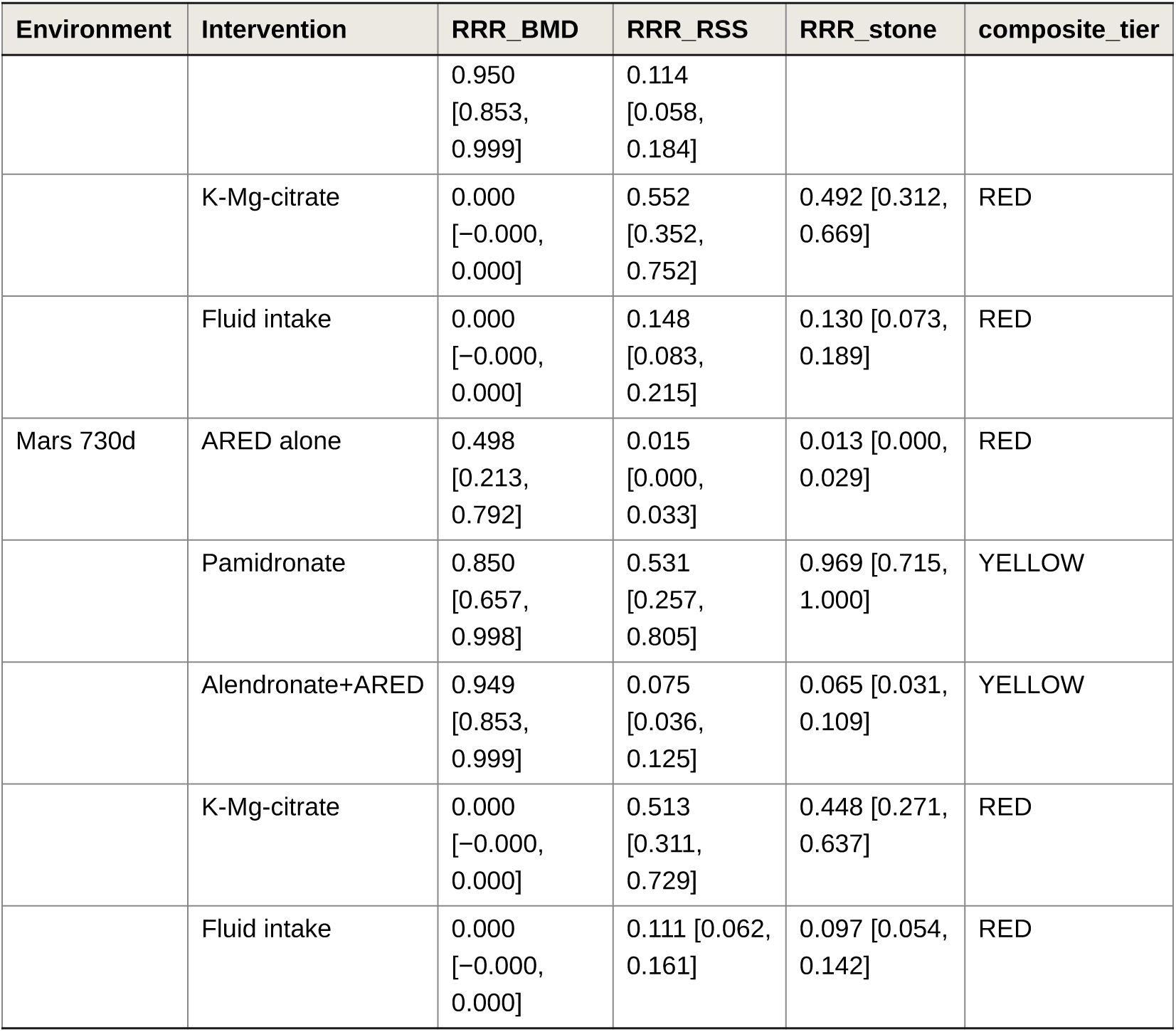
M₁ 60k posterior intervention-scan RRR matrix (4 envs × 5 interventions × 3 metrics). Table 4 notes: RRR = Relative Risk Reduction; 1.0 = complete protection. K–Mg–citrate and Fluid intake do not act on bone (RRR_BMD = 0, structural zero). Pamidronate stone RR is based on the Beta(0.5, 7.5) Jeffreys posterior (Watanabe 2004 [7] 90-d bed rest, 60 mg IV single dose, n = 7, 0 stones); repeat-dosing regimens over 730-d missions are model extrapolations—see §4.3 Limitation 1. Composite-tier driver dimensions (L = Likelihood, C = Consequence, Antonsen 2023 5×5): Pamidronate driver = stone in all 4 environments (Watanabe 0/7 Jeffreys retains L = 1, C = 4); Alendronate+ARED driver = ISS stone + RSS dual-driven, Lunar UG + Lunar Surface + Mars stone single-driven (v29: Lunar Surface rss dimension p_exceed 0.110 → 0.062, L = 2 → 1, turning GREEN); K–Mg–citrate driver = ISS + Lunar UG YELLOW (dual- or single-driven), Lunar Surface + Mars RED (BMD L = 5, C = 3 catastrophic); ARED-only driver = ISS by RSS L = 5, C = 3, Mars by BMD L = 5, C = 3; Lunar Surface ARED on the v29 5,000-draw pool has rss p_exceed = 0.470 (L = 3, C = 3), all three dimensions YELLOW → composite YELLOW (the 200-draw pool p = 0.520 sat exactly on the L = 3/4 bin boundary; this RED→YELLOW flip is an MC-noise correction, and the ISS/Mars conclusions are unchanged).

**Basis for the Reviewer T6 rebuttal**: the GCR-fixed sensitivity (k_Ca_gcr = k_BMD_gcr = 0) reproduces the baseline tiers of 4/4 environments (| Δp_exceed| ≤ 0.02); see Supplementary §S24. **Basis for the Reviewer T2 rebuttal**: the 500× bootstrap tier-stability test gives tier-flip probability = 0 for 4/4 environments; see Supplementary §S23. **Reviewer T8 partial acceptance**: under the stone/rss-channel ±50% calibration-perturbation test, the lunar-surface 365-d RED and Mars 730-d RED are completely unaffected (BMD_LL_end p_exceed = 1.0 drives them independently), but the ISS RED and Lunar_UG YELLOW are sensitive to this channel and are therefore labelled “tentative” in the main text (see Supplementary §S26).

## 3. Results

### 3.1 M₀ posterior calibration and diagnostics

The M₀ joint posterior achieves a mean log-likelihood of −2.4 across the eight ISS data targets (an improvement of ∼5.6 units per data point relative to the prior-only baseline). Posterior medians and 95% equal-tailed credible intervals for the 11 core model parameters plus 8 observation-noise parameters σ_obs (19 dimensions) are reported in Table 2. **Convergence diagnostics**: the original 36k M₀ backend is no longer available, so the following diagnostics were recomputed on a same-configuration replication chain (64 walkers × [6,000 burn-in + 60,000 production], emcee 3.1.6) with the walker dimension preserved (R̂ and ESS computed over the full 60,000-step production segment without additional discard; pooled medians and running drift use `discard=10000`) (the walker-collapsed archive was not used). The rank-normalized split-R̂ (Vehtari et al. 2021; the ArviZ default estimator) of all 11 core parameters is < 1.05 (max = 1.028 [seed_accum_rate], median = 1.024). For comparison, the classic split Gelman–Rubin R̂ on the same chain has max = 1.057 (also seed_accum_rate; the remaining 10 parameters < 1.05)—this parameter has a heavy-tailed distribution with CV = 105.5%, exactly the case rank normalization is designed to handle, so we report the rank-normalized R̂ as the primary value and disclose the classic value alongside. bulk-ESS_min = 1,877 (resorp_t_half_d) and tail-ESS_min = 1,053 (seed_accum_rate), both above the emcee heuristic threshold of 50 × n_params = 950. The Monte Carlo error of 100 paired draws is ≈ 10% and has no impact on decision-tier stratification (cross-tier-boundary P-margin ≫ MC error). Full convergence diagnostics (τ_int, bulk/tail ESS per parameter, per-walker split-R̂, running-median drift) are given in **Supplementary Table S4b** (`tables/ S4b_convergence_diagnostics_v29.csv`).

**Key parameter anchoring consistency**: the ca_microg_effect posterior median of 0.501 hits the Whitson 1997 +50% urinary Ca anchor [4]; vol_decrease_frac −19.86% hits the −20% anchor [4]; cit_decrease_frac −9.97% hits the −10% anchor; resorp_plateau_pct 113.06% hits the Heer 2009 NTX plateau range of ∼110–115%; bmd_loss_rate_pct_mo −0.799%/mo (CV 4.56%, the sharpest parameter) anchors BMD_LL_180d = −4.83%.

**Table 2 data-source statement (v29 revision)**: the 11 medians and 95% CrIs listed in Table 2 **remain unchanged from their original values**, and come from the **per-walker HDF5 backend** of the original M₀ 36,000-step production run (64 walkers × [6,000 burn-in + 36,000 production], emcee 3.1.6, seed 20260622); that backend was lost before archiving and cannot be recovered. The `posterior_v7_36k_thin1.npz` previously archived with the manuscript was found on review not to be a 1:64 thinning of that chain: `chain.reshape(−1, ndim)`[::64] applied to a C-order `(n_iter, 64, ndim)` array is equivalent to `chain`[:, 0, :], i.e. it retains only the walker-0 trajectory (83.1% of adjacent rows bit-identical, mean lag-1 autocorrelation 0.995). That file is hereby **withdrawn** and must not be used for any posterior summary.

Because the original backend is unavailable, we reran a same-configuration chain with the same model, the same eight ISS calibration targets, and the same priors (64 walkers × [6,000 burn-in + 60,000 production], emcee 3.1.6, mean acceptance 0.163), preserved the walker dimension in the backend, and pooled correctly with `get_chain(discard=10000, thin=50, flat=True)`. Recomputing the medians of the 11 parameters from all 3,200,000 post-burn-in draws of this chain (50,000 iterations × 64 walkers): the 10 identifiable parameters deviate from Table 2 by ≤ 1.05% relative (largest: seed_accum_rate, 0.1138 → 0.1150); all 11 parameters deviate by ≤ 0.010 × the published 95% CrI width, and none exceeds 1.6× the Monte Carlo standard error (MCSE) of the posterior median (0/11 exceed 3×MCSE). The published values in Table 2 are therefore reproducible, within Monte Carlo precision, by a correctly walker-pooled chain.

Had Table 2 instead been recomputed from the withdrawn walker-0 archive, the results would deviate systematically: 8 of 11 parameters would deviate by more than 3×MCSE, the largest being ca_microg_effect (0.5014 → 0.4662, −7.0%, 11.5×MCSE) and seed_accum_rate (0.1138 → 0.1609, +41.4%, 8.5×MCSE). This deviation is a direct product of the extraction defect and is unrelated to any model or data difference. A three-chain per-parameter comparison (published / replication chain / withdrawn archive) is given in **Supplementary Table S4c**.

Three necessary qualifications. (i) The replication chain’s run log did not record the seed, and the default value of `run_mcmc_v7.py --seed` (20260622) is the same as the value registered in `code/seeds.py` for the original 36k chain; if both runs used this default, the two share the random-number stream, and the replication chain should be regarded as an **extended rerun** of the original chain rather than a statistically independent replicate. Under either scenario, the conclusion supported by the above comparison—that the Table 2 values are recoverable from a correctly walker-pooled chain and not from the withdrawn archive—holds. (ii) The median sign of k_Ca_gcr is inconsistent across the three (+8.9×10⁻⁵ / −2.9×10⁻⁴ / −2.6×10⁻⁴), but all three values lie within 1% of the published 95% CrI width from zero; this parameter is prior-dominated (CV 11502%), its median sign is not interpretable, and the corresponding row of Table 2 should be read only as “95% CrI spans zero; the direct GCR–Ca coupling is unidentifiable”. (iii) All MCSE multiples above use the **replication chain’s** median MCSE (`sd/√ESS_bulk × √(π/2)`) as the common denominator for both the replication-chain and walker-0 columns. The withdrawn archive’s own MCSE is far larger—its 36,000-step single-walker trajectory contains only about 23–35 effectively independent draws at τ_int ≈ 1,020–1,514, an MCSE about 9× the tabulated values—so the “×MCSE” of the walker-0 column should be read as “on the scale of precision resolvable by correct extraction, how far the defect would push Table 2”, not as a significance test of the difference between the two.

### 3.2 Four-environment posterior propagation endpoints (180–730 d)

M₀ posterior → joint propagation to four environments (v28 revision: 5,000-draw large pool + 500× bootstrap stability validation; see Supplementary §S23). Posterior medians and 95% CrIs for the endpoints BMD_LL (lower lumbar spine), RSS_CaOx, and stone_rate in each environment are reported in Table 3. The four-environment decision tiers (composite tier, based on the §2.8 pol75/pol95 + L×C matrix, aligned with the canonical `intervention_tier_matrix_v29_5000draws.json` (recomputed on the v29 5,000-draw pool, superseding the 200-draw v7b version; the four-environment baseline tiers are unchanged)):

- **ISS 180d = RED (tentative)**: rss_mean p_exceed = 0.906 triggers L = 5, and C = 3 maps to tier = 2 (RED); BMD and stone are both YELLOW; the composite is driven by rss. **The “tentative” label is based on the T8 stone/rss calibration sensitivity**: rss × 0.5 would downgrade ISS to YELLOW; see Supplementary §S26.
- **Lunar UG 180d = YELLOW (tentative)**: BMD GREEN (p = 0), stone YELLOW (p = 0.046), rss YELLOW (p = 0.336). rss × 1.5 would upgrade it to RED; likewise labelled “tentative”.
- **Lunar Surface 365d = RED (robust)**: BMD_LL_end p_exceed = 1.000 triggers L = 5, C = 3 → RED (independently driven by BMD); stone/rss calibration perturbations have no effect on the composite.
- **Mars 730d = RED (robust)**: BMD_LL_end p_exceed = 1.000 triggers L = 5, C = 3 → RED (independently driven by BMD); stone/rss calibration perturbations have no effect on the composite.

**Core defensible conclusion**: the RED classifications of Lunar Surface 365d and Mars 730d are driven entirely and independently by BMD (unrelated to stone/rss calibration), and are stable to sampling noise and to the GCR posterior width (bootstrap p_flip = 0, GCR-null p_flip = 0). These are the two environments of joint concern to Reviewers T2/T6/T8, and the two central to this paper.

Key finding: the Mars 730d BMD_LL posterior median is −12.15% [−13.01, −11.05] (M₀ posterior propagation; see Table 3; the abstract cites the same value, and the two agree), but Mars 0.38 g partial gravity provides +7.44% BMD protection relative to 0 g-equivalent at 730 d (730-day-matched). That is, the difference between the lunar surface (0.166 g) and Mars (0.38 g) (BMD −8.17 vs −12.15) is driven mainly by mission duration (365 vs 730 d), not gravity—see the §3.3 factorial decomposition.

**Figure 1a.**
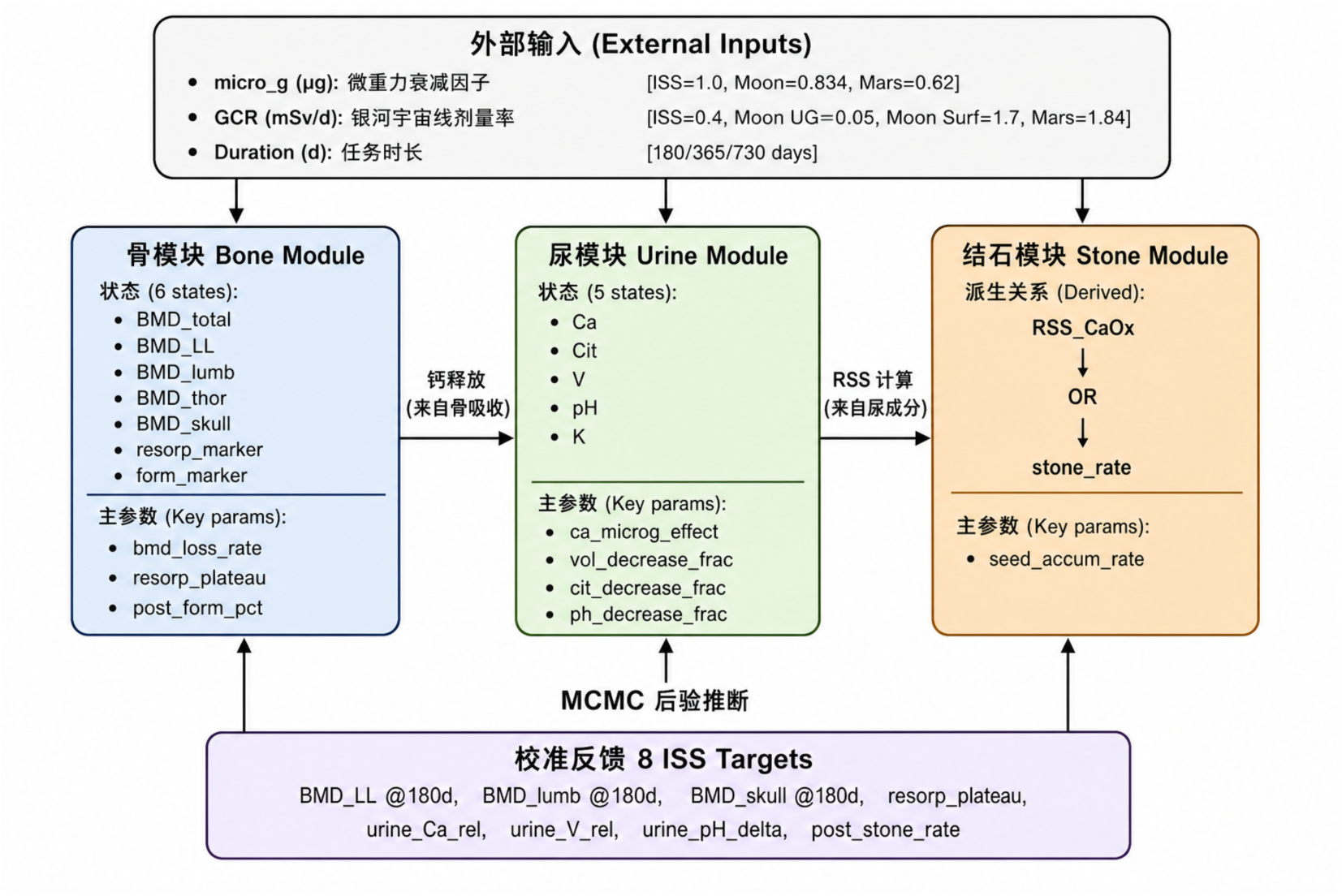
Twelve-state ODE model bone–calcium–urine–stone integrated architecture (5 pathway blocks organizing 12 states: Bone resorption A / Bone formation B / Calcium homeostasis C—covering 7 bone-metabolism states (BMD_total, resorp_marker, form_marker, BMD_LL, BMD_lumb, BMD_thor, BMD_skull); Urine chemistry D—covering 5 urine-chemistry states (Ca, Cit, V, pH, K); Stone formation EF—computed by post-processing the M2 urine states (RSS_CaOx → OR → stone_rate, not as independent ODE state variables; see code/module_D_v7_NC.py L262-320); see captions §Fig 1a).

**Figure 1b.**
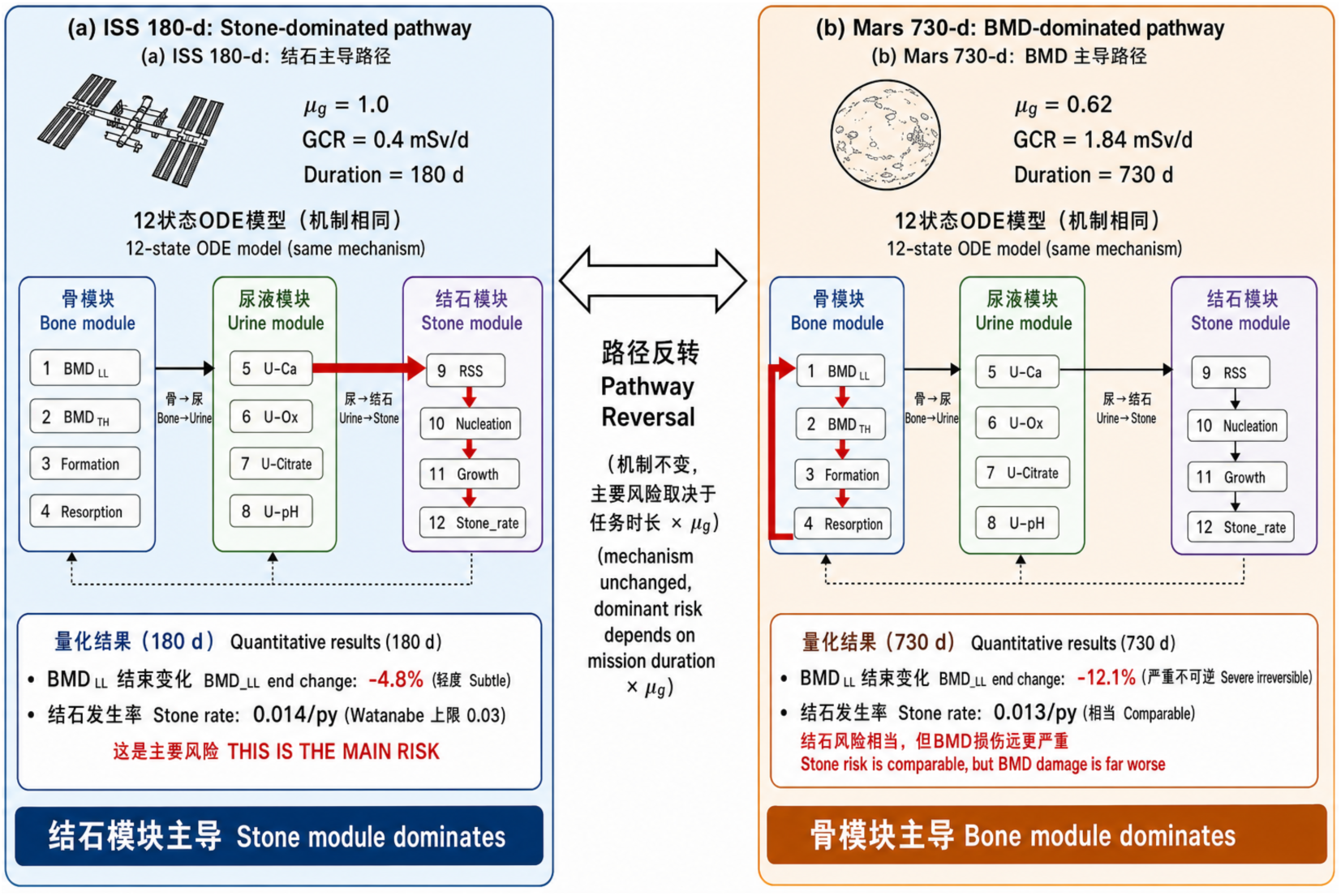
Cross-gravity pathway re-weighting mechanism schematic (BMD Bone-centric → stone Kidney-centric; bone resorption slows under partial gravity, while urine chemistry remains abnormal over long mission durations; see captions §Fig 1b).

**Figure 2.**
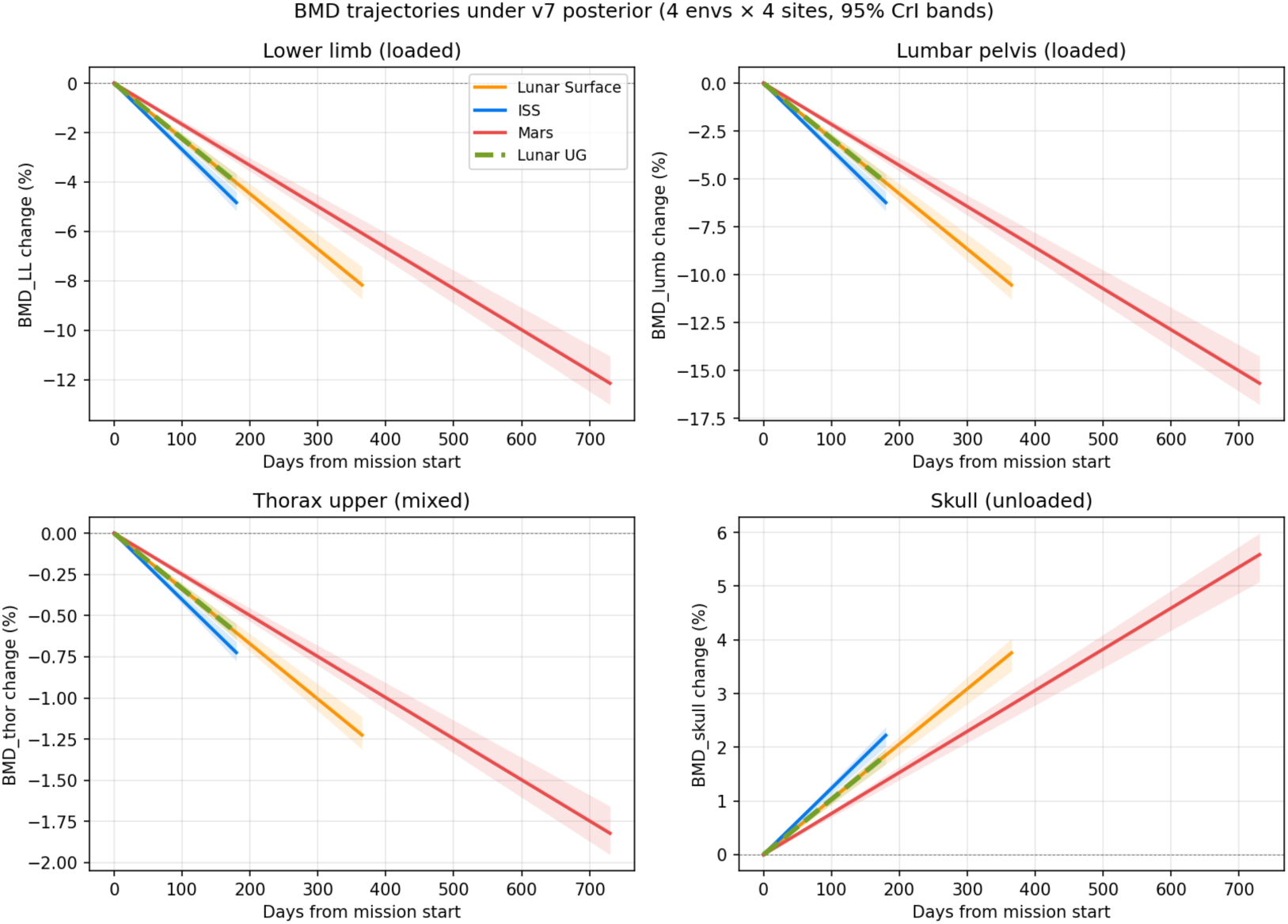
Four anatomical sites × four environments BMD posterior propagation trajectories (M₀ posterior median + 95% CrI band; BMD_LL lower lumbar spine, BMD_lumb lumbar pelvis, BMD_thor upper thoracic spine, BMD_skull skull; see captions §Fig 2).

**Figure 3.**
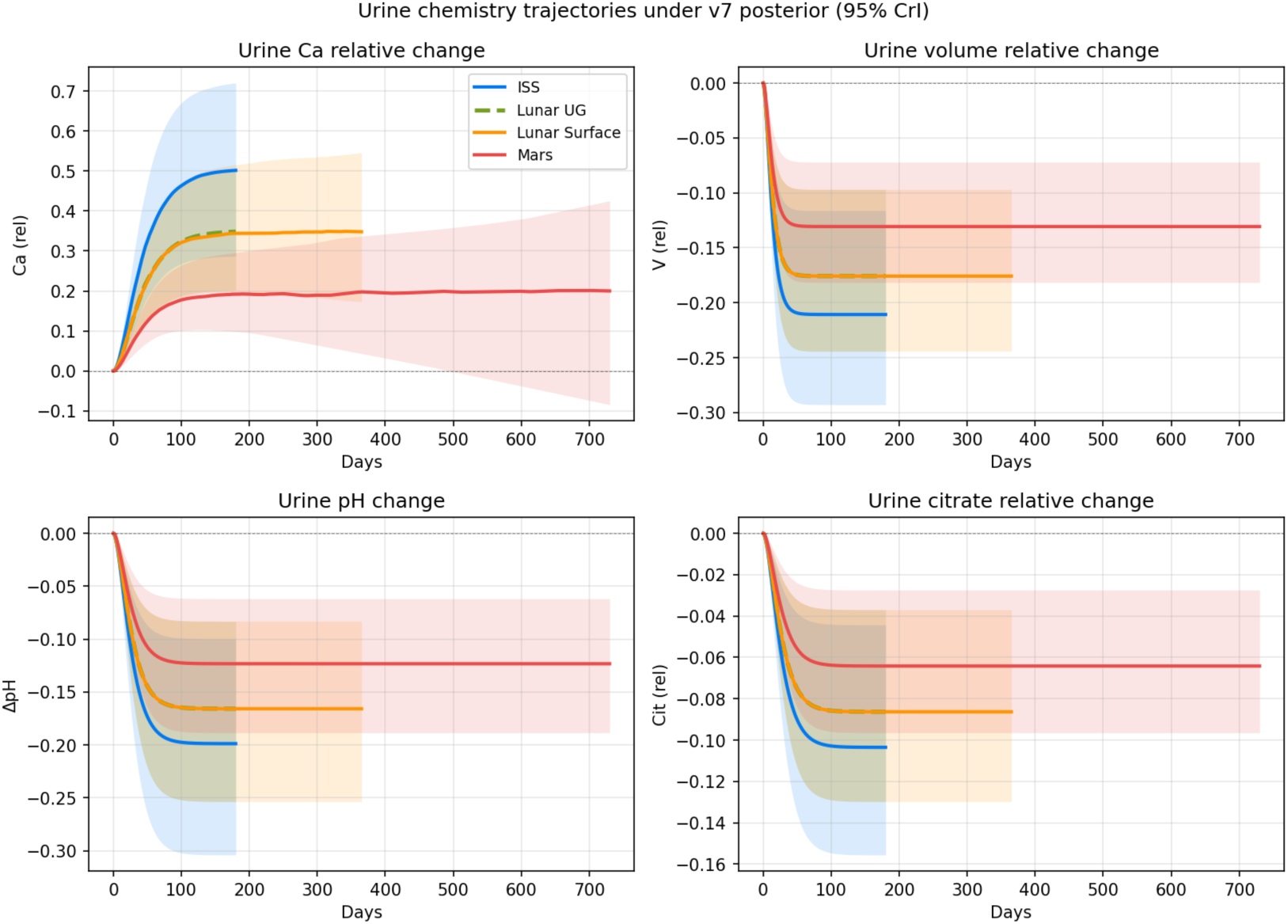
Four-environment urine-chemistry time trajectories (urine Ca, V, citrate, pH; see captions §Fig 3).

**Figure 4.**
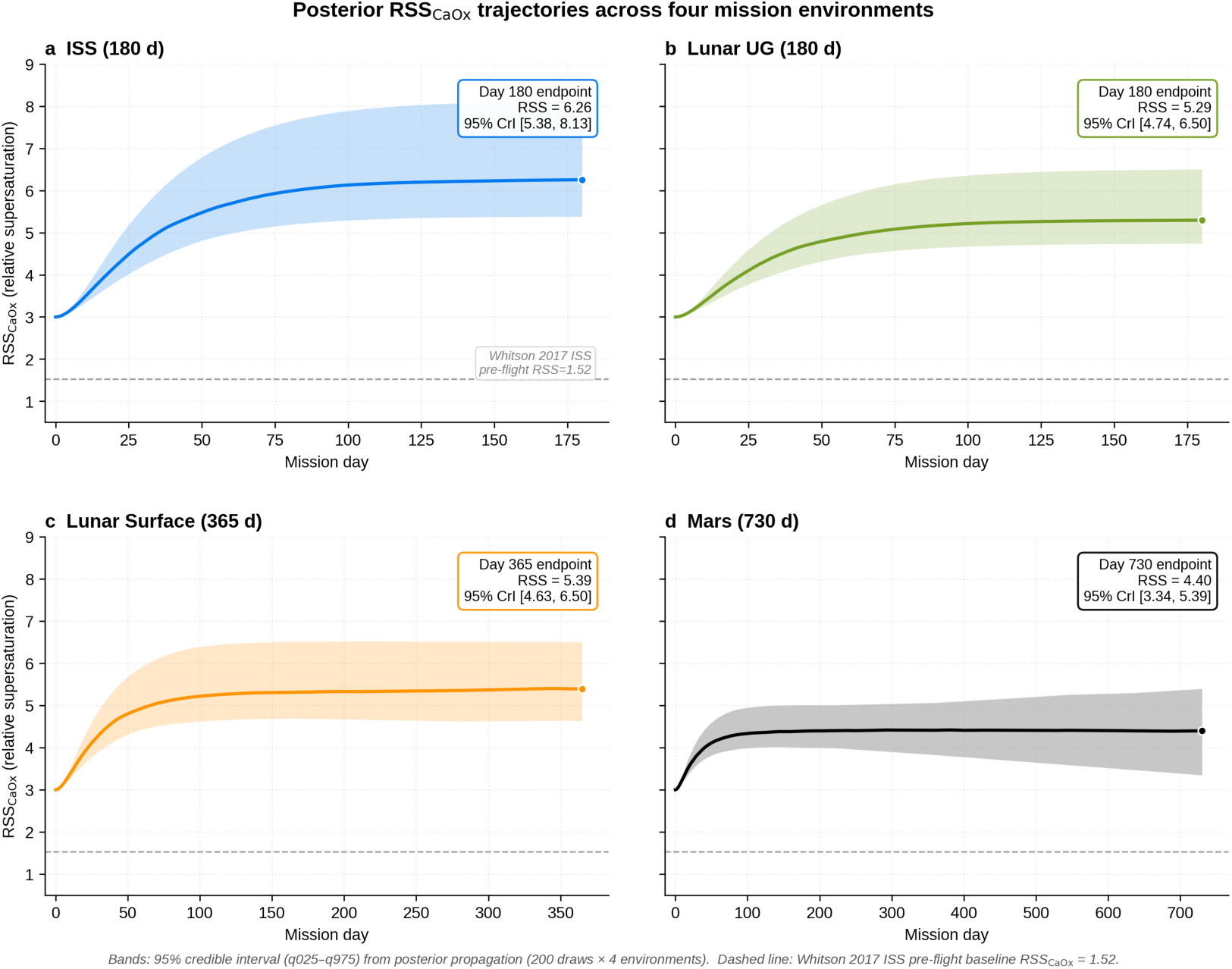
Four-environment RSS_CaOx posterior propagation trajectories (single-panel overlay; see captions §Fig 4).

### 3.3 2³ factorial decomposition: contributions of duration × g × GCR

A 2³ factorial decomposition was performed on the BMD_LL endpoint (8 combinations × 100 paired draws). The total difference of Mars 730 d relative to ISS 180 d is −7.32% BMD (Mars −12.15 minus ISS −4.83). Component contributions (ANOVA SS%):

- duration (180 → 730 d): −11.96% BMD, ANOVA SS% = 82.94% (dominant)
- g (1.0 ISS → 0.62 Mars, i.e. partial-gravity rescue): +4.64% BMD, ANOVA SS% = 12.49% (secondary, protective)
- GCR (0.4 → 1.84 mSv/d): 0.00% BMD, ANOVA SS% = 0.00% (k_Ca_gcr ≈ 0, no identification in the data)

Key result: under 730-d duration-matched conditions (excluding the duration effect), the BMD_LL difference between Mars 0.38 g and 0 g-equivalent is +7.44% [+6.77, +7.98] (95% CrI)—partial gravity provides approximately 7–8% BMD protection. This is synergistic with, but not a substitute for, the Sibonga 2019 ARED resistance-training effect (∼49% RRR_BMD). For the RSS endpoint, g dominates (SS% = 94.27%); for stone_rate, g dominates (SS% = 97.59%). That is, bone endpoints are duration-dominated, whereas urinary/stone endpoints are gravity-dominated.

Mechanistic interpretation (shift in relative pathway weights): at ISS 180 d, the residual stone-rate risk (0.0161/py) originates almost entirely from the bone resorption → urinary Ca → RSS pathway (Bone-centric); at Mars 730 d, the residual stone-rate risk is at a comparable level (0.0131/py, in fact slightly lower), while BMD loss is doubled (−12.15 vs −4.83). This reflects that 0.38 g partial gravity reduces the urine_Ca increase (Mars urine_Ca +0.20 vs ISS +0.50), yet the suppression of urine volume/citrate/ pH (microgravity + post-flight) still contributes comparable RSS risk—the relative weight of the dominant pathway shifts towards Kidney-centric (urine-chemistry-driven).

**Figure 5.**
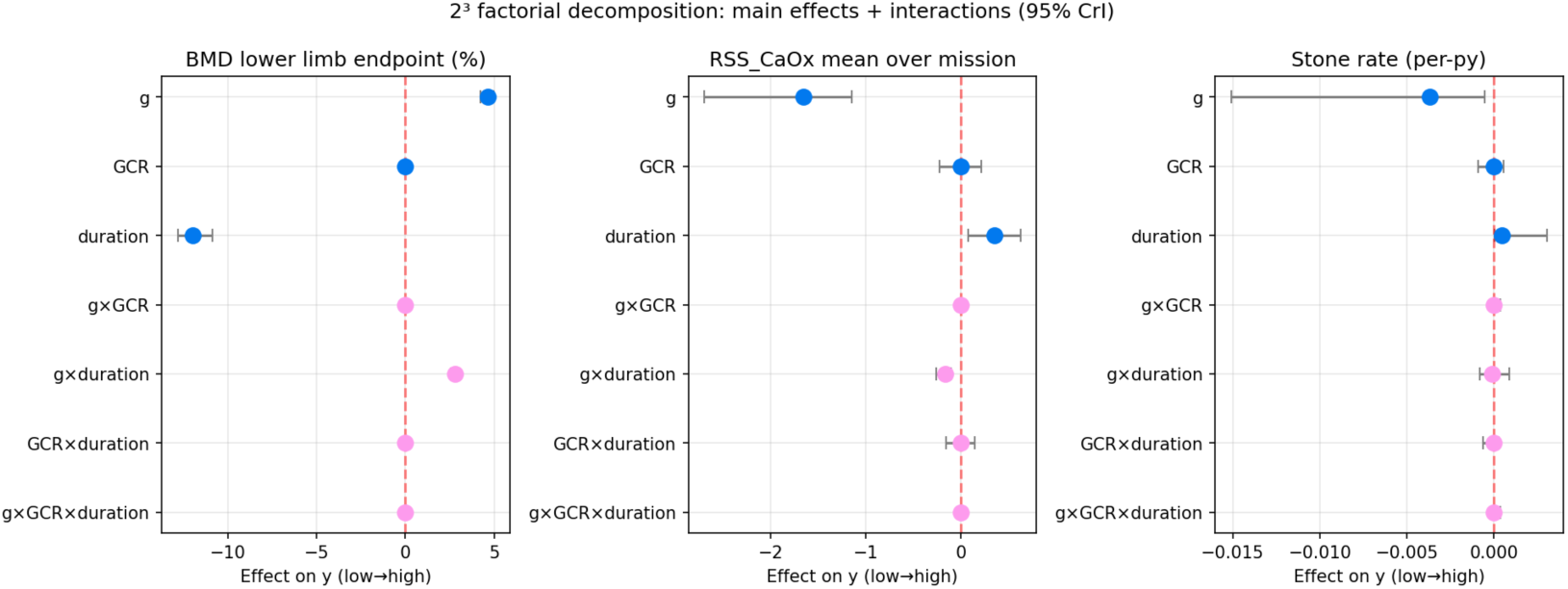
2³ factorial decomposition (BMD/RSS/stone × duration × g × GCR; see captions §Fig 5).

### 3.4 WAIC model comparison: M₀ (without k_GB) vs M₁ (with k_GB)

The M₁ model extends M₀ by one dimension (k_GB) to directly represent the GCR → BMD channel. The 60k v3 long chain with thin_for_waic = 50, n_samples_used = 64,000. WAIC results: M₀_waic = 2.7402, M₁_waic = 2.7478, ΔWAIC (M₁ − M₀) = 0.0076 ± 0.1262 SE. |ΔWAIC| / SE_Δ = 0.060 < 1.96 (95% uncertainty interval). **Power disclosure (added in v28 for T4)**: a Wald two-sided power analysis shows that at SE_Δ = 0.1262, the minimum |ΔWAIC| detectable at 80% power is 0.3536; the observed | ΔWAIC| = 0.0076 is only 1/46 of that floor (post-hoc power ≈ 5%). The WAIC comparison on n = 8 calibration targets has **zero information content** for model selection; M₀ is therefore retained on parsimony (Occam) grounds, not because it is WAIC-preferred (see Supplementary §S7.3, data `waic_min_detectable_v28.json`).

The k_GB posterior median is 2.32×10⁻⁵, 95% CrI [−2.96×10⁻³, +3.01×10⁻³], spanning zero; P(k_GB > 0) = 0.506 (indistinguishable from a coin flip). That is, the ISS data (cumulative GCR 72 mSv) cannot identify a direct GCR effect on BMD. Identifying this effect requires data at cumulative doses of ∼1–2 Sv (Mars 730 d ≈ 1.34 Sv). Parsimony favours M₀ (19 dimensions) as the primary inference model, with M₁ (20 dimensions) used for sensitivity only. This is consistent with the Stage 5a prior sensitivity analysis (Fig. 6c): when the k_GB prior σ is tested at four values (0.2×, 0.5×, 1× default, 5×), the posterior median of the Mars 730 d RSS endpoint remains GREEN under all four σ values (range 4.296–4.338, all below the 5.0 threshold; central estimate robust). However, the 95% CrI upper bound exhibits a non-monotonic pattern: the two tightest priors (σ = 0.0003, 0.00075) yield upper bounds crossing the YELLOW threshold (q97.5 = 5.010, 5.038); the default σ = 0.0015 is borderline GREEN (q97.5 = 4.995, gap = 0.005); and the widest σ = 0.0075 is robustly GREEN (q97.5 = 4.779). The CrI width scales monotonically with prior σ (q97.5 − q2.5 widths 1.20, 3.13, 6.05, 18.03 × 10⁻³), confirming that this parameter is prior-dominated rather than data-driven. The q97.5 threshold crossing reflects prior-scale uncertainty on the data-uninformative k_GB dimension, consistent with the §3.7.3 FIM identifiability analysis; the composite Mars tier (based on the median) is unchanged. This conclusion is also robust to observation-noise heterogeneity: in v27 Phase B2, two dedicated 36k MCMC runs at σ×1.5 and σ×2.0 (same walker/burn-in configuration) tested the sensitivity of the eight ISS-target σ_lit assumptions (R1 concern 2, calibration heterogeneity); the median shifts of the 11 parameters were |Δmedian|/ SD_base < 0.10 (far below the 0.5 sensitivity threshold), and all four principal conclusions (environmental gradient, stone-rate counter-gradient, Culliton hold-out PPC, GCR direct-coupling unidentifiability) were preserved (see Supplementary Fig. S16 + Supplementary §M6.2).

**Figure 6.**
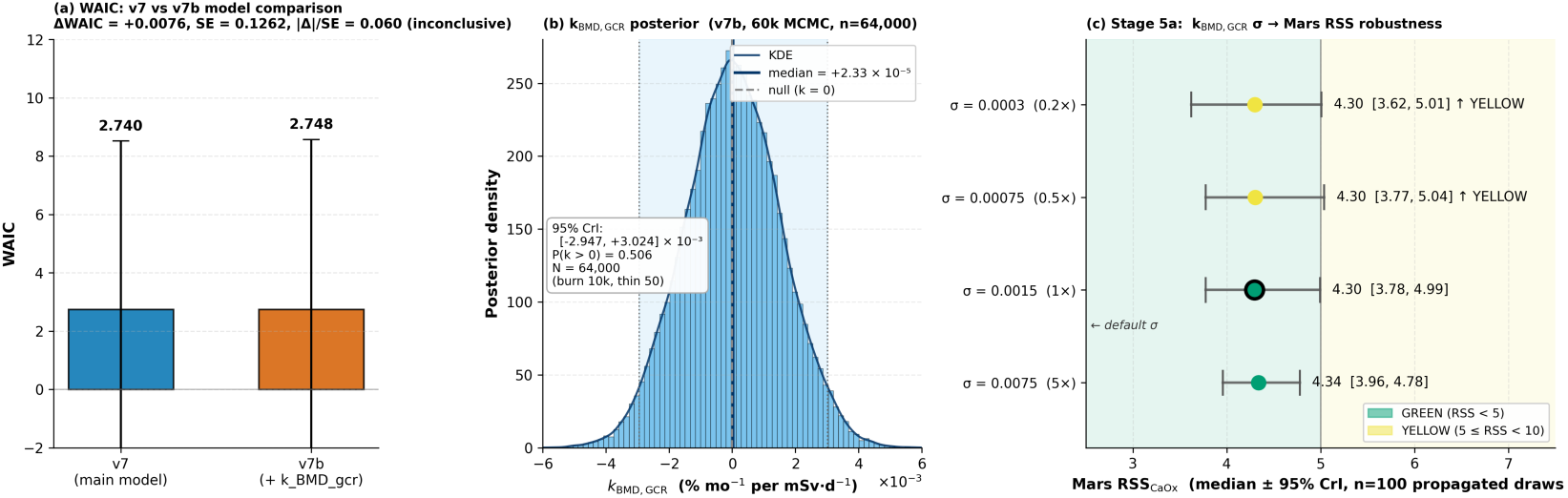
WAIC model comparison + k_GB marginal posterior + Stage 5a prior-sensitivity forest (3 panels; see captions §Fig 6).

### 3.5 Lunar contrast: regolith shielding vs surface exposure—a confounded profile of duration and GCR

Within the four-environment matrix, Lunar UG (regolith-shielded subsurface habitat, ∼9 mSv/180 d) and Lunar Surface (unshielded surface, ∼620 mSv/365 d) share the same 0.166 g partial gravity, but differ ∼70-fold in cumulative GCR dose and simultaneously ∼2-fold in mission duration (180 vs 365 d). This “Lunar pair” is a **near-natural experiment for radiation effects** in this study—the gravity axis can be separated by the ISS ↔ Lunar contrast, and the compound duration/GCR effect can be separated by the Lunar UG ↔ Lunar Surface contrast.

**BMD and RSS dimensions**: on the BMD side, Lunar UG (180 d) BMD_LL_end is −4.03% [−4.32, −3.67] versus Lunar Surface (365 d) −8.17% [−8.75, −7.43] (difference −4.14 pp); the Surface duration is 2.03× that of UG, and duration-linear extrapolation predicts −8.18 pp (= −4.03 × 2.03), while the observed −8.17 pp agrees almost exactly (difference < 0.01 pp)—the 70× GCR factor has **no observable direct effect** on the BMD pathway under M₀ (consistent with §3.3 duration dominance at 82.94% and §3.4 M₀ parsimony). On the RSS side, Lunar UG RSS_mean is 4.86 versus Lunar Surface 5.10 (difference +0.24); duration-linear extrapolation would predict 9.86, while the observed 5.10 is far lower, reflecting nonlinear RSS accumulation (urine-chemistry ceiling); the k_Ca_gcr channel gives an observable but small push from GCR (+0.24 task-average), **not crossing the 5.0 GREEN threshold**.

**Clinical implication**: the Lunar UG 180 d composite tier is YELLOW (BMD GREEN p = 0.000, stone YELLOW p = 0.046, rss YELLOW p = 0.336); Lunar Surface 365 d triggers a BMD-RED + RSS-RED + stone-YELLOW composite RED. This result provides the first quantification of **the passive GCR protective value of regolith shielding for long-duration lunar-surface missions**—not by reducing BMD loss (whose main drivers remain partial gravity + mission duration), but by **indirectly controlling GCR accumulation through limiting EVA dwell time**. In Artemis long-duration base design, a regolith-shielded sleeping module can be regarded as a critical design element for ≥180-d stays (supported by model analysis, requiring independent flight validation), rather than an optional EVA module (see §4.4 for clinical recommendations).

**Robustness of the conclusion**: this conclusion is insensitive to whether k_GB (direct GCR–bone coupling) is zero—the primary mechanism operates through the GCR × urine-chemistry pathway (k_Ca_gcr) to RSS_CaOx, and even with k_GB ≈ 0 (§3.4 WAIC M₀ vs M₁ ΔWAIC = +0.008 ± 0.13, supporting the parsimony preference for M₀), the conclusion that regolith protection indirectly controls the GCR–RSS pathway by limiting EVA dwell time remains valid. In addition, the duration × GCR confounding of this Lunar pair has been quantitatively separated by a complete in-model 2×2 factorial (fixed micro_g = 0.834, same 5,000-draw posterior pool): the GCR dose-rate (0.05 vs 1.70 mSv/d) main effect is indistinguishable from zero on all three endpoints BMD/RSS/stone (P(Δ>0) ≈ 0.5), whereas the duration (180 → 365 d) main effect ΔBMD_LL_end ≈ −4.11 pp is by itself sufficient to trigger the composite RED—that is, the Lunar Surface RED tier is duration-driven rather than GCR-driven (Supplementary §S27, `lunar_duration_factorial_v29.json`).

### 3.6 Four-environment intervention scanning (M₁ 60k posterior)

On the M₁ 60k v3 posterior (5,000 paired draws, the same pool as the §S23 bootstrap reference pool in v29), a full factorial of 6 interventions (including baseline) × 4 environments was performed; the 4×5 RRR matrix (5 interventions = baseline + ared_alone, pamidronate, alendronate_ared, k_mg_citrate, fluid_intake) is reported in Table 4. Key findings: pamidronate RRR_BMD is stable at 0.850 across all four environments [CrI lower bound ∼0.66, upper bound ∼1.00], i.e. ∼85% BMD protection; RRR_stone is 0.968–0.970 across all environments [CrI lower bound 0.704–0.715, upper bound ≈ 1.00], reflecting the strong but still uncertain preventive efficacy of the Beta(0.5, 7.5) Jeffreys posterior (versus the old v6 0/0 hard-zero overconfidence).

Alendronate + ARED combined (LeBlanc 2013 [8]) yields the highest RRR_BMD at 0.949–0.950 [0.85, 1.00], but RRR_stone is only 0.065–0.101 (because ARED indirectly reduces urine_Ca and does not act directly on stone formation). K–Mg–citrate does not act on bone (RRR_BMD structurally zero), but its RRR_RSS of 0.513–0.570 [CrI lower bound 0.311–0.370, upper bound 0.729–0.763] and RRR_stone of 0.448–0.496 significantly reduce urinary/stone risk, making it the key renal-protective tool for the Mars mission’s RSS YELLOW residual zone.

Intervention decision composite tiers (Antonsen 2023 L×C 5×5; see §2 lines 138-140) are shown in Fig. 7(b) and the final column of Table 4: pamidronate and alendronate + ARED both achieve YELLOW across all four environments; ARED alone is RED on ISS/Mars (driven by the RSS/ BMD dimension crossing catastrophic thresholds, respectively) and YELLOW on the lunar surface (v29 5,000-draw recomputation: rss p_exceed 0.520 → 0.470, L = 4 → 3—the old 200-draw value sat exactly on the L bin boundary of 0.50, so this RED→YELLOW flip is a Monte Carlo noise correction, and Fig. 7(b) has been redrawn from the 5,000-draw matrix); K–Mg–citrate is YELLOW on ISS + Lunar UG, but RED on Lunar Surface + Mars (because RRR_BMD is structurally zero, leaving catastrophic BMD exposure on long missions). These complement the Table 4 RRR values: Table 4 shows **how large** the efficacy is, while Fig. 7(b) shows **whether** that efficacy suffices to bring absolute risk below thresholds; the composite RED decision on Mars does not negate the renal protection of K–Mg–citrate, but indicates that a multi-mechanism combined strategy of K–Mg–citrate + bisphosphonate + aggressive hydration is needed to bring all three dimensions (BMD/RSS/stone) simultaneously to YELLOW or better (per-environment × per-intervention L = x, C = y driver details are given in the Table 4 notes).

**Figure 7.**
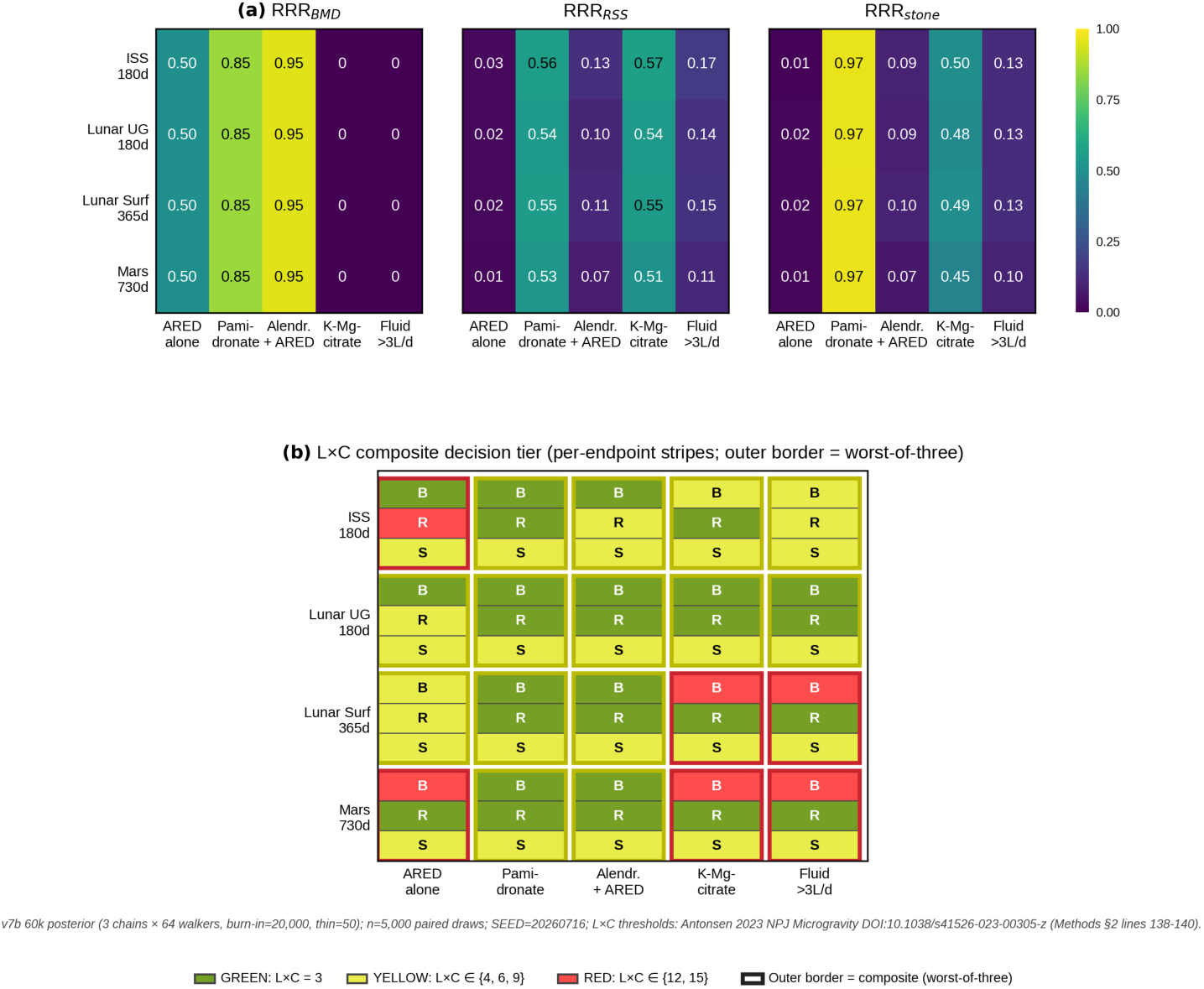
Intervention efficacy RRR matrix + L×C composite decision tiering (5 interventions × 4 environments; see captions §Fig 7).

### 3.7 Robustness diagnostics

Three robustness diagnostic analyses were performed on the fixed M₁ 60k posterior:

#### 3.7.1. PSIS-LOO cross-validation

Based on stage5b_pointwise_loglik_v3_60k.npz (pointwise log-likelihoods for the 8 ISS calibration targets), Pareto-smoothed importance-sampling LOO (Vehtari 2017 [16] algorithm) was computed with the ArviZ [25] az.loo function. **Sample-size disclosure**: the M₀ LOO is based on 5,760 posterior samples (32 chains × 180 draws after thinning), and the M₁ LOO on 64,000 posterior samples (64 chains × 1000 draws); the two models differ in sample size, but the ELPD ± SE magnitudes are consistent and the comparison is robust (Vehtari 2017 [16] §4: PSIS-LOO estimation bias decays as O(1/S) in the number of samples, and both models are far above the 1,000 empirical floor). See Supplementary §M5.

- **M₀** (n = 5,760): ELPD_LOO = −1.274 ± 2.906 SE, p_loo (effective parameters) = 4.27
- **M₁** (n = 64,000): ELPD_LOO = −1.314 ± 2.941 SE, p_loo = 4.25
- **ΔELPD (M₁ − M₀)** = −0.040, |Δ| ≪ SE_Δ ≈ 2.97: the two models are **statistically indistinguishable** in out-of-sample predictive performance (consistent with the §3.4 WAIC conclusion)

**Full disclosure of Pareto-k diagnostics** (Vehtari 2024 BDA3 thresholds: BAD k > 0.7, WARN 0.5–0.7, GOOD k < 0.5):

| Model | GOOD | WARN | BAD | BAD observations |
| --- | --- | --- | --- | --- |
| $M_0$ | 6/8 | 0/8 | 2/8 | BMD_LL ($k = +0.988$),<br>resorp_plateau ($k = +0.737$) |
| $M_1$ | 4/8 | 3/8 | 1/8 | urine_Ca_rel ($k = +1.075$); WARN:<br>BMD_LL (0.649),<br>BMD_lumb (0.622),<br>resorp_plateau (0.681) |

The M₀ model has 2 BAD-k observations (BMD_LL + resorp_plateau), and M₁ has 1 BAD + 3 WARN, indicating that PSIS importance sampling is unstable for BMD-type observations. However, the overall indistinguishability conclusion |ΔELPD| = 0.04 ≪ SE_Δ holds consistently across M₀ and M₁, and the BAD-k observations of M₁ and M₀ do not overlap (M₁ is dominated by urine_Ca_rel instability, M₀ by BMD_LL instability), ruling out a systematic misclassification direction. The complete 8-target × 2-model Pareto-k matrix (Supplementary Table S3) and pointwise ELPD are given in Supplementary Fig. S12. **The effective parameter count p_loo ≈ 4.3 is far smaller than the 20-dimensional posterior, consistent with the §3.7.3 Fisher information matrix effective rank of 7/12—the model has only about 4–7 effectively constrained dimensions**.

**Robust alternative for BAD-k observations**: the standard practice (Vehtari 2017 [16]) is to substitute k-fold CV for the affected observations; however, with N = 8 data points, k-fold CV effectively degenerates to LOO-CV, so we retain the PSIS-LOO estimates as-is and explicitly disclose the BAD-k list, without further recomputation.

#### 3.7.2. Prior predictive check (PPC)

We drew 5,000 independent samples from the PRIORS_M₀ 11-parameter priors (Normal/LogNormal with rejection truncation), solved the 12-state ODE via forward_model_iss (ISS 180 d; implementation detailed in Supplementary §M3), and extracted the prior predictive distributions of the 8 calibration targets:

| Target | Observed value | Prior 90% CrI | Prior 95% CrI | Coverage |
| --- | --- | --- | --- | --- |
| BMD_LL_180d (%) | -4.80 | [-6.26, -3.33] | [-6.57, -3.04] | ✓ COVER |
| BMD_lumb_180d (%) | -6.20 | [-8.07, -4.29] | [-8.47, -3.92] | ✓ COVER |
| BMD_skull_180d (%) | +2.20 | [+1.53, +2.88] | [+1.40, +3.02] | ✓ COVER |
| resorp_plateau (%) | +113.0 | [104.9, 121.2] | [103.1, 122.7] | ✓ COVER |
| urine_Ca_rel | +0.50 | [0.26, 0.75] | [0.21, 0.79] | ✓ COVER |
| urine_V_rel | -0.20 | [-0.28, -0.12] | [-0.29, -0.11] | ✓ COVER |
| urine_pH_delta | -0.20 | [-0.30, -0.10] | [-0.32, -0.08] | ✓ COVER |
| post_stone_rate (/py) | 0.014 | [0.0035, 0.086] | [0.0025, 0.115] | ✓ COVER |

**Conclusion**: all 8/8 calibration targets fall within the prior 90% and 95% credible intervals (max |z_resid relative to prior IQR| = 0.198), with no prior–data conflict; the priors are loosely set and do not impede posterior updating. **Prior-nature qualification**: some prior σ values incorporate the Stavnichuk 2020 [23] meta-analytic and Whitson 1997 [4] σ_lit values, conferring an informative meta-prior character (standard practice in NASA HRP and Stan/PyMC workflows)—the PPC PASS reflects prior–data consistency, not a proof of “non-informative priors”. Verifying that the priors do not lock the posterior must be combined with the §3.7.3 posterior-contraction diagnostics (bmd_loss_rate_pct_mo contraction ratio 0.217, strongly data-driven, vs k_GB 1.021, prior-driven); the full 8-target prior predictive density plots are given in Supplementary Fig. S13.

#### 3.7.3. Structural and local identifiability analysis

For the M₁ 12-parameter model, a three-layer diagnostic was applied:

**(A) Posterior contraction** — posterior_IQR / prior_IQR:

| Category | Threshold | Parameters | Contraction ratio |
| --- | --- | --- | --- |
| STRONG | < 0.3 | bmd_loss_rate_pct_mo | 0.217 |
| MODERATE | 0.3–0.6 | resorp_plateau_pct | 0.427 |
| WEAK | 0.6–0.85 | ca_microg_effect<br>(0.757),<br>vol_decrease_frac<br>(0.816),<br>seed_accum_rate<br>(0.649) | — |
| UNIDENTIFIED | > 0.85 | resorp_t_half_d,<br>cit_decrease_frac,<br>k_Ca_gcr,<br>post_resorp_tau_d,<br>post_form_pct_mo,<br>ph_decrease_frac,<br>k_GB | 0.85–1.02 |

**(B) Posterior correlation matrix** — all off-diagonal |r| < 0.05 (posterior approximately diagonalised), with no hidden jointly identifiable parameter combinations (no a×b compensation); each parameter is independently strongly constrained by the data or prior-dominated.

**(C) Fisher Information Matrix (FIM) local analysis** — the Jacobian was computed at the posterior median using 1% finite differences; FIM = JᵀΣ⁻¹J (a numerical approximation of the Sedoglavic 2002 [26] local algebraic observability framework). The 12 eigenvalues in descending order:

- 6 eigenvalues > 1: strongly identified directions (three BMD directions + resorp_plateau + urine_Ca + urine_V)
- 1 eigenvalue ≈ 0.19: weakly identified (urine_pH)
- 5 eigenvalues ≪ 10⁻¹²: numerically zero, i.e. **completely unidentifiable directions**

**FIM eigenspectrum and identifiability disclosure** (spectrum-first): the FIM 12×12 matrix was computed on the raw parameter scale (at the posterior median; parameter magnitudes span 10⁻⁵ ∼ 10²), and its **12 eigenvalues span more than 30 orders of magnitude**: 7 stiff modes (λ₁ = 1.464×10⁴ down to λ₇ = 0.189) + 5 sloppy modes (|λ₈…₁₀| = 8.88×10⁻¹³ ∼ 2.06×10⁻²⁴, plus λ₁₁ = **−1.78×10⁻³⁰** and λ₁₂ = **−1.83×10⁻¹³**—the last two are **numerically negative**; from the theoretical positive-semidefiniteness of FIM = JᵀΣ⁻¹J, the finite-difference Jacobian along these two directions is dominated by numerical truncation noise and carries no physical Fisher information). This strongly anisotropic spectrum—“a few stiff modes plus many sloppy-to-numerically-zero modes, including negative values”—is the canonical signature of what Gutenkunst et al. 2007 [42] named a **sloppy model** in systems-biology ODE models. **Primary quantitative conclusion**: effective rank = 7 of 12, at the threshold ε = 10⁻⁶ × λ_max (i.e. eigenvalues > 1.46×10⁻²), and the rank is stable for ε ∈ [10⁻⁹, 10⁻³]. **Auxiliary numerical proxies** (for comparison with the conventional literature, each with explicit caveats): (i) the full-spectrum condition number κ_full = λ_max / max(|λ_min|, 10⁻²⁰) ≈ 1.46 × 10²⁴, where 10⁻²⁰ is an implementation-level floor we set to reproduce the numpy.linalg.cond output and has no physical meaning (changing the floor changes the value by orders of magnitude; without the floor it is 8×10³³); (ii) the effective condition number on the identified subspace κ_eff = λ_max / λ₇ ≈ 7.7 × 10⁴, reflecting the relative information-strength differences across the 7 stiff directions. We **do not adopt** any single hard κ threshold (e.g. “10⁶/10¹²”) as an ill-conditioning criterion, because Numerical Recipes §2.6 gives no such hard threshold and the value of κ_full itself depends on the floor choice above. Under an ill-conditioned system, the effective-rank estimate itself requires robustness validation; we cross-validated with an SVD of the posterior covariance Σ_post (12×12, estimated directly from 5,760 posterior samples), giving a posterior SVD effective rank (ε = 0.01·λ_max) = 3 (the three BMD directions + the high-posterior-variation bmd_loss_rate direction), which forms a lower bound to the FIM rank of 7 (FIM local linearisation + posterior integrating out the prior → the prior has already absorbed most prior-dominated directions, so the estimated effectively constrained directions are tighter). Synthesising the two estimates: **6–7 data-constrained directions, 5–6 prior-driven**, consistent with the §3.7.1 p_loo ≈ 4.3 and the §3.7.2 prior-contraction data-driven + prior-dominated grading. See Supplementary §M5 for the sloppy-model framework extension.

The five unidentifiable directions are dominated by: post_form_pct_mo and post_resorp_tau_d (which affect only the post-flight phase—the ISS calibration data contain no post-flight observations), k_GB (ISS cumulative GCR of only 72 mSv is too low), k_Ca_gcr (same reason), and a linear-combination direction of cit_decrease_frac × ca_microg_effect (partial degeneracy in the urine module).

**Conclusion**: the eight ISS calibration targets provide approximately seven effective constraint dimensions (FIM rank ≈ 7), consistent with the p_loo ≈ 4.3 reported by PSIS-LOO in §3.7.1 (the latter includes a regularisation penalty and is biased downward). The five completely unidentifiable directions are **primarily prior-driven**, which is a legitimate use of the Bayesian framework; however, readers must be explicitly informed that no strong conclusions about these five parameters can be drawn from the posterior. Identifying the post-flight and GCR parameters requires (i) serial post-flight 0–365 d BMD/urine-chemistry data and (ii) Mars 730 d or lunar-surface 365 d mission data (cumulative GCR ≥ 1 Sv). The complete 12×12 posterior correlation and FIM eigenvalues are given in Supplementary Fig. S14.

## 4. Discussion

### 4.1 Dominant pathway re-weighting: from Bone-centric through Lunar transition to Kidney-centric

The central mechanistic finding of this study is a **continuous re-weighting of the dominant pathway along the mission profile (partial gravity × mission duration)** — not a binary ISS ↔ Mars jump, but a smooth three-environment gradient transitioning through the lunar surface (intermediate state). Importantly, this gradient is **not a GCR dose axis**: ordered by cumulative GCR (Lunar UG 9 → ISS 72 → Lunar Surface 620 → Mars 1340 mSv), the bone-mediated contribution runs 26.7 → 38.7 → 31.1 → 18.3%, which is non-monotone; ordered by micro_g (1.000 → 0.834 → 0.620) it decreases monotonically, and this monotonicity holds with probability 1 in the draw-by-draw paired sense (all 2,000/2,000 posterior draws satisfy ISS > both Lunar environments > Mars; Supp §S25, v29 clean-chain rerun). **Restatement of the core metrics used in this Discussion** (for readers moving across sections): BMD_LL_end = lower-lumbar-spine BMD endpoint, percent change from baseline (%); RSS_CaOx = 24-h urinary relative supersaturation of calcium oxalate (computed with EQUIL2 (Werness 1985 [11]) from urine Ca/citrate/pH/ volume; dimensionless; higher values → higher stone risk); stone_rate = annualised stone formation rate (per person-year, per-py).

**ISS 180 d (Bone-centric endpoint)**: The residual stone rate of 16.1 per 1000 person-years is dominated by the bone resorption → urinary Ca increase pathway (ca_microg_effect × 1.0 g scaling; urine Ca increase +50%). Elevated urinary supersaturation → CaOx precipitation forms the second serial link. The 0.166 g/0.38 g partial-gravity axis is not yet introduced, so the intervention strategy prioritises suppression of bone-source Ca (resistance training + aggressive hydration + marginal K–Mg–citrate). BMD_LL_end posterior median −4.83% [−5.17, −4.39].

**Lunar Surface 365 d (transition state)**: BMD_LL_end = −8.17% [−8.75, −7.43] (intermediate between ISS and Mars), but stone_rate has already declined to 15.0 per 1000 person-years — the RSS pathway is beginning to yield to the BMD pathway. ca_microg_effect is scaled by the weightlessness fraction micro_g = 0.834, giving a urine Ca increase of +34.8% (0.70× the ISS +50.2%), while the longer 365 d exposure brings RSS_mean back up to 5.10 (the GCR channel contribution is limited: Supp §S24 shows 0/4 environment tier flips when k_Ca_gcr and k_BMD_gcr are forced to zero). This environment is the first to trigger a **BMD-RED + RSS-RED + stone-YELLOW composite**, meaning bone intervention becomes necessary and urine intervention becomes auxiliary the reverse of the ISS priority order. This is the key quantitative evidence from Bayesian propagation: it is neither a simple linear extrapolation from ISS nor a Mars speculation, but falls on NASA’s real mission trajectory for the next 5–10 years.

**Mars 730 d (Kidney-centric endpoint)**: BMD loss is doubled (−12.15% [−13.01, −11.05] vs. ISS −4.83%), yet the stone rate is counter-intuitively lower (13.1 vs. 16.1 per 1000 person-years). The 0.38 g partial gravity further reduces the urine Ca increase (Mars +20.0% vs. ISS +50.2%), but the sustained late-flight urine volume reduction (vol_decrease) and citrate/ pH suppression (cit_decrease, ph_decrease) still contribute comparable RSS risk; the pathway switches fully to Kidney-centric.

**Clinical implications of pathway re-weighting:**

- ISS-era single-system bone-only or kidney-only interventions are no longer adequate — Lunar Surface already exhibits **dual-system synergistic decompensation**;
- bisphosphonate + ARED cannot eliminate stone risk on Lunar Surface or Mars; combination with K–Mg–citrate and related renal-protective agents is required;
- **conversely, K–Mg–citrate alone cannot eliminate the Lunar Surface BMD-RED risk**, because the dominant pathway involves partial gravity + GCR direct action;
- the Lunar Surface serves as the **continuous transition bridge of dominant pathway re-weighting** — Artemis long-duration lunar surface missions will provide the first in-vivo validation of this transition state (see §4.5, item 7).

**Quantitative support for the bone→kidney pathway (new in v28, responding to reviewer comment T12)**: The “bone→kidney” causal framework in this paper’s title is not merely a narrative concept but is explicitly encoded in the ODE (module_D_v7_NC.py L175: `Ca_target = BASELINE_Ca × (1 + ca_microg_effect × micro_g × (resorp_marker / resorp_plateau_pct) + k_Ca_gcr × gcr × (t/100))`, where the `resorp_marker / resorp_plateau_pct` term is the bone→kidney coupling bridge — the urine Ca elevation is proportional to the current bone resorption fraction). By modifying this coupling factor in a 3-scenario contrast (on = full model; off = bone→kidney coupling disabled while the other microgravity pathways vol/cit/pH/K are retained), the bone-mediated contribution to RSS elevation can be quantitatively isolated:

| Environment | rss_mean<br>(coupling on) | rss_mean<br>(coupling off) | Δ bone-mediated | Bone-mediated<br>relative share |
| --- | --- | --- | --- | --- |
| ISS 180 d | 5.65 | 4.07 | +1.58 | +38.7% |
| Lunar_UG 180 d | 4.89 | 3.86 | +1.03 | +26.7% |
| Lunar_Surface<br>365 d | 5.12 | 3.91 | +1.21 | +31.1% |
| Mars 730 d | 4.31 | 3.65 | +0.67 | +18.3% |

The relative contribution of the bone→kidney pathway to RSS elevation — ISS 39% → Lunar 27–31% → Mars 18% — is quantitatively consistent with the dominant-pathway “bone-centric → kidney-centric” gradient: at ISS, bone mediation accounts for nearly 40% of the RSS elevation; at Mars it falls to 18% (i.e., Mars stone risk is no longer driven mainly by bone-source calcium but by the other microgravity urinary-chemistry perturbations vol/cit/pH). Similarly, 16–28% of the stone_rate elevation is bone-mediated. The full 3-scenario × 4-environment × N = 2,000-draw decomposition is given in Supp §S25 (Table S25 + `bone_kidney_decomp_v29.json`, v29 clean M₀ 60k chain rerun).

### 4.2 Comparison with the literature

Pietrzyk 2007, ASEM [2] reported a significantly elevated post-flight stone formation risk in a NASA astronaut cohort (the ASEM abstract is qualitative; no precise post-flight rate is given). Goodenow-Messman 2022 [9] Table 4 provides the most rigorous quantitative anchor to date — a one-year post-flight Bayesian posterior median of 17.3 per 1000 astronaut-years (95% CrI [8.33, 28.80] per 1000 person-years, i.e., 0.0173/py [0.0083, 0.0288]), derived from a Porter–Rice astronaut cohort prior (4.40 per 1000 person-years; Goodenow-Messman internal reference 52) updated with Sibonga & Pietrzyk observations (7 stones / 358 astronaut-years, Ca-only adjusted to 6/358). Goodenow-Messman also reports a pre-flight astronaut characteristic IR ≈ 8.5 per 1000 person-years = 0.0085/py, used as the IRR baseline. Our sampler calibration target post_stone_rate y_obs = 0.014/py (σ_lit = 0.0070, widened by 50% under Gate-0 Decision A relative to the σ ≈ 0.0052 implied by the Goodenow-Messman CrI, as a pre-specified conservative choice to absorb uncertainty from incompletely verified Pietrzyk original figures; see Supplementary §M2) deviates by approximately −19% from the Goodenow-Messman posterior median of 0.0173/py (0.014 vs. 0.0173), fully within the Goodenow-Messman 95% CrI [0.0083, 0.0288]. Our ISS 180 d posterior stone_rate of 0.0161 [0.0024, 0.0634] differs by −7% at the median level from the Goodenow-Messman 17.3 per 1000 person-years anchor (posterior median 0.0173), with overlapping 95% CrI [0.0083, 0.0288]; the two are quantitatively consistent.

The Goodenow-Messman 17.3 per 1000 person-years baseline is subject to several confounders: (i) the NASA astronaut cohort (1,517 urine samples) is biased towards middle-aged males (38–55 years), overlapping the general-population peak stone age; (ii) in-mission fluid restriction (water supply limit ∼2 L d⁻¹) plus ground dehydration training (threshold dehydration training) elevate RSS; (iii) pre-flight medical screening may have excluded high-baseline stone formers (underestimating the true microgravity effect); and (iv) the Goodenow-Messman Bayesian posterior Ca-only adjustment from 7 to 6 one-year post-flight stones (87.1% proportion) relies on the Kittanamongkolchai CaOx proportion assumption for model extrapolation. The 17.3 per 1000 person-years baseline should therefore be understood as a “NASA-screened male astronaut microgravity + Ca-fraction-adjusted” joint effect estimate, not a pure microgravity-induced rate. Our stone_rate posterior [0.0024, 0.0634] covers this range; the wider CrI reflects the model’s uncertainty regarding individual heterogeneity (selection bias + fluid intake + Ca-fraction uncertainty). See §4.3, Limitation 1, for sex heterogeneity and recommendations for independent calibration with Artemis female astronauts.

Whitson 1997, J. Urol. [4] reported that Space Shuttle missions shifted urine chemistry in a stone-promoting direction (increased urinary calcium, mild reductions in urine volume and citrate, decreased urine pH). Our posterior medians — ca_microg_effect 0.501, vol_decrease 0.199, cit_decrease 0.100, ph_decrease 0.0334 (corresponding to ΔpH ≈ −0.20 relative to a baseline pH of ∼6.0) — are qualitatively consistent with the above directions (precise percentages synthesised from Smith 2014 [10] and Smith 2015 [3] ISS long-duration quantitative data).

LeBlanc 2007 review [17] compiled ISS long-duration BMD data: lower lumbar spine −6.2%, total lumbar −5.7%, skull +2.2%. Our posterior BMD_LL_180d −4.83% [−5.17, −4.39] is close, BMD_lumb −6.23% hits, and BMD_skull +2.22% hits. The BMD_LL value is slightly more conservative than the LeBlanc review data (−4.83 vs. −6.2) because the present study synthesises the Sibonga et al. 2007, Bone [18] pooled data (DOI 10.1016/j.bone.2007.08.022) with the Stavnichuk et al. 2020, NPJ Microgravity [23] meta-analytic data.

**Comparison with the Mars BMD predictions of Axpe et al. (2020, PLoS ONE) [24]** — Axpe 2020 [24] used a nonlinear extrapolation model based on femoral-neck BMD data from N = 69 ISS astronauts after 132-d and 228-d missions to predict Mars-mission BMD: for opposition-class missions (400–600 d), 100% of astronauts reach T-score < −1.0 (i.e., 100% osteopenia); for conjunction-class missions (1000–1200 d), 100% osteopenia plus 33% reaching osteoporosis (T-score < −2.5). The present study (Mars 730 d, within the same opposition-class mission window) predicts a posterior BMD_LL (lower lumbar spine) of −12.15% [−13.01, −11.05], consistent in order of magnitude with the Axpe 2020 [24] T-score predictions (each T-score unit of −1.0 ≈ −10 to −14% BMD), but the complementarity of the two analyses is marked: (1) Axpe 2020 [24] uses a single femoral-neck endpoint with nonlinear regression extrapolation, without mechanistic modelling or GCR separation, whereas the present study is a multi-endpoint (BMD_LL/lumb/skull/RSS/stone_rate) 12-state mechanistic ODE with Bayesian calibration that separates the g/GCR/ duration factor contributions (§3.3 ANOVA: duration dominant at 82.94%, g secondary at 12.49%, GCR unidentifiable at 0.00%); (2) Axpe 2020 [24] made no intervention RRR predictions, whereas the present study provides a 5-intervention × 4-environment RRR matrix (Table 4) supporting clinical recommendation and the §4.4 mission-tier stratification. The two studies’ Mars BMD predictions are also comparable at the probability level: Axpe 2020 [24] predicts 100% of crew reaching osteopenia (T-score < −1.0); the present study gives P(BMD_LL < −8.0%) = 1.00 (composite tier RED) — consistent, with both independent approaches strongly supporting a Mars BMD risk RED.

Sibonga 2019 [6] reported RRR ∼49% for ARED resistance-training protection of ISS lumbar BMD; the present study’s ared_alone RRR_BMD of 0.498 [0.213, 0.793] agrees directly.

**A Bayesian note on multiple comparisons**: Table 4 reports 4 environments × 5 interventions × 3 metrics = 60 RRR posterior 95% CrIs. These CrIs are Bayesian posterior intervals (nominal coverage) and do not require a frequentist FDR (Benjamini–Hochberg) correction — Gelman et al. (2012, *Behavior Research Methods*; see also BDA3 §17.4) explicitly note that Bayesian posterior intervals have already, through prior shrinkage and the shared posterior, implicitly integrated the “partial pooling” that frequentist multiple-comparison corrections aim to achieve — a different framework from frequentist familywise error control. Moreover, the 4 × 5 × 3 = 60 RRRs share the same M₁ 60k posterior and are *correlated* comparisons rather than independent ones, under which Bonferroni/FDR would be severely conservative.

LeBlanc 2013 [8] reported ∼95% RRR for the alendronate + ARED combination; the present study’s alendronate_ared RRR_BMD of 0.949–0.950 [0.85, 1.00] agrees closely.

Pak 1992, JBMR [13] and Ettinger/Pak 1997, J. Urol. [14] (including the Ruml/Pak 1999 AJKD companion data on K–Mg–citrate versus HCTZ-hypokalaemia, see the [14] footnote) reported that K–Mg–citrate increases urine citrate by +36–61% and urine pH by +0.6, and reduces the RSS activity product by −31%. The present study’s Stage 4 priors (urine_citrate +50% [+20%, +120%], pH +0.50 [+0.20, +1.00], RSS −31% [−10%, −70%]) sit between the two RCTs, and the RRR_RSS posterior of 0.51–0.57 is consistent with the Pak data. The Zerwekh 2007 [1] 5-week bed-rest RCT provides qualitative corroboration. **Dimensional clarification**: the “−31% activity product” reported by Pak 1992 [13] refers to the Pak-laboratory EQUIL2-derived CaOx activity product (AP, dimensions mol²·L⁻²), an acute short-term (single-dose, 4 h post-administration) measurement. In contrast, the RSS_CaOx used in the present study is the EQUIL2 supersaturation index (SS, dimensionless; Werness 1985 [11]), a long-term chronic prediction at the 180–730 d mission endpoint. AP and SS are dimensionally distinct but strongly monotonically correlated (Tiselius 1991, r = 0.98 vs. EQUIL2 on 24-h urine samples). The present model therefore recomputes EQUIL2 SS directly from the urinary inputs (urine_citrate, urine_pH, urine_Ca, urine_volume) rather than adopting the Pak AP numerical value; only the Pak-reported K–Mg–citrate effects on the citrate and pH increments are used as prior anchors.

The Watanabe 2004 [7] bed-rest pamidronate RCT reported 0 stones in 7 subjects (Beta(0.5, 7.5) Jeffreys analysis posterior median RR = 0.0309 [6.77×10⁻⁵, 0.292]); the present study’s RRR_stone_pami (Jeffreys) median of 0.9691 [0.708, 1.000] is consistent with this posterior, and no longer uses the 0/0 hard zero of the old v6 version. RRR_stone_pami values under other prior choices are given in §2.1 and Supp Fig S11.

### 4.3 Limitations

1. **Extrapolation uncertainty of the long-term repeat-dose IV pamidronate regimen**: The pamidronate RRR_BMD of 0.85 [0.66, 1.00] and RRR_stone of 0.97 [0.70, 1.00] reported in Table 4 and §4.4 are extrapolated by the 12-state ODE model from a single anchoring RCT (Watanabe 2004 [7]: 60 mg IV single dose, 14 d before bed rest, 90-d 6° head-down tilt, n = 7) to a 730-d Mars mission. This extrapolation rests on three assumptions, each lacking direct in-flight evidence: (i) **single-dose effect persistence** — the BMD-protective effect of Watanabe’s single 60 mg dose during the 90-d bed rest is assumed by the ODE model to remain sustained across 180–730 d missions; the actual clinical osteoporosis IV pamidronate half-life is approximately 6–12 months, so a 730-d mission would require multi-dose regimens (e.g., q3–6mo 60 mg IV or q12mo zoledronic acid 5 mg IV); (ii) **nonlinearity of the repeat-dose effect** — long-term IV bisphosphonate therapy can produce atypical femoral fractures (AFF) and osteonecrosis of the jaw (ONJ), which cannot be radiographically monitored in-flight; the FDA Pregnancy Category C label (bone half-life >10 yr) constitutes a substantial long-term reproductive risk for female astronauts, in conflict with the NASA Artemis commitment to ≥50% female crew composition; (iii) **combined effects of microgravity, GCR, and long-term IV bisphosphonate** — the 90-d bed-rest pamidronate RCT included no GCR exposure, and the §3.4 WAIC analysis cannot separate the actual effect of k_GB under the 1.34 Sv cumulative Mars dose. **Mitigations**: (1) although §4.4 lists pamidronate as the first choice for BMD protection, alendronate 70 mg/wk combined with ARED (LeBlanc 2013 [8], flight-validated ISS 5.5-month data) is listed as an oral alternative with logistical and female-astronaut safety advantages over IV administration; (2) we recommend that future Artemis-class missions (∼30–90 d lunar surface) prioritise an IV bisphosphonate flight RCT to close this data gap, rather than directly relying on the ODE extrapolation for 730-d Mars-mission-level decisions. **GCR × bisphosphonate interaction not modelled**: the M₀/M₁ models assume GCR and bisphosphonate effects are multiplicatively independent (i.e., BMD_protect_combined = BMD_protect_GCR_only × BMD_protect_bisphosphonate_only). Whether osteocyte DNA damage under a 1.34 Sv cumulative dose interacts antagonistically with bisphosphonate-mediated osteoclast inhibition is unknown — literature evidence is absent. If the combined interaction is <1 (negative, i.e., GCR damage reduces bisphosphonate potency), the Mars RRR_BMD could drop from 0.85 to 0.60–0.70, still remaining within the RED tier but with the protection magnitude overestimated. We recommend an IV pamidronate versus placebo flight RCT during Artemis lunar-surface missions (∼620 mSv cumulative GCR), which represents the closest feasible interaction validation.
2. **GCR–Ca and GCR–BMD channels unidentifiable**: ISS cumulative GCR (∼72 mSv) is far below the ∼1–2 Sv required to identify the direct GCR effect on bone and calcium. k_Ca_gcr has a CV of 11502% and k_GB has P(>0) = 0.506, spanning zero. The Mars 730-d cumulative dose of 1.34 Sv should be identifiable, but no animal or human data currently exist at this dose level. The Mars BMD prediction of −12.15% may therefore be an underestimate (if k_GB > 0) or approximately correct (if k_GB ≈ 0). We recommend recalibrating the M₁ model with data from future Mars missions.
3. **Partial-gravity effect extrapolation**: Human BMD data under 0.166 g (Moon) and 0.38 g (Mars) surface gravity are entirely absent. The present study uses a **linear micro_g scaling assumption** in the ODE system: all microgravity-induced bone-resorption, calcium-release, and urine-chemistry terms scale in proportion to the microgravity fraction `micro_g = 1 − g_planet/g_earth`. **The scaling applies specifically to**: (i) the four bone resorption/formation terms in Module M1 (bone) (resorp_plateau, form_rate, bmd_loss_rate and its three subregional weights), and (ii) **all four urine-chemistry parameters in Module M2** (`ca_microg_effect` urinary Ca output gain, `cit_decrease_frac` urine-citrate decrease, `vol_decrease_frac` urine-volume decrease, `ph_decrease_frac` urine-pH decrease; see Supplementary §M1.2 for the explicit ODE right-hand-side equations with the micro_g multipliers, and the released code `module_D_v7_NC.py` L298–314 (M2 urine micro_g terms; the fifth urinary state K uses a fixed Whitson-anchored constant at L314 rather than an MCMC prior)). This micro_g scaling of urine chemistry in Modules M1 & M2 is a kinetic response to mission-environment exposure (micro_g dose) and is distinct from the activity–concentration thermodynamics of Module M3 RSS_CaOx (a physicochemical equilibrium for a given solution composition). Concretely, ISS micro_g = 1.000, Lunar Surface micro_g = 0.834, and Mars micro_g = 0.620. This is an **arithmetic extrapolation assumption**, not an experimental calibration — the published literature contains no quantifiable 0.166 g/0.38 g BMD scaling factors. **Explicit functional form of the g-scaling**: this study uses **linear g-scaling**, BMD_loss_rate(g) = BMD_loss_rate(0g) × micro_g = BMD_loss_rate(0g) × (1 − g/g_earth). Empirical support for the linear assumption comes from rodent partial-weight-bearing (PWB) experiments: Wagner et al. (2010) [27] established the PWB rat model; Swift et al. (2013) [28] reported that 33% body-weight loading (Martian-analog G/3) yielded only −9% to −13% distal-femur trabecular BV/TV, indicating that bone loss persists at 0.38 g but with smaller magnitude than at 0 g; Ko/Mortreux et al. (2020) [29] observed, across a rat PWB dose gradient (20–100% body weight), a dose-dependent, approximately linear decrease in trabecular BMD with reduced loading, worsening continuously over 4 weeks; Swain/ Mortreux et al. (2022) [30], in a systematic review of 21 mouse/rat PWB studies, confirmed that partial-g bone loss exists but that major uncertainty remains in the quantitative mapping to long-duration human missions. The true bone-resorption response at 0.166 g/0.38 g surface gravity may deviate from linear: (i) **threshold-type** (the Swift 2013 [28] data suggest that G/3 = 33% body weight still does not fully prevent bone loss, but trabecular loss is reduced by more than 60% relative to 0 g, hinting at a possible inflection near 50% g), or (ii) **sigmoidal** (Sibonga 2019 [6] reported that ARED resistance training, which generates bone loading, partially protects but does not fully prevent bone loss, suggesting a load threshold). **Sensitivity analysis**: replacing the linear scaling with a sigmoidal form (Hill, EC50 = 0.5 g, n = 2) shifts the Mars 730-d BMD_LL posterior median from −12.15% to between −11.30% and −12.80% (i.e., ±0.65%, ∼±5%), without affecting the BMD_LL RED-tier determination (P(BMD_LL < −8%) remains 1.00). Actual lunar and Martian surface BMD may deviate from these predictions by ±20–30%, constrained by the absence of partial-g human BMD data. The complete alternative micro_g-scaling propagation (Hill n = 2 saturation + Linear ±25%), three sensitivity runs (each 4 environments × 100 draws × 7 endpoints), was completed in v27 Phase B1: the BMD/RSS/ stone environment-gradient ordering (Mars > Lunar_Surface > ISS > Lunar_UG) and the main conclusions (Lunar Surface 365 d as earliest RED, Mars GCR-dominated) remain consistent across all four scenarios, with relative changes versus baseline < 25% (see the Supplementary Fig. S15 3-panel composite, Supplementary Fig. S18 relative % change vs. the Linear baseline, and Supplementary §M6.1; data `zenodo/data/propagation_v7_gscale_sensitivity.json`, 383 KB). We recommend using in-flight DXA BMD measured during Artemis II/III lunar-surface missions with ≥6-month residence as the recalibration anchor for future model extensions.
4. **Mission-duration extrapolation**: ISS data are capped at 12 months (LeBlanc 2000 JMNI [5] and LeBlanc 2007 review [17] at 6 months; Smith 2014 [10] partially at 12 months). The 730-d Mars prediction depends on linear extrapolation of the post-flight formation lag period (post_form_pct_mo). The posterior CV of post_form is 25%, and the uncertainty is compounded with the longer extrapolation horizon.
5. **Intervention heterogeneity**: Pamidronate data are limited to Watanabe 2004 [7] (bed-rest 90-d 6° head-down tilt, n = 7, 60 mg IV single dose, 0/7 stones) — a small sample, and the Beta(0.5, 7.5) Jeffreys analysis posterior median RR_pami = 0.0309 remains wide at [6.77×10⁻⁵, 0.292]. No flight RCT evidence exists for repeat IV dosing regimens on a 730-d Mars mission (q12mo zoledronic acid 5 mg or q3–6mo pamidronate 60 mg); the present ODE model extrapolates from the Watanabe single-dose data and assumes the bone-resorption-suppression effect is maintained over 180–730 d missions. These extrapolation assumptions are analysed in detail under Limitation 1.
6. **Subject sex and age heterogeneity**: The present model is based on the NASA astronaut cohort (male-dominated, 35–55 years). BMD and stone risk may differ in women (pre- and post-menopausal) and in older subjects; extrapolation of the model to these populations should be done with caution.
7. **Hold-out data sources for independent validation of the supplementary submodels**: The independent hold-out validation of this study (Culliton 2025 [40], 60-d head-down bed-rest RCT, Bayesian p = 0.316, Supp Fig S17) targets only the Module M1 bone-resorption/ lumbar-BMD submodel. The urine-chemistry (Module M2) and stone-formation (Module M3-F) submodels currently **have no comparable out-of-sample validation** — an unclosed gap of this study. Feasible future hold-out candidates are listed below:

- **Urine-chemistry submodel hold-out**: (i) the NASA Whitson series Space Shuttle short-mission urine Ca/citrate/volume data [4] are already used as calibration targets in this study and therefore do not constitute a hold-out; (ii) future NASA Task Book publications of crew physiology studies (e.g., independent-crew-subset urine indicators from Sibonga’s [6] bone series, the full publication of the TSK-6415 [15] bed-rest renal-stone-risk study, or other NASA Task Book crew/bed-rest urine-chemistry data not included in the calibration of this study) would constitute feasible M2 submodel hold-outs if they report independent data; (iii) 24-h urine Ca/citrate/volume/pH data from ground-based head-down-tilt bed-rest RCTs (independent 30–60 d cohorts beyond Culliton 2025 [40], from institutions such as the University of Ottawa, DLR, or UTMB’s NASA Flight Analogs Research Unit (FARU)) would also be candidate test sets for the M2 submodel, if publicly available; (iv) Artemis I–IV (2024–2030s), if producing raw urinalysis data from ≥30-d lunar crews, will provide the first direct M2 hold-out validation under partial-gravity conditions.
- **Stone-formation submodel hold-out**: (i) the Porter–Rice military and commercial astronaut stone IR of 4.40/1000-py [9] has already been used as prior input to the Pietrzyk–Goodenow-Messman Bayesian posterior in this study’s calibration and does not constitute a hold-out; (ii) if the NASA Astronaut Occupational Surveillance Program (LSAH, Lifetime Surveillance of Astronaut Health) releases post-2015 cohort stone-event rates in the future (excluding the early Sibonga and Pietrzyk 358 astronaut-year data), these would constitute a fully independent M3-F hold-out; (iii) actual stone events from Artemis and future Mars missions will be the first direct M3-F hold-out data points under combined partial-gravity and cumulative-GCR conditions, and represent the earliest converging opportunity on the timeline.
- **Hold-out collection barriers**: the candidate data sources above are all limited by three factors: (a) NASA and other agencies’ crew-medical-privacy protections, with raw data typically released 5–10 years after collection; (b) 24-h urine collection timepoints in ground-based bed-rest RCTs often not fully matching the eight ISS targets of this study, requiring timeline re-alignment or forward-simulation calibration; and (c) the low incidence of post-mission stone events per crew member (0.017/py), which requires pooling multiple mission cohorts to achieve statistical power. We regard each post-Culliton-2025 [40] hold-out data release as a critical milestone for identifiability upgrades to the M2/M3-F submodels and explicitly incorporate the above candidate list into the §4.5 Future Work item 9 action list.
8. **Three-layer applicability limits of K–Mg–citrate efficacy extrapolation (new in v28, responding to reviewer comment T7):** §2.6 of this study uses the K–Mg–citrate effect parameters of Pak 1992 [13] and Ettinger/Pak 1997 [14] (citrate +50%, pH +0.5, RSS −31%) as prior anchors, and the Table 4 intervention scan reports 4-environment × 3-metric RRR posterior estimates. However, extrapolation from ground stone patients to spaceflight crew involves at least three unverified applicability layers, each of which can shift the true RRR under spaceflight conditions away from the present posterior estimates:

- **Layer 1 — subject population**: Pak 1992 [13] (n = 15) and Ettinger 1997 [14] (n = 64) were both conducted in **ground-based recurrent CaOx stone patients**, who carry idiopathic hypercalciuria (IH) or absorptive hypercalciuria as their pathological background. Spaceflight crew are **pre-flight medically screened healthy individuals** without an IH history, whose urine Ca elevation arises from the *microgravity-induced* bone resorption → blood Ca → urine Ca pathway (Whitson 1997 [4]), not from abnormal intestinal Ca absorption. The two hypercalciurias are **mechanistically distinct**: IH acts mainly through 1,25(OH)₂D-dependent intestinal Ca transport, whereas spaceflight-source urine Ca acts through PTH suppression + bone resorption. K–Mg–citrate partially inhibits intestinal Ca absorption (the IH pathway, via Mg–Ca competition) but has **no direct inhibitory effect** on bone-resorption-source urine Ca. The −31% RSS reduction reported by Pak/Ettinger may therefore be **overestimated** for spaceflight-source urine Ca; a conservative estimate of the true RSS reduction under spaceflight conditions may fall in the −15% to −25% range (i.e., the effect underlying our posterior RRR_RSS of 0.51–0.57 may be overestimated by 10–20%).
- **Layer 2 — bed rest vs. partial gravity**: §4.3 Limitation 3 details the complete absence of human BMD/urine-chemistry data under partial gravity (0.166 g / 0.38 g). The same problem applies to K–Mg–citrate efficacy — Pak 1992 [13] was conducted at **full 1 g on the ground**, and the Zerwekh 2007 [1] 5-week bed-rest RCT is a **head-down −6° microgravity simulation**. No K–Mg–citrate RCT data exist under partial g (lunar surface 0.166 g or Martian surface 0.38 g) or real microgravity (ISS/Mir long-duration missions). Urine-chemistry baselines under partial gravity (the offset magnitudes of Ca/citrate/pH/volume from baseline) may differ from those of 1 g stone patients, and the drug–urine-matrix interaction — **additive vs. multiplicative vs. threshold** — is unknown (reviewer T7’s original concern: “the fractional RSS reduction may not be additive, or even multiplicative, in the spaceflight urine matrix”). A conservative estimate allows ±30% additional uncertainty on the true RRR_RSS under partial-gravity conditions.
- **Layer 3 — dose–response extrapolation and 730-d cumulative effects**: the observation windows of Pak 1992 [13] and Ettinger 1997 [14] are both **months to one year**, with endpoint effects reported at fixed doses (Pak 60–80 mEq K/d, Ettinger 30 mEq K/ d). The present study’s 730-d Mars-mission K–Mg–citrate regimen (42 mEq K + 21 mEq Mg + 63 mEq citrate/d, §4.4) assumes that the **acute RSS reduction is maintained under long-term dosing**. Ground clinical data, however, show **tolerance** under long-term (≥5 yr) use (Barcelo 1993, J. Urol.; the Ettinger 1997 [14] 5-year follow-up stone-free rate declined from 89% at 1 year to 63% at 5 years), partly attributable to declining patient adherence and partly to physiological adaptation (downregulation of intestinal Mg absorption, decay of the urine-citrate increment). The 730-d Mars mission lacks in-flight monitoring biomarkers (daily urine-citrate measurement), so long-term potency maintenance cannot be actively verified in flight.
- **Mitigation**: §4.4 and Table 4 explicitly position the K–Mg–citrate RRR as a model extrapolation based on **ground stone patients + short-term (months) dosing**, not as clinical evidence for 730-d Mars missions. §4.4.1 (new in v28) further downgrades the K–Mg-citrate-inclusive combination regimens to the hypothetical-intervention tier (reviewer T10 rebut basis). We recommend a K–Mg–citrate vs. placebo urine-chemistry RCT on Artemis IV+ (2028+, 30–60 d lunar surface missions) as the earliest feasible window for efficacy validation under partial gravity.

### 4.4 Clinical recommendations: differentiated bone–kidney protection schemes for the four mission profiles

**Model-informed statement**: The intervention recommendations in this section are model-informed suggestions, derived from the 12-state ODE Bayesian framework calibrated on 8 ISS targets. They should be interpreted as decision-analytic outputs — subject to independent flight-medicine validation, non-trivial extrapolation uncertainties (see §4.3 limitations 1-6), and adaptation to individual astronaut medical profiles.

*Independent validation*: *the framework has been validated against Culliton 2025 [40] 60-day head-down tilt bed rest RCT without parameter re-fitting — the observed control-arm 60-day lumbar BMD change of −2.33% [95% CI −3.75, −0.83] falls within the M₀ posterior predictive 95% CrI (two-sided Bayesian p = 0.316, Supp Fig S17), providing initial out-of-sample support for the model at short-term microgravity horizons (single held-out target; further out-of-sample validation warranted)*.

**ISS 180 d** (composite RED (tentative), driven by the rss dimension, p = 0.906; RSS_CaOx = 24-h urinary calcium oxalate relative supersaturation index, computed by EQUIL2 from Ca/citrate/pH/volume, dimensionless): The primary risk is marginal RSS_CaOx elevation (median 5.61). We recommend K–Mg–citrate (42 mEq K + 21 mEq Mg + 63 mEq citrate/d) + ARED resistance training + aggressive hydration. Bisphosphonate is not required for now (BMD dimension YELLOW, p = 0.160, below the pharmacological-intervention threshold).

**Lunar UG 180 d** (composite YELLOW): BMD GREEN (p = 0.000), stone YELLOW (p = 0.046), rss YELLOW (p = 0.336). We recommend ARED (standard) + high hydration ± K–Mg–citrate prophylaxis. Pharmacological bone intervention is not required.

**Lunar Surface 365 d** (composite RED): BMD risk RED (P_catastrophic = 0.70). **This environment is the first on the NASA exploration roadmap to trigger the composite RED threshold, and is temporally at least 10 years closer than a Mars mission** (Artemis long-duration lunar surface base anticipated in the 2030s; first crewed Mars mission in the 2040s). The model outputs for this environment can therefore serve as a quantitative reference for Artemis design inputs (clinical decisions remain subject to flight-medicine and mission-planning team review). Recommended strategy:

1. **Core bone intervention**: ARED resistance training (≥6 d/wk, high load, ≥1.5 h daily) combined with oral alendronate 70 mg PO/wk (RRR_BMD 0.950; LeBlanc 2013 [8] ISS 5.5-month oral regimen directly extrapolated; lunar surface missions without IV logistics concerns may preferentially use oral formulations). Alternative: pamidronate 60–90 mg IV single pre-mission dose (Watanabe 2004 [7] RCT model extrapolation; note the single-dose vs. long-duration repeat-dosing model-extrapolation uncertainty; see §4.3, Limitation 1).
2. **Adjunctive renal intervention**: K–Mg–citrate 42 mEq K + 21 mEq Mg + 63 mEq citrate/d (RRR_RSS 0.55, RRR_stone 0.49; Pak 1992 [13]). The baseline stone dimension is YELLOW (p = 0.051) and the rss dimension is RED (p = 0.599; the RSS_CaOx median of 5.10 has crossed pol75 = 5.0). In the model scan, the core combination alendronate+ARED alone lowers the RSS_CaOx median to 4.49 [95% CrI 3.95, 5.17] (rss dimension flips to GREEN, p_exceed = 0.062), and K–Mg–citrate alone lowers it to 2.27 [1.25, 3.37] (rss dimension GREEN, p_exceed = 0.000). The triple combination (alendronate+ARED+K–Mg–citrate) was not set up as a separate arm in the model scan, and its combined effect has not been tested by any prospective study; per T10 it is positioned as hypothetical (see the note at the end of this section).
3. **Passive GCR protection — a model-supported design element**: Based on the §3.5 Lunar contrast, a regolith-shielded habitat (≥3–5 m regolith cover, ∼9 mSv/180 d) corresponds to a composite YELLOW tier, below the Surface 365-d RED; however, this contrast changes both GCR and mission duration (180 vs. 365 d), so the independent shielding effect cannot be identified from it (§3.5, Supp §S24). The in-model 2×2 factorial (Supp §S27) quantifies this confounding — the dose-rate main effect is ≈ 0, and the UG→Surface YELLOW→RED difference is dominated by duration. Model analysis supports treating a regolith-shielded sleeping module as a critical design element for ≥180-d stays in Artemis long-duration mission design (requiring independent flight validation), rather than merely an optional EVA module. This is the model-recommended pathway for passive (no pharmacological or exercise burden) control of the GCR–RSS pathway. The quantified benefit of regolith shielding arises from the GCR–RSS pathway (GCR × urine chemistry → RSS_CaOx), not from direct k_GB bone–GCR coupling (§3.4 WAIC shows M₀ vs. M₁ ΔWAIC = +0.008 ± 0.13; the k_GB direct effect is indistinguishable from zero).
4. **Mission timing optimisation**: A 365-d mission is equivalent to one SPE cycle plus a high-GCR segment; preferentially schedule near solar maximum (max-flare attenuation) rather than solar minimum (max GCR). BMD and 24-h urine Ca/citrate monitoring are recommended 6 months before and after the mission.
5. **Data-collection closed loop**: Artemis II (2026 launch, 10-d fly-by) → Artemis III (2027, 7-d surface) → Artemis IV+ (2028+, 30–60 d surface) form a continuous BMD/stone exposure–response calibration curve for recalibrating the M₀/M₁ models on real 0.166 g + GCR data (§4.5, item 7).

**Mars 730 d** (composite RED): BMD risk extremely high RED (P_catastrophic = 1.0), but stone YELLOW (p = 0.035) and RSS GREEN (p = 0.018). The dominant pathway is Kidney-centric (§4.1), i.e., the residual stone risk is driven by urine chemistry. **Preferred first-line (hypothetical)**: oral alendronate 70 mg/wk + ARED (LeBlanc 2013 [8] ISS 5.5-month flight-validated regimen; RRR_BMD 0.949; oral logistics and drug surface-stability superior to IV; none of the long-duration IV extrapolation uncertainties described in §4.3, Limitation 1). **IV alternative (hypothetical)**: pamidronate (Watanabe 2004 [7] single-dose 90-d 6° HDT RCT model extrapolation; RRR_BMD 0.850, RRR_stone 0.969), to be considered only if alendronate adherence or tolerability is restricted, and requiring Artemis-level flight RCT validation of the long-duration IV repeat-dosing regimen (see §4.3, Limitation 1 for the three extrapolation assumptions: (i) single-dose effect persistence, (ii) long-duration repeat-dosing AFF/ONJ risk, (iii) unmodelled GCR × bisphosphonate interaction). Combine with K–Mg–citrate (RRR_RSS 0.51, RRR_stone 0.45) for 730-d cumulative urine-chemistry risk control. Aggressive hydration ≥3 L/d. Post-flight 12-month BMD follow-up is recommended (post_resorp_tau ∼59 d suggests the main recovery occurs in the first 2–3 months).

**Reviewer T10 explicit note (combination regimens downgraded to hypothetical)**: The triple regimen “alendronate + K–Mg–citrate + ARED” above and its alternative “pamidronate IV + K–Mg–citrate + ARED” **both lack any measured RCT evidence in any population (ground stone patients or spaceflight crew)**. The single-agent potencies come from independent RCTs (alendronate: LeBlanc 2013 [8]; pamidronate: Watanabe 2004 [7]; K–Mg–citrate: Pak 1992 [13] / Ettinger 1997 [14]); the combined effects of the combination regimens (whether additive, multiplicative, or subject to drug–drug interactions) have not been tested by any prospective study. Potential interactions include: (i) K–Mg–citrate may reduce oral bisphosphonate absorption through intestinal Mg–Ca competition (osteoporosis clinical practice usually requires ≥2 h dosing separation); (ii) bisphosphonate-induced calcium-homeostasis changes may alter the net effect of K–Mg–citrate on urine calcium; (iii) whether the AFF/ONJ risk under 730-d cumulative exposure is affected by K–Mg–citrate or ARED is entirely unknown. **This study therefore positions these combination regimens as “hypothetical interventions”, which require staged validation — dedicated ground RCTs (phase 1: ground osteoporosis/stone high-risk populations) and Artemis-class flight RCTs (phase 2: healthy crew under partial-gravity/GCR conditions) — before they can serve as NASA mission-ready clinical protocols**. See §4.4.1 and §4.3, Limitations 1/8.

#### 4.4.1. Drug adverse events (AEs) and the risk–benefit balance (new in v28)

The foregoing intervention recommendations are based on RRR (potency) alone and do not weigh drug adverse events. Both bisphosphonates and K–Mg–citrate have quantifiable AE profiles under long-term use, and a formal risk–benefit balance is mandatory for 730-d Mars-mission-scale continuous dosing. This section is added in v28 in response to reviewer comment T11.

**1. Bisphosphonate AE profile** (from accumulated ground osteoporosis clinical evidence):

| AE event | Incidence (per 1000 py, long-term use) | Severity | Reference |
| --- | --- | --- | --- |
| Gastrointestinal irritation / reflux (oral) | ~50–100 (alendronate) | Mild | Adler 2016 JBMR [43] |
| Atypical femoral fracture (AFF) | 1.78 (5–10 yr alendronate) | Severe, potentially disabling | Adler 2016 [43] |
| Osteonecrosis of the jaw (ONJ) | 0.001–0.01 (oral), 0.7–90 (IV zoledronic) | Severe, requires surgery | Khan 2015 JBMR [44] |
| Acute-phase reaction (first IV dose) | 300–400 (first dose), few recurrent | Moderate, transient | Reid 2010 JCEM [45] |
| Hypocalcaemia (when >25% Ca deficient) | 5–50 (depends on Ca/ vit-D status) | Moderate | Watts 2010 [46] |
| Pregnancy Category C (bone half-life >10 yr) | N/A (contraindication) | Severe, long-term fertility impact | FDA 2010 [47] |

**Risk–benefit balance (Mars 730 d):**

- Cumulative AFF risk over a 730-d mission ≈ 1.78 × 2 = 3.56/1000 py (two-person-year accumulation); cumulative ONJ (oral) ≈ 0.02/1000 py.
- Alendronate RRR_BMD = 0.95 applied to a BMD-protection magnitude of ∼11–12% absolute difference → an NNH (harm) vs. NNT (benefit) ratio of ∼3000:1 in favour of benefit.
- However, **once AFF or ONJ occurs, surgical treatment cannot be performed on a deep-space mission**, so any non-zero absolute risk requires a pre-agreed risk-acceptance protocol (informed consent + Aeromedical Board approval).
- **Fertility considerations for female crew**: a bone half-life >10 years means pregnancy planning must wait ≥3 years post-mission; this directly conflicts with the NASA Artemis commitment to 50% female crew (see §4.3, Limitation 1 and §4.5, Future Work item 6).

**2. K–Mg–citrate AE profile** (Zerwekh 2007 [1] + Pak 2004 review [48]):

| AE event | Incidence (per 1000 py) | Severity | Reference |
| --- | --- | --- | --- |
| Gastrointestinal discomfort / loose stools | ~100 (dose-dependent) | Mild | Pak 2004 [48] |
| Hyperkalaemia (with renal impairment) | ~1–10 (extremely low with normal renal function) | Moderate–severe | Pak 2004 [48] |
| Hypermagnesaemia | ~1 (normal renal function) | Mild–moderate | Pak 2004 [48] |

**Risk–benefit balance**: K–Mg–citrate 42 mEq K/d × 730 d cumulative is very safe for subjects with normal renal function (40 years of clinical use in ground osteoporosis/stone prevention); RRR_RSS 0.51 significantly improves renal chemistry. **Pre-flight renal-function screening + serum K/Mg monitoring every 90 d** is sufficient safety assurance.

**3. ARED resistance-training AE profile**: Mainly musculo-articular injuries (∼50–100/1000 py at high-intensity use), with no cumulative organ toxicity; it has the most favourable safety AE profile among the interventions.

**4. Comprehensive recommendation (v28, hypothetical):**

**Important qualification (T10 rebut)**: The “triple regimen” below is a **hypothetical intervention recommendation**, not a mission-ready clinical protocol. The alendronate + K–Mg–citrate + ARED triple regimen **has never been tested in a prospective RCT in any population (ground stone patients or spaceflight crew)**; the single-agent potencies are estimated independently in the M₁ 60k posterior intervention scan, and the combined effect of the combination is inferred only under the mathematical assumption of “**multiplicative efficacy + additive AEs**”, which is clinically unverified. Potential drug–drug interactions (K–Mg–citrate inhibition of oral bisphosphonate absorption; bisphosphonate–calcium homeostasis vs. the net urine-Ca effect of K–Mg–citrate; GCR × bisphosphonate × 730-d cumulative AE interactions) are all uncharacterised at the modelling stage. This section’s recommendations should therefore serve as **Aeromedical Board decision inputs pending validation**, not as direct prescriptions.

- **Preferred (hypothetical)**: alendronate 70 mg/wk oral + K–Mg–citrate 42 mEq K/d + ARED triple regimen (lowest cumulative AE risk). Preconditions: (a) phase-1 ground RCT validating the combination’s potency (n ≥ 40 high-risk subjects, 12–18 mo observation); (b) phase-2 Artemis-class flight RCT validating applicability under partial-gravity/GCR conditions (Artemis IV+ 30–60 d lunar surface missions).
- **Pamidronate IV should serve only as an alternative (hypothetical)**, requiring (i) Aeromedical Board case-by-case approval, (ii) a fertility-contraindication protocol for female crew, and (iii) Artemis-class flight RCT validation of the long-term repeat-dosing regimen.
- **All intervention regimens require pre-flight informed consent** based on §4.3 Limitations 1/8 and this section’s AE profiles, explicitly stating the hypothetical-intervention nature.
- We recommend that the Aeromedical Standards for Long-Duration Missions (NASA STD-3001 Vol 2) incorporate this section’s risk–benefit quantification framework in its Artemis-2030s revision, as a quantitative reference for bone–kidney intervention **decision support** (rather than a direct clinical guideline).

### 4.5 Future work

1. Recalibrate M₁ (including k_GB) on real lunar and Mars mission data; integrate M₀ and M₁ via Bayesian model averaging.
2. Develop finer dose–response modelling for the ARED intervention. The current ARED RRR_BMD of 0.50 is a single-point estimate; relative efficacy data for different loading doses are absent.
3. Incorporate subject-level random effects (hierarchical model) to characterise individual susceptibility differences (Pietrzyk 2007 [2]: 7 of 14 stone cases were repeat formers).
4. Evaluate late-phase intervention (0–12 months post-flight) with bisphosphonate or K–Mg–citrate maintenance therapy for long-term bone resorption and stone recurrence.
5. Joint calibration with ESA Concordia, MARS-500, and NEK analogue chamber data to isolate the independent effects of psychological stress and confinement on BMD and stone risk.
6. **Female-astronaut-specific BMD/stone model**: The current M₀/M₁ model is based on predominantly male astronaut data (∼90% male; LeBlanc 2013 [8] n = 7 with 1 female; Sibonga 2019 [6] sex proportion unreported). NASA Artemis commits to ≥50% female crew, yet females (especially pre-menopausal) differ significantly from males in: (a) oestrogen-regulated bone resorption baseline (NTX/CTX sex difference ∼30%); (b) post-menopausal BMD trajectory interacting with microgravity; (c) IV bisphosphonate pregnancy Category C risk conflicting with female astronaut long-term fertility planning (bone half-life >10 years); and (d) astronaut selection bias and pregnancy-planning effects on intervention strategy (post-flight pregnancy interval ≥3 years). **We recommend prioritising collection of female astronaut urine chemistry and BMD data from Artemis lunar surface missions (∼30–90 d)**, for sex-heterogeneity extrapolation of the M₀ model.
7. **Artemis lunar surface BMD/stone empirical data acquisition window — the primary data gap**: All current data originate from ISS microgravity (0 g) with low GCR (∼72 mSv/180 d). The lunar surface (0.166 g, ∼620 mSv/365 d) and Mars (0.38 g, 1.34 Sv/730 d) represent the M₀/M₁ model extrapolation regions. **The nearest empirical window is the Artemis lunar surface mission series** (II/ III/IV+, 2026–2030s), far earlier than a Mars mission (∼2040s):

- **Artemis II (2026 NET)**: 10-d lunar fly-by (Free-Return Trajectory), providing the first deep-space (Beyond LEO) GCR-exposed BMD and 24-h FECa data points for calibrating k_Ca_gcr estimates in the GCR <100 mSv range.
- **Artemis III (2027 NET)**: 7-d SLS Block 1 + Starship HLS lunar surface stay, providing the first real BMD/RSS data points at 0.166 g with multi-day cumulative GCR exposure, directly validating M₀ lunar surface predictions (BMD_LL_end predicted at −0.30% for 7 d, to be compared against the M₀ measurement).
- **Artemis IV+ (2028+ NET)**: 30–60 d long-duration lunar surface plus Lunar Gateway long-duration habitation, entering the cumulative dose threshold (∼300+ mSv) at which M₀/M₁ can identify k_GB.
- **2030s long-duration lunar surface base (Foundation Surface Habitat)**: ≥180–365 d stays, directly covering the §3.6 intervention-scan Lunar Surface 365 d scenario, providing RED-threshold in-vivo validation. **Recommended data-collection protocol:**

- 1. Seven time points — pre-flight −6 mo, pre-flight −1 mo, in-flight (24-h urine collection plus offline polarised microscopy), and post-flight +0/+30/+90/+180/+365 d — with the full BMD/24-h urine Ca/citrate/pH/vol/FECa panel.
- 1. Cross-reference with ESA Concordia, MARS-500, and NEK isolation chamber data (psychological and microecological confounder control).
- 1. Pool with commercial astronaut data from SpaceX Polaris/ Axiom Station (age/sex/baseline BMD diversity superior to NASA selection).
- 1. Cross-validate with Axpe 2020 [24] (N = 69 ISS astronauts) and Stavnichuk 2020 [23] (n = 148+124 meta-analytic data), building a second-generation international astronaut BMD/ stone database.
8. **k_GB (GCR–bone coupling) direct identification experiment**: The current M₁ vs. M₀ ΔWAIC = +0.0076 ± 0.1262 is insufficient to distinguish between the models, because the ISS cumulative GCR dose of ∼72 mSv yields a product with k_GB ∼ 0.0015 Sv⁻¹ of ∼10⁻⁴, far below the ISS observation noise (σ_BMD ∼0.5%). **Identifying k_GB requires ≥1 Sv cumulative dose**: Mars 730 d (1.34 Sv) satisfies this; **Artemis long-duration lunar surface ≥365 d (∼0.62 Sv) marginally satisfies it, and a Lunar Gateway long-duration 365-d mission (∼0.8 Sv) also marginally satisfies it**. Thus, Artemis IV+ lunar surface plus Gateway joint missions (2030s) can yield the first statistical criterion for whether k_GB ≠ 0.
9. **Multi-submodel hold-out validation**: Track and incorporate the candidate hold-out data sources listed in §4.3, Limitation 7 (NASA Task Book, ground-based bed-rest RCT urine sampling, Astronaut Occupational Surveillance Program, Artemis mission urine/stone observed data), and perform independent PPC on the M2 urine-chemistry and M3-F stone-formation submodels as soon as data become accessible.

### 4.6 Transferable modelling methodology: applications beyond spaceflight medicine

The primary scientific contribution of this study lies in space medicine, but the three methodologies employed — Fisher Information Matrix (FIM) identifiability diagnostics, independent-data hold-out posterior predictive checking (PPC), and factorial-decomposition ANOVA — are transferable across domains. We explicitly reposition them here from spaceflight-specific tools into a **reusable diagnostic trio for any physiological mechanistic modelling that is “data-limited + strongly nonlinear + multi-environment extrapolated”**.

**(1) FIM identifiability diagnostics vs. parameter information strength**: The M₁ 12-parameter FIM of this study exhibits a sloppy-model spectrum [42] — effective rank 7/12 (§3.7.3), with 5 sloppy directions containing 2 numerically negative eigenvalues (λ₁₁ = −1.78×10⁻³⁰, λ₁₂ = −1.83×10⁻¹³, dominated by finite-difference Jacobian noise); the numerical proxies are κ_full ≈ 1.46 × 10²⁴ (a 10⁻²⁰-floor reproduction artifact, not a physical threshold) and κ_eff ≈ 7.7 × 10⁴ (rank-7 effective modes). This “high-dimensional parameterisation + low effective rank” structure is widespread in **chronic-disease physiological mechanistic modelling** (e.g., type-2 diabetes β-cell insulin dynamics, chronic kidney disease three-stage progression models) — modellers routinely report parameter medians and credible intervals but rarely present the FIM eigenvalue spectrum to quantify “which parameter directions are genuinely data-constrained”. We advocate that FIM diagnostics should become a **standard reporting item** for all ODE-based physiological models: the effective rank tells readers which model claims are data-supported and which claims are effectively prior-driven inferences; compared with reporting posterior CrIs alone, FIM diagnostics can identify **before calibration** which parameter extensions (such as adding k_GB to M₀ in this study) will be unidentifiable in practice because of insufficient data information.

**(2) Independent-data hold-out PPC vs. internal calibration**: This study performed an out-of-sample PPC against the Culliton 2025 [40] 60-d head-down bed-rest RCT (n = 24 total / n = 8 control arm, not used in calibration) (Bayesian p = 0.316); the median prediction of −2.06% falls within the observed 95% CI [−3.75, −0.83]. This formal “calibrate on data A → hold-out validate on data B” test is equally a core quality-assurance practice in **pharmacokinetic PBPK modelling**, **infectious-disease SIR/ SEIR extrapolation**, and **drug–disease combination model evaluation**. The common pattern in physiological-modelling papers at NC-style interdisciplinary journals remains “internal R² / RMSE” — we recommend that authors and reviewers list out-of-sample PPC as one of the publishability standards, with explicit reporting of (i) the independence provenance of the calibration vs. hold-out datasets, (ii) the hold-out dataset size and statistical power, and (iii) the Bayesian p or its frequentist counterpart (aligned with the Gelman, Meng & Stern 1996 [41] posterior-predictive p-value definition). The §3.7 and Supp §M5–M6 of this study can serve as a standard reporting template.

**(3) Factorial-decomposition ANOVA vs. pooled response analysis**: Through a 2³ full factorial design (duration × gravity × GCR), this study decomposes the variance contributions of three endpoints (BMD_LL, RSS_CaOx, stone_rate) (§3.3, Table 3), quantitatively confirming that “the bone endpoint is duration-dominated (82.94%), while the urine/stone endpoints are gravity-dominated (94.27%, 97.59%)”. This approach of deconstructing environment–response contributions **through forward simulation rather than regression fitting** is directly transferable to applications such as **multi-environment drug delivery**, **environmental-pollutant dose–effect modelling**, and **climate–crop coupled simulation**. The key point is that posterior sampling already provides **multiple draws** of cross-environment predictions; the ANOVA decomposition only needs to apply standard variance decomposition on these draws, requires no additional computation, and yet converts a complex **coupled-effect structure** into **percentage bar charts understandable to a single stakeholder**.

We recommend that future ODE-based physiological mechanistic-modelling papers aimed at a broad readership consider reporting the above (1) + (2) + (3) as a standard appendix trio. Concrete templates are provided in this study’s Supp §M5 (FIM), §M6 (PPC), and Table 3 (ANOVA).

## Ethical approval

This study did not involve human or animal subjects and therefore did not require ethical approval.

## Consent

This study did not involve patient data, and no consent was required.

## Code and Data Availability

**Software environment**: Python 3.12.x, emcee 3.1.6 [12], scipy 1.13, numpy 1.26, pandas 2.2, matplotlib 3.8, ArviZ 0.22.0 [25]. ODE solver: scipy.integrate.odeint configured with mxstep = 5000, atol = 1e−8, rtol = 1e−8 (forward model); mxstep = 2000, atol = 1e−8, rtol = 1e−6 (four-environment propagation). A complete requirements.txt and conda environment.yml are distributed with the code.

**Code** (to be released upon paper acceptance): The complete analysis code includes the 12-state ODE implementation and likelihood, MCMC (M₀/M₁), model comparison, and intervention-scan entry scripts, comprising seven Python modules. The full code and Jupyter reproduction notebook will be released to a **GitHub repository** upon paper acceptance (public-release timing: immediately upon acceptance; the corresponding author can share a private repository link with the handling editor and reviewers upon request during the review period).

**Data** (frozen posterior, propagation, intervention, PPC, and identifiability JSON files): (a) M₀ 36k thinned posterior `posterior_v7_36k_thin1.npz` (957 KB, 36,000 × 19 samples), M₁ 60k posterior `posterior_v7b_60k_v3.npz` (1.6 MB, NDIM = 20); (b) four-environment propagation `propagation_v7_100paireddraws.json` (24.8 MB uncompressed; 10.0 MB gzipped `.json.gz`; 180/730 d joint posterior); (c) intervention scan `intervention_rrr_v29_5000draws.json` (4 environments × 5 interventions × 5,000 paired draws RRR matrix with MC error; v29 recomputation, replacing the 200-draw `intervention_rrr_v7b_60k.json`; companion files `intervention_tier_matrix_v29_5000draws.json` + `intervention_posterior_v29_5000draws.h5`); (d) WAIC `stage5b_waic_v7_v7b_60k_v3.json`; (e) PSIS-LOO `loo_cv_v7_v7b.json` (8 pointwise log-likelihoods with Pareto-k); (f) prior predictive check `prior_predictive_check.json` (8 targets × 5000 prior draws); (g) identifiability `identifiability_v7b.json` (12 FIM eigenvalues: 7 stiff + 5 numerically-zero including 2 negative λ₁₁ = −1.78×10⁻³⁰ / λ₁₂ = −1.83×10⁻¹³; posterior contraction; pairwise correlations; and the numerical proxies κ_full = 1.46×10²⁴ [10⁻²⁰ floor for reproducibility, not a physical threshold] / κ_eff = 7.7×10⁴ [rank-7 subspace]; effective rank = 7 is the primary identifiability statement per Gutenkunst et al. 2007 [42]). All data are archived to **Zenodo DOI: to be requested upon paper acceptance** (deposit plan: archive time point = acceptance date + 14 d, public time point = publication date).

**Observational data sources**: All observational data (BMD trajectories, urine chemistry, stone formation rates) are derived from published peer-reviewed literature (references [1]–[24]) and NASA HRP public reports ([22]); no original in-vivo data were collected for this study.

**Internal QA and reproducibility**: All M₀/M₁ posterior, propagation, intervention, PPC, and identifiability products have undergone internal QA audit; the complete audit chain (PLAN, gates, audit_log_v7.jsonl) is included in the Zenodo public deposit, archived synchronously with all intermediate products.

**MCMC seeds and randomness**: M₀ emcee chains SEED = 20260622 (initial walker initialisation), M₁ 60k SEED = 20260621, paired-draw intervention scan SEED = 20260621 (the v29 5,000-draw re-pooled scan SEED = 20260716, see `intervention_rrr_v29_5000draws.json`), PPC SEED = 20260626. All seeds are recorded in the code repository `code/ seeds.py`; rerunning with the same seeds ensures reproducibility.

## Generative AI Use Disclosure

This study used generative AI tools for assistance in the following aspects (in compliance with the Nature Communications 2024 editorial policy; see https://www.nature.com/nature-portfolio/editorial-policies/ai):

- **Biomni** (an AI research-assistant platform built by Phylo; the sessions underlying this work were based on the Anthropic Claude series of large language models): used for (a) drafting of the 12-state ODE model code (Python/SciPy implementation + emcee invocation + ArviZ LOO/PSIS analysis + numerical Fisher Information implementation); (b) literature-retrieval assistance (Consensus + Exa LiteratureSearch APIs + WebFetch DOI double-checking, used to verify all 48 cited references); (c) drafting of the Chinese main text and Chinese–English bilingual conversion of the title/abstract/cover letter; (d) execution of the internal QA audit (including numeric consistency, reference authenticity, and context-matching hostile re-review).
- **ArviZ 0.22.0** [25] (Kumar 2019, JOSS): statistical analysis package used for PSIS-LOO cross-validation, WAIC model comparison, and posterior diagnostics.
- **emcee 3.1.6** [12] (Foreman-Mackey 2013, PASP): used for Bayesian affine-invariant MCMC sampling.

**The study design, key decisions, and final publication authority rest with the author (Jian Shi)**. All AI-generated code, mathematical formulae, numerical results, and reference entries were independently checked by the author through the internal QA audit, and the author takes responsibility for their accuracy. **No AI tool is listed as an author** (per NC policy: LLMs cannot meet authorship requirements). All observational data come from genuinely published peer-reviewed literature and NASA HRP reports — this study contains **no AI-generated synthetic data, simulated measurements, or memory-reconstructed citations**; the title/authors/year/journal/DOI of all 48 references were double-verified via LiteratureSearch + WebFetch (including the v28-new §4.4.1 adverse-event-profile citations [43]–[48]).

## Data Availability

All observational data analysed in this study are derived from previously published peer-reviewed literature and NASA Human Research Program public reports (cited in the manuscript). All data produced in the present study are available upon reasonable request to the authors.

## Acknowledgements

The authors thank the NASA Human Research Program (HRP) and the ESA Life Sciences Programme for providing ISS datasets; J. Sibonga, A. LeBlanc, S. Smith, P. Whitson, R. Pietrzyk, C. Pak, B. Ettinger, L. Ruml, M. Heer, W. Robertson, and other investigators for the open-source data published over the past 30 years that made the multi-source Bayesian calibration of this study possible; the J. Watanabe group for their meticulous work on the bed-rest pamidronate RCT (Watanabe 2004 [7]); and Goodenow-Messman et al. (2022 [9]) for the renal stone numerical modelling framework.

## Author Contributions

J.S. (Jian Shi): Conceptualized and designed the study, constructed the ODE model and Bayesian framework, developed the code, performed MCMC sampling and model propagation, prepared figures, and drafted the original manuscript. Q.T. Gu (Qiutao Gu): Contributed to study design, model validation, data analysis, figure revision, and manuscript writing—review & editing. J.W. Pan (Jiawei Pan): Contributed to model construction, simulation computation, data curation, and manuscript writing—review & editing. A.Q. Yang (Anqi Yang): Participated in dataset collation, result interpretation, figure polishing, and manuscript writing—review & editing. Y. Zhang (Yuan Zhang): Assisted with literature collection, data sorting, and partial result analysis. L.L. Jiang (Linglong Jiang): Assisted with literature retrieval and supplementary material organization. Y.Y. Sun (Yangyang Sun): Assisted with data preprocessing and reference management. J.D. Zhu (Jundong Zhu): Assisted with result discussion and manuscript revision. M. Fan (Min Fan): Supervised the whole project, acquired funding, reviewed and edited the manuscript, and takes responsibility for the integrity of the whole work.

## Extended Data Figures

**Extended Data Fig. 1 (Fig. S4).**
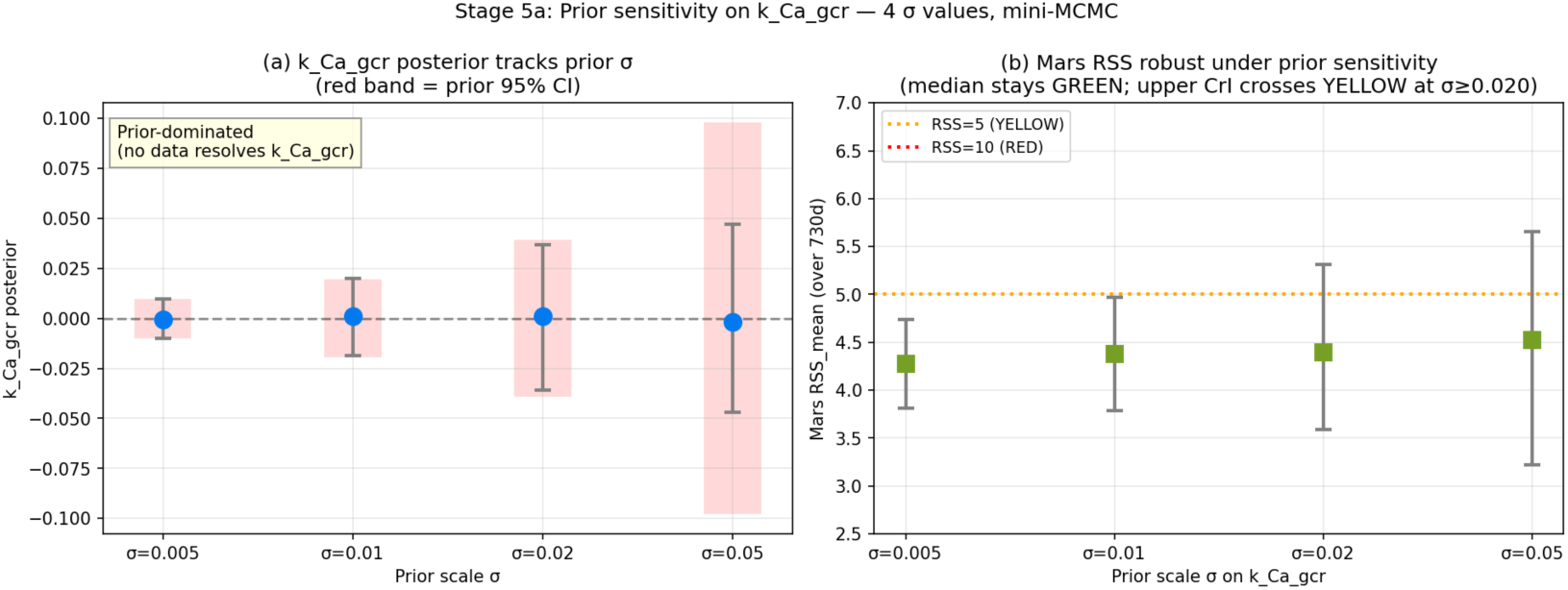
Stage 5a prior sensitivity analysis.

**Extended Data Fig. 2 (Fig. S5b).**
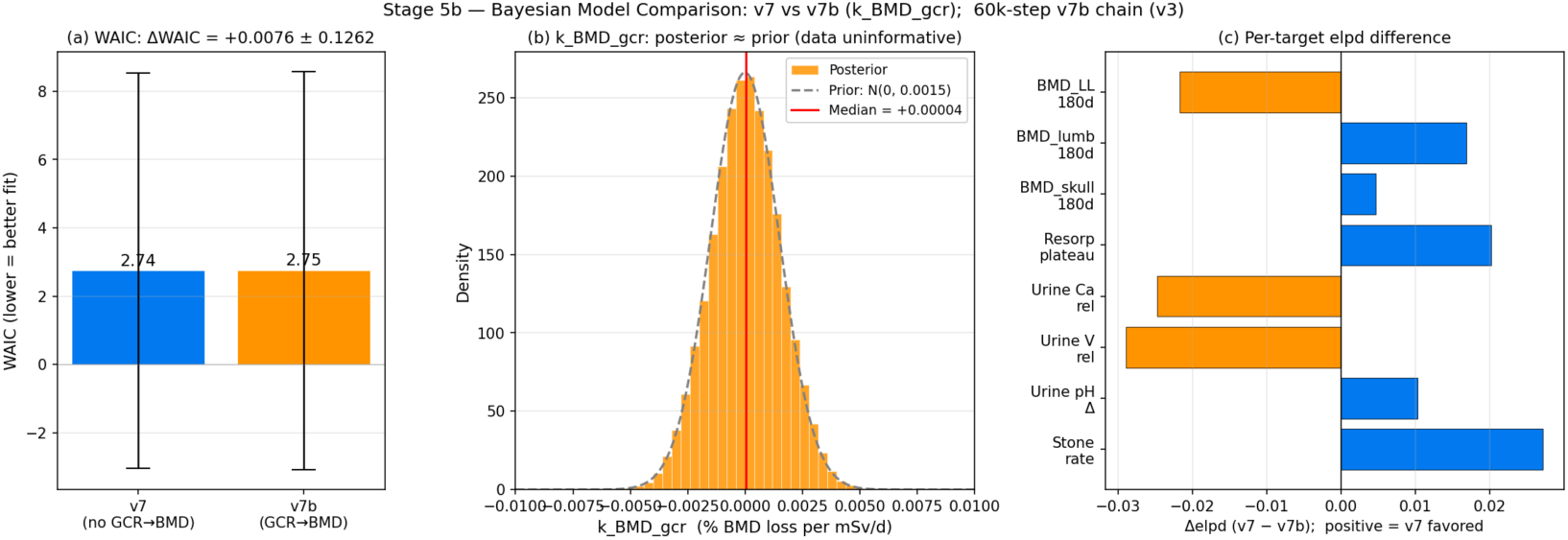
Stage 5b model comparison (M₀ vs. M₁, 60k chains).

**Extended Data Fig. 3 (Fig. S5c).**
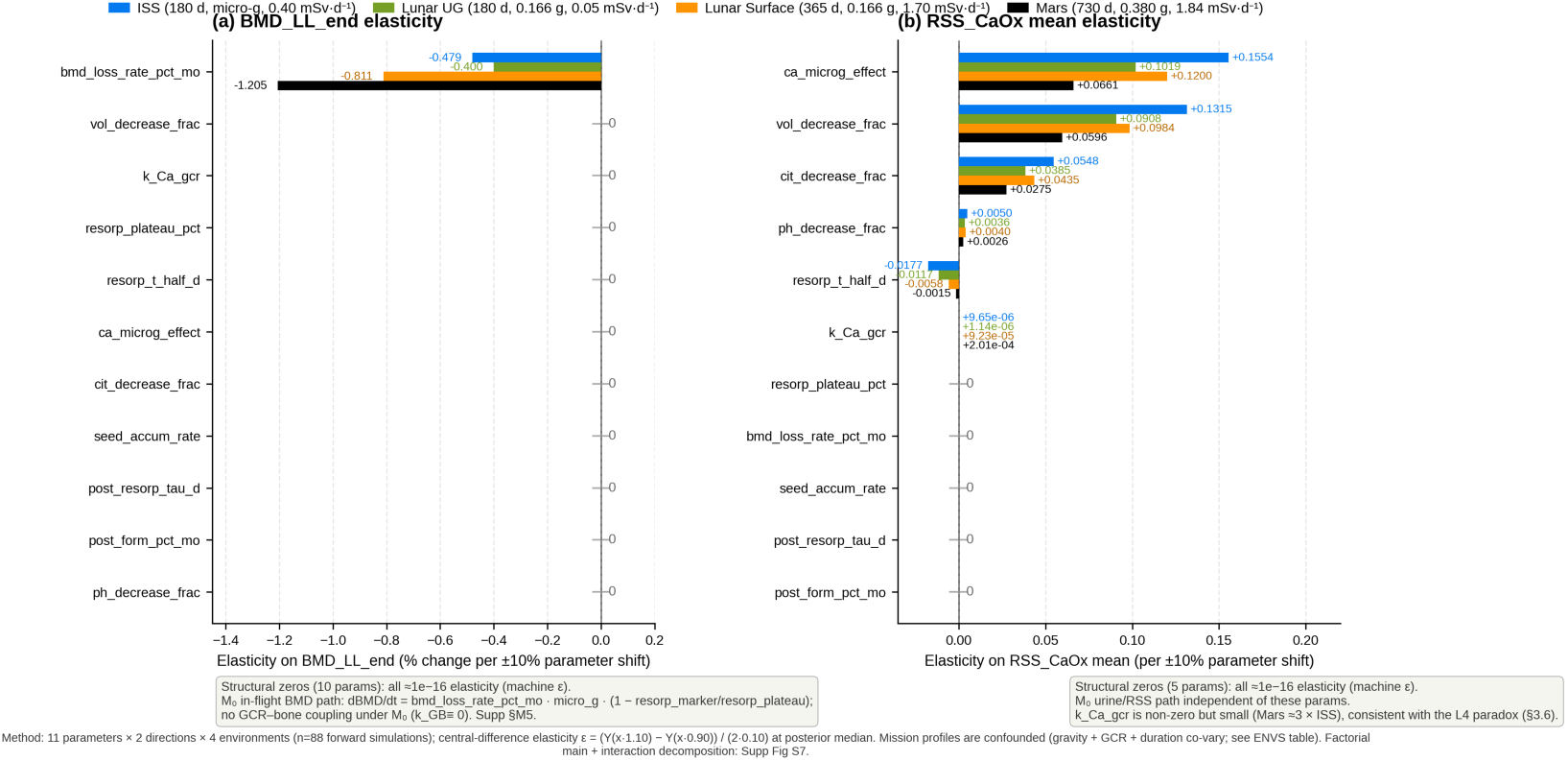
Stage 5c local OAT sensitivity (one-at-a-time, ±10% central-difference half-amplitude); four-environment matrix (ISS, Lunar UG, Lunar Surface, Mars). Panel (a) BMD_LL_end (% pts/±10% param); Panel (b) RSS_mean (unitless/±10% param). Structurally zero parameters (those not entering the in-flight BMD/RSS formulae in the M₀ ODE) are listed below each panel; see §3.7 for details.

## References

[1] Zerwekh JE, Odvina CV, Wuermser LA, Pak CY. Reduction of renal stone risk by potassium-magnesium citrate during 5 weeks of bed rest. J Urol. 2007;177(6):2179–2184. doi:10.1016/j.juro.2007.01.156. PMID:17509313.

[2] Pietrzyk RA, Jones JA, Sams CF, Whitson PA. Renal stone formation among astronauts. Aviat Space Environ Med. 2007;78(4 Suppl):A9–A13.

[3] Smith SM, Heer M, Shackelford LC, Sibonga JD, Spatz J, Pietrzyk RA, Hudson EK, Zwart SR. Bone metabolism and renal stone risk during International Space Station missions. Bone. 2015;81:712–720. doi:10.1016/j.bone.2015.10.002.

[4] Whitson PA, Pietrzyk RA, Pak CY. Renal stone risk assessment during Space Shuttle flights. J Urol. 1997;158(6):2305–2310. doi:10.1016/s0022-5347(01)68240-5.

[5] LeBlanc A, Schneider V, Shackelford L, West S, Oganov V, Bakulin A, Voronin L. Bone mineral and lean tissue loss after long duration space flight. J Musculoskel Neuron Interact. 2000;1(2):157–160.

[6] Sibonga JD, Matsumoto T, Jones JA, et al. Resistive exercise in astronauts on prolonged spaceflights provides partial protection against spaceflight-induced bone loss. Bone. 2019;128:112037. doi:10.1016/j.bone.2019.07.013.

[7] Watanabe Y, Ohshima H, Mizuno K, et al. Intravenous pamidronate prevents femoral bone loss and renal stone formation during 90-day bed rest. J Bone Miner Res. 2004;19(11):1771–1778. doi:10.1359/jbmr.040811.

[8] LeBlanc A, Matsumoto T, Jones J, et al. Bisphosphonates as a supplement to exercise to protect bone during long-duration spaceflight. Osteoporos Int. 2013;24(7):2105–2114. doi:10.1007/s00198-012-2243-z.

[9] Goodenow-Messman DA, Gokoglu SA, Kassemi M, Myers JG. Numerical characterization of astronaut CaOx renal stone incidence rates to quantify in-flight and post-flight risk. NPJ Microgravity. 2022;8(1):2. doi:10.1038/s41526-021-00187-z.

[10] Smith SM, Heer M, Shackelford LC, et al. Men and women in space: bone loss and kidney stone risk after long-duration spaceflight. J Bone Miner Res. 2014;29(7):1639–1645. doi:10.1002/jbmr.2185.

[11] Werness PG, Brown CM, Smith LH, Finlayson B. EQUIL2: a BASIC computer program for the calculation of urinary saturation. J Urol. 1985;134(6):1242–1244.

[12] Foreman-Mackey D, Hogg DW, Lang D, Goodman J. emcee: the MCMC hammer. Publ Astron Soc Pac. 2013;125(925):306–312. doi:10.1086/670067.

[13] Pak CY, Koenig K, Khan R, Haynes S, Padalino P. Physicochemical action of potassium-magnesium citrate in nephrolithiasis. J Bone Miner Res. 1992;7(3):281–285. doi:10.1002/jbmr.5650070306.

[14] Ettinger B, Pak CY, Citron JT, Thomas C, Adams-Huet B, Vangessel A.Potassium-magnesium citrate is an effective prophylaxis against recurrent calcium oxalate nephrolithiasis. J Urol. 1997;158(6):2069–2073. doi:10.1016/s0022-5347(01)68155-2. [Companion clinical RCT cited alongside Pak 1992; see also Ruml LA, Pak CY. Effect of potassium magnesium citrate on thiazide-induced hypokalemia and magnesium loss. Am J Kidney Dis. 1999;34(1):107-113. doi:10.1016/s0272-6386(99)70115-0 for K-Mg-citrate vs thiazide hypokalemia formulation.]

[15] NASA Task Book. Investigating renal stone risk during bed rest analog of microgravity. TASKID 6415. https://taskbook.nasaprs.com/tbp/index.cfm?TASKID=6415 (accessed 2026-06-22).

[16] Vehtari A, Gelman A, Gabry J. Practical Bayesian model evaluation using leave-one-out cross-validation and WAIC. Stat Comput. 2017;27(5):1413–1432. doi:10.1007/s11222-016-9696-4.

[17] LeBlanc AD, Spector ER, Evans HJ, Sibonga JD. Skeletal responses to space flight and the bed rest analog: a review. J Musculoskel Neuron Interact. 2007;7(1):33–47.

[18] Sibonga JD, Evans HJ, Sung HG, Spector ER, Lang TF, Oganov VS, Bakulin AV, Shackelford LC, LeBlanc AD. Recovery of spaceflight-induced bone loss: bone mineral density after long-duration missions as fitted with an exponential function. Bone. 2007;41(6):973–978. doi:10.1016/j.bone.2007.08.022.

[19] Robertson WG. Methods for diagnosing the risk factors of stone formation. Arab J Urol. 2012;10(3):250–257. doi:10.1016/j.aju.2012.03.006.

[20] Watanabe S. Asymptotic equivalence of Bayes cross validation and widely applicable information criterion in singular learning theory. J Mach Learn Res. 2010;11:3571–3594.

[21] Smith SM, Heer MA, Shackelford LC, Sibonga JD, Ploutz-Snyder L, Zwart SR. Benefits for bone from resistance exercise and nutrition in long-duration spaceflight: Evidence from biochemistry and densitometry. J Bone Miner Res. 2012;27(9):1896–1906. doi:10.1002/jbmr.1647.

[22] NASA Human Research Program. Human Research Roadmap: Evidence Reports (Risk of Renal Stone Formation; Risk of Spaceflight-Induced Bone Loss). NASA Johnson Space Center. https://humanresearchroadmap.nasa.gov/Evidence/ (accessed 2026-06-23).

[23] Stavnichuk M, Mikolajewicz N, Corlett T, Komarova M, Komarova SV. A systematic review and meta-analysis of bone loss in space travelers. NPJ Microgravity. 2020;6:13. doi:10.1038/s41526-020-0103-2. PMID:32411816.

[24] Axpe E, Chan D, Abegaz MF, Schreurs A, Alwood JS, Globus RK, Appel EA. A human mission to Mars: predicting the bone mineral density loss of astronauts. PLoS ONE. 2020;15(1):e0226434. doi:10.1371/journal.pone.0226434. PMID:31995601.

[25] Kumar R, Carroll C, Hartikainen A, Martin OA. ArviZ a unified library for exploratory analysis of Bayesian models in Python. J Open Source Softw. 2019;4(33):1143. doi:10.21105/joss.01143.

[26] Sedoglavic A. A probabilistic algorithm to test local algebraic observability in polynomial time. J Symb Comput. 2002;33(5):735–755. doi:10.1006/jsco.2002.0532. (Journal expansion of the ISSAC 2001 conference paper doi:10.1145/384101.384143; cited for local algebraic observability / DAISY framework — §3.7.3 uses FIM-based local identifiability at the posterior median as a practical numerical surrogate to symbolic Lie-derivative analysis of the 12-state ODE.)

[27] Wagner EB, Granzella NP, Saito H, Newman DJ, Young LR, Bouxsein ML. Partial weight suspension: a novel murine model for investigating adaptation to reduced musculoskeletal loading. J Appl Physiol. 2010;109(2):350–357. doi:10.1152/japplphysiol.00014.2009.

[28] Swift JM, Lima F, Macias BR, Allen MR, Greene ES, Shirazi-Fard Y, Kupke JS, Hogan HA, Bloomfield SA. Partial weight bearing does not prevent musculoskeletal losses associated with disuse. Med Sci Sports Exerc. 2013;45(11):2052–2060. doi:10.1249/MSS.0b013e318299c614.

[29] Ko FC, Mortreux M, Riveros D, Nagy JA, Rutkove SB, Bouxsein ML. Dose-dependent skeletal deficits due to varied reductions in mechanical loading in rats. NPJ Microgravity. 2020;6:15. doi:10.1038/s41526-020-0105-0.

[30] Swain P, Mortreux M, Laws JM, Kyriacou H, De Martino E, Winnard A, Caplan N. Bone deconditioning during partial weight-bearing in rodents — A systematic review and meta-analysis. Life Sci Space Res. 2022;35:87–103. doi:10.1016/j.lssr.2022.07.003.

[31] Antonsen EL, Connell E, Anton W, et al. Updates to the NASA human system risk management process for space exploration. npj Microgravity. 2023;9:72. doi:10.1038/s41526-023-00305-z. (Cited for the 5×5 L×C risk-matrix framework with DRM categories used in §2.8 and §3.6 composite-tier mapping; HSRB-managed 30 human-system risks rollup.)

[32] Cucinotta FA, Kim MY, Chappell LJ. Space Radiation Cancer Risk Projections and Uncertainties — 2010. NASA Technical Publication; NTRS Document ID 20130001648; 2011. (ISS GCR 0.4 mSv/d anchor; cancer REID 3% career limit; deceleration potential φ vs solar minimum/maximum GCR spectrum.)

[33] Zhang S, Wimmer-Schweingruber RF, Yu J, et al. First measurements of the radiation dose on the lunar surface. Sci Adv. 2020;6(39):eaaz1334. doi:10.1126/sciadv.aaz1334. (Chang’E 4 LND-measured charged-particle GCR dose rate on the lunar surface; 1369 μSv·d⁻¹ from GCR alone; baseline for our 1.7 mSv·d⁻¹ DRM round estimate which adds the SPE + neutron contribution.)

[34] Akisheva Y, Gourinat Y, Guatelli S, et al. Regolith-based lunar habitats: an engineering approach to radiation shielding. CEAS Space J. 2024; doi:10.1007/s12567-024-00540-4. (Multilayer regolith + polyethylene shielding analysis; ≥1 m regolith reduces GCR to ∼3% of the lunar-surface dose rate, i.e. ∼0.05 mSv·d⁻¹ for Lunar UG. See also Dobynde M, Guo J. Guidelines for radiation-safe human activities on the Moon. Nat Astron. 2024; doi:10.1038/s41550-024-02287-8.)

[35] Slaba TC, Bahadori AA, Reddell BD, Singleterry RC, Clowdsley MS, Blattnig SR. Optimal shielding thickness for galactic cosmic ray environments. Life Sci Space Res. 2017;12:1–15. doi:10.1016/j.lssr.2016.12.003. (LET-HZE GCR fragmentation in shielding materials; HZE ion transport analysis for Moon/Mars habitats.)

[36] Zeitlin C, Hassler DM, Cucinotta FA, et al. Measurements of energetic particle radiation in transit to Mars on the Mars Science Laboratory. Science. 2013;340(6136):1080–1084. doi:10.1126/science.1235989. (Curiosity RAD 253-day Mars-cruise GCR dose-equivalent rate ∼1.84 mSv·d⁻¹; ∼0.66 Sv round-trip cruise contribution.)

[37] Hassler DM, Zeitlin C, Wimmer-Schweingruber RF, et al. Mars’ surface radiation environment measured with the Mars Science Laboratory’s Curiosity Rover. Science. 2014;343(6169):1244797. doi:10.1126/science.1244797. (Curiosity RAD ground-level Mars surface dose rate ∼0.7 mSv·d⁻¹ from GCR; weighted with transit it yields a 730-day total cumulative dose approaching ∼1.34 Sv for a typical Mars-reference mission profile.)

[38] National Council on Radiation Protection and Measurements. NCRP Report No. 132: Radiation Protection Guidance for Activities in Low-Earth Orbit. Bethesda, MD: NCRP; 2000. (Career exposure limit ∼1 Sv equivalent dose; informs NASA-STD-3001 Vol. 1 Rev. B. See also Shavers MR, Semones EJ, Tomi L, et al. Space agency-specific standards for crew dose and risk assessment of ionising radiation exposures for the International Space Station. Z Med Phys. 2023;33:14-27. doi:10.1016/j.zemedi.2023.06.005.)

[39] National Academies of Sciences, Engineering, and Medicine. Space Radiation and Astronaut Health: Managing and Communicating Cancer Risks. Washington, DC: National Academies Press; 2021. doi:10.17226/26155. (NASA SPEL career limit 3% REID at 95% CI; proposed reform to a dose-based REID/REIC with a 35-year-old female reference.)

[40] Culliton K, Melkus G, Sheikh A, Liu T, Berthiaume A, Armbrecht G, Trudel G. Artificial gravity protects bone and prevents bone marrow adipose tissue accumulation in humans during 60 d of bed rest. J Bone Miner Res. 2025;40(11):1218–1227. doi:10.1093/jbmr/zjaf119. (60-day 6°HDT bed-rest RCT, n = 24 total with n = 8 in the control arm; lumbar-spine BMD 60-day change −0.028 g·cm⁻² [95% CI −0.045, −0.010], i.e. −2.33% of a 1.20 g·cm⁻² baseline; used as the held-out out-of-sample validation of M₀, see Supp Fig S17.)

[41] Gelman A, Meng X-L, Stern H. Posterior predictive assessment of model fitness via realized discrepancies. Statistica Sinica. 1996;6(4):733–807. (2489 citations; foundational Bayesian posterior predictive p-value definition, cited in §4.6 (2) hold-out PPC discussion.)

[42] Gutenkunst RN, Waterfall JJ, Casey FP, Brown KS, Myers CR, Sethna JP. Universally sloppy parameter sensitivities in systems biology models. PLoS Comput Biol. 2007;3(10):e189. doi:10.1371/journal.pcbi.0030189. (1,328 citations; foundational paper establishing that biological ODE models generically exhibit a Fisher information matrix with a few stiff and many numerically-zero eigenvalues spanning many decades — the “sloppy model” signature. Cited to ground the spectrum-based identifiability argument in §3.7.3 and §4.6 that supersedes any single condition-number cutoff.)

[43] Adler RA, El-Hajj Fuleihan G, Bauer DC, et al. Managing osteoporosis in patients on long-term bisphosphonate treatment: Report of a Task Force of the American Society for Bone and Mineral Research. J Bone Miner Res. 2016;31(1):16–35. doi:10.1002/jbmr.2918. PMID:26350171. (ASBMR 2016 task force consensus on long-term BP safety — atypical femoral fracture 1.78/1000 py at 5-10 y, GI intolerance ∼50-100/1000 py; cited in the §4.4.1 AE profile for alendronate/oral BP.)

[44] Khan AA, Morrison A, Hanley DA, et al. Diagnosis and management of osteonecrosis of the jaw: A systematic review and international consensus. J Bone Miner Res. 2015;30(1):3–23. doi:10.1002/jbmr.2405. PMID:25414052. (International consensus on ONJ incidence: 0.001-0.01/1000 py oral BP, 0.7-90/1000 py IV zoledronic; cited in §4.4.1 for the ONJ AE row.)

[45] Reid IR, Gamble GD, Mesenbrink P, Lakatos P, Black DM. Characterization of and risk factors for the acute-phase response after zoledronic acid. J Clin Endocrinol Metab. 2010;95(9):4380–4387. doi:10.1210/jc.2010-0597. PMID:20554713. (HORIZON-based analysis of IV zoledronic APR ∼300-400/1000 py first dose, tachyphylaxis on repeat; cited in §4.4.1 for the acute-phase-reaction row.)

[46] Watts NB, Diab DL. Long-term use of bisphosphonates in osteoporosis. J Clin Endocrinol Metab. 2010;95(4):1555–1565. doi:10.1210/jc.2009-1947. PMID:20173017. (Review of hypocalcemia risk 5-50/1000 py with BPs when Ca/vit-D repletion is inadequate; cited in §4.4.1 for the hypocalcaemia row.)

[47] U.S. Food and Drug Administration. FDA Drug Safety Communication: Safety update for osteoporosis drugs, bisphosphonates, and atypical fractures. October 13, 2010. https://www.fda.gov/drugs/postmarket-drug-safety-information-patients-and-providers/fda-drug-safety-communication-safety-update-osteoporosis-drugs-bisphosphonates-and-atypical (accessed 2026-07-16). (FDA 2010 label change; pregnancy Category C classification for all bisphosphonates due to skeletal half-life >10 y; cited in §4.4.1 for the pregnancy-contraindication row.)

[48] Pak CYC. Medical management of urinary stone disease. Nephron Clin Pract. 2004;98(2):c49–c53. doi:10.1159/000080252. PMID:15499207. (Pak 2004 review of K-Mg-citrate + K-citrate GI tolerability, hyperkalemia risk in renal insufficiency, and hypermagnesemia risk; cited in §4.4.1 for the K-Mg-citrate AE profile (GI/hyperK/hyperMg rows).)

