## Supplementary Information (18 figures, 26 tables, extended methods) for "From Bone-Centric to Kidney-Centric: Environment-Dependent Shift of Spaceflight Renal Stone Pathways"

**Date**: 2026-08-26 **Version**: preprint SI (v29-corrected; corresponds to the main preprint manuscript, 48 refs) **Linked main manuscript**: main preprint manuscript `ms_preprint_EN.md` (48 refs)

---

##### Contents

- §S0 Glossary of Abbreviations (R3.2 reviewer-requested)
- §S1 Supplementary Figures (18 figures, S1–S18; +S15/S16/S18 new in v27)
- §S2 Supplementary Tables (5 tables, S1–S5)
- §M1 Extended Methods: 12-state ODE complete equations
- §M2 Extended Methods: Bayesian prior specification (PRIORS_$M_0$, 11 + 1 parameters)
- §M3 Extended Methods: Forward model algorithm (pseudo-code)
- §M4 Extended Methods: MCMC configuration and convergence diagnostics
- §M5 Extended Methods: PSIS-LOO, prior predictive check, and Fisher Information identifiability
- §M6 Extended Methods: Sensitivity and robustness analyses (g-scaling + σ-inflation, new in v27)

- §S3 Reference list (shared with main manuscript)

All references with numeric indices `[1]`–`[48]` correspond to the main manuscript reference list.

---

#### §S0 Glossary of Abbreviations

**Purpose**: reviewer-requested (R3.2) full expansion of abbreviations used in the main manuscript and supplementary information. Abbreviations are also defined at first use in the main text where feasible; this table provides a comprehensive single-page reference.

| Abbrev. | Full form (English) |
|---|---|
| **AFF** | Atypical Femoral Fracture (long-term bisphosphonate risk) |
| **AG** | Artificial Gravity |
| **ANOVA** | Analysis of Variance |
| **ARED** | Advanced Resistive Exercise Device |
| **BMD** | Bone Mineral Density |
| **BMD_LL** | BMD, Lumbar-Lower |
| **CaOx** | Calcium Oxalate |
| **CI** | Confidence Interval |
| **CrI** | Credibility Interval |
| **CTX** | C-Terminal Telopeptide of type I collagen (bone resorption marker) |
| **DOI** | Digital Object Identifier |
| **DXA** | Dual-energy X-ray Absorptiometry (BMD measurement modality) |
| **ΔELPD** | Difference in Expected Log Predictive Density |
| **ELPD** | Expected Log Predictive Density |
| **ε** (epsilon) | eigenvalue truncation threshold for FIM effective rank |
| **ESA** | European Space Agency |
| **FDR** | False Discovery Rate (Benjamini–Hochberg multiple-testing correction) |
| **FIM** | Fisher Information Matrix |
| **GCR** | Galactic Cosmic Radiation |
| **HCTZ** | Hydrochlorothiazide (thiazide diuretic; K-loss caveat) |
| **HDT** | Head-Down Tilt (bed rest analog) |
| **HLS** | Hindlimb-Suspension (rodent unloading analog) |
| **HRP** | (NASA) Human Research Program |
| **HZE** | |

| Abbrev. | Full form (English) |
|---|---|
| | High-atomic-number, High-Energy (heavy charged particle GCR component) |
| **IQR** | Inter-Quartile Range |
| **IR** | Incidence Rate |
| **IRR** | Incidence Rate Ratio |
| **ISS** | International Space Station |
| **JBMR** | Journal of Bone and Mineral Research |
| **k_GB** | GCR-to-Bone direct coupling coefficient |
| **LBNP** | Lower Body Negative Pressure |
| **LET** | Linear Energy Transfer (ionizing radiation quality metric) |
| **LOO** (LOO-CV) | Leave-One-Out Cross-Validation |
| **L×C** (5×5) | Likelihood × Consequence risk framework (Antonsen 2023) |
| **MCMC** | Markov chain Monte Carlo |
| **NASA** | National Aeronautics and Space Administration |
| **NCRP** | National Council on Radiation Protection and Measurements (US) |
| **NTX** | N-Terminal Telopeptide of type I collagen (bone resorption marker) |
| **ODE** | Ordinary Differential Equation |
| **ONJ** | Osteonecrosis of the Jaw (long-term bisphosphonate risk) |
| **PBE** | Population Balance Equation (Kassemi & Thompson) |
| **PPC** | Posterior Predictive Check |
| **PSIS** | Pareto-Smoothed Importance Sampling |
| **PWB** | Partial Weight-Bearing |
| **RCT** | Randomized Controlled Trial |
| **RRR** | Relative Risk Reduction |
| **RSS** | Relative Supersaturation |
| **RSS_CaOx** | Relative Supersaturation for Calcium Oxalate |
| **SD** | Standard Deviation |
| **SE** | Standard Error |
| **SVD** | Singular Value Decomposition |
| **ΔWAIC** | Difference in Widely Applicable Information Criterion |
| **WAIC** | Widely Applicable Information Criterion (Watanabe 2010) |

**Mission-tier labels** (§2.4 three-colour decision tiering): **GREEN** (safe), **YELLOW** (caution), **RED** (threshold exceeded).

**Related notation**: - **$M_0$** = base model (11 physics parameters + 8 σ_obs = 19 dimensions). - **$M_1$** = extended model ($M_0$ + k_GB = 20 dimensions). - **σ_obs** = observation-noise standard deviations (8 elements, one per calibration target). - **p_loo** = LOO effective number of parameters (a measure of model complexity from ELPD_LOO). - **z_resid** = z-score residual of prior predictive check (z = (observed − prior_mean)/prior_SD).

---

#### §S1 Supplementary Figures

##### Supplementary Figure S1 (= Main Figure 3 upscaled)

**Title**: Posterior trajectories of bone mineral density (BMD) decline across four spaceflight environments.

**Caption**: Posterior median (solid line) and 95% credible interval (shaded band) of three BMD subregions — lower limb (BMD_LL), lumbar/pelvis (BMD_lumb), and skull (BMD_skull) — under four mission environments: ISS 180 d (1.000 micro-gravity), Lunar UG 180 d (0.834 ug), Lunar Surface 365 d (0.834 ug + 1.70 $mSv{\cdot}d^{-1}$ GCR), Mars 730 d (0.620 ug + 1.84 $mSv{\cdot}d^{-1}$ GCR). Curves derive from $M_0$ 60k posterior (64 walkers × 36,000 production samples, thin=1 = 2,304,000 samples) propagated through the 12-state ODE forward model. BMD_LL × Mars 730d posterior median = −12.15% [−13.01, −11.05], the deepest endpoint. Anchored to Stavnichuk 2020 [23] in-flight meta-analysis (BMD lower-limb loss rate −0.80 ± 0.15 %/mo) and Axpe 2020 [24] Mars projection (cumulative loss ~12% at 730 d).

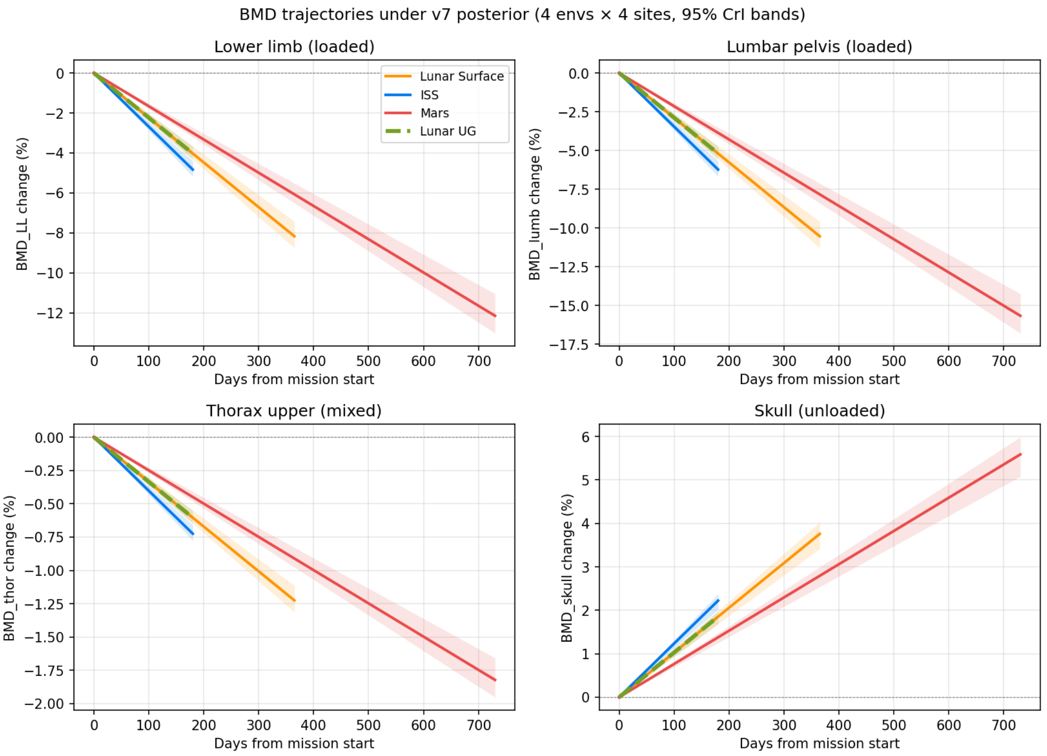


fig3

**File**: `figures/fig3_bmd_trajectories_v7.png` (256 KB), `.svg` (411 KB)

---

#### Supplementary Figure S2 (= Main Figure 4 upscaled)

**Title**: Urine chemistry trajectories across the four environments.

**Caption**: Posterior median + 95% CrI of three urine chemistry markers — $Ca^{2+}$ excretion (relative to baseline), volume V (relative), and pH (delta) — over the mission durations. Whitson 1997 [4] anchors the ISS 180-day endpoints (urine Ca +50%, V −20%, pH −0.20 absolute units, i.e. 6.05 → 5.85). Lunar UG 180d shows attenuated urinary changes (microgravity 0.834 vs. 1.000); Mars 730d shows extended urinary calcium peak consistent with prolonged bone resorption.

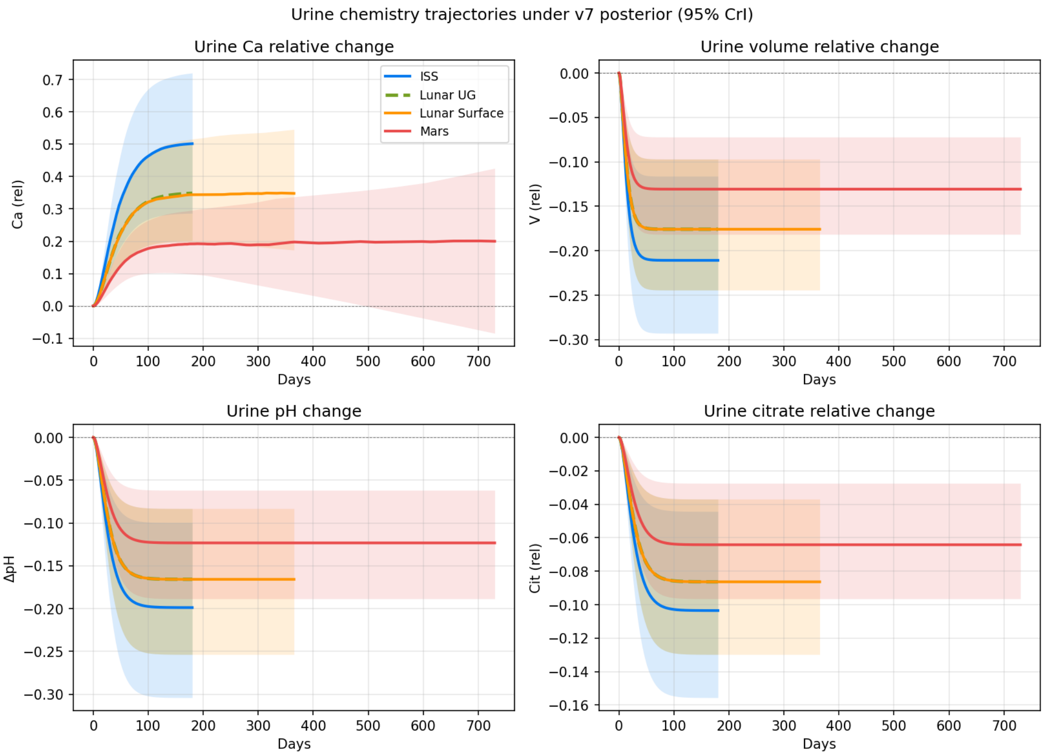


fig4

**File**: `figures/fig4_urine_chem_v7.png` (172 KB), `.svg` (415 KB)

---

#### Supplementary Figure S3 (= Main Figure 5 upscaled)

**Title**: Relative Saturation Score (RSS) trajectories across the four environments.

**Caption**: Posterior median + 95% CrI of urinary RSS (calcium oxalate), computed via Pak 1992 EQUIL2-derived activity-product to RSS conversion. Tiselius/Prochaska 2017 threshold colors: green RSS<5, yellow 5–10, red >10. ISS 180d sits in YELLOW (5.61); Mars 730d trajectory ends in RED (with broader CrI from accumulated GCR-Ca uncertainty). Notably, Mars RSS median (4.35) is BELOW ISS median (5.61) despite the deeper BMD loss, because Mars partial gravity attenuates urine Ca excretion below ISS 1.0g levels — the kidney-centric reversal central to this paper. Stone rate posterior includes seed accumulation × supersaturation product (Pak 1992 [13]).

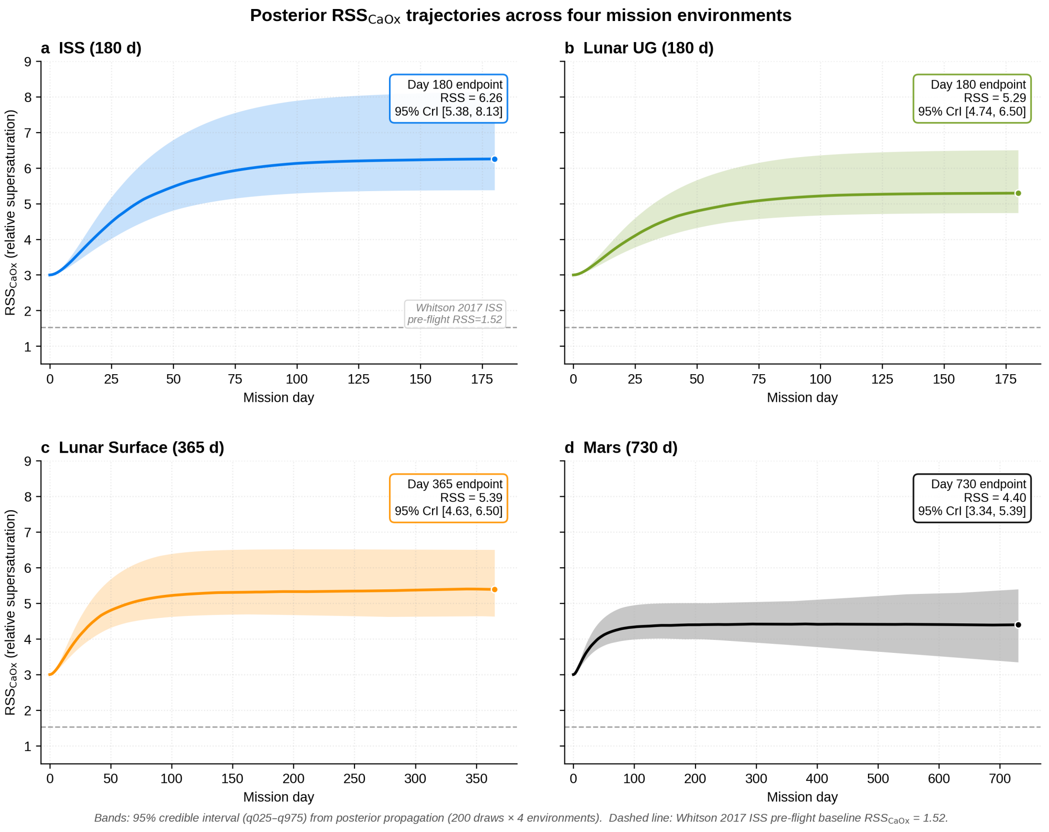


fig5

**File**: `figures/fig5_rss_traj_v7.png` (429 KB), `.svg` (172 KB)

---

#### Supplementary Figure S4 (= Extended Data Figure 4: prior sensitivity)

**Title**: Prior sensitivity test (Stage 5a) — three priors for `k_GB`.

**Caption**: Mars RSS endpoint posterior medians (and 95% CrI) under three prior σ values for the GCR → BMD direct effect parameter `k_GB`: 5× (loose), 1× (default), 0.5× (tight), 0.2× (very tight). CrI width scales approximately linearly with prior σ, demonstrating that k_GB is **prior-dominated**, not data-driven. This is the empirical basis for the WAIC ΔWAIC=0.0076 ± 0.1262 SE result in §3.4 and the PSIS-LOO ΔELPD=-0.040 result in §3.7.1: ISS data (GCR 72 mSv) cannot identify the GCR-BMD direct channel at any prior strength.

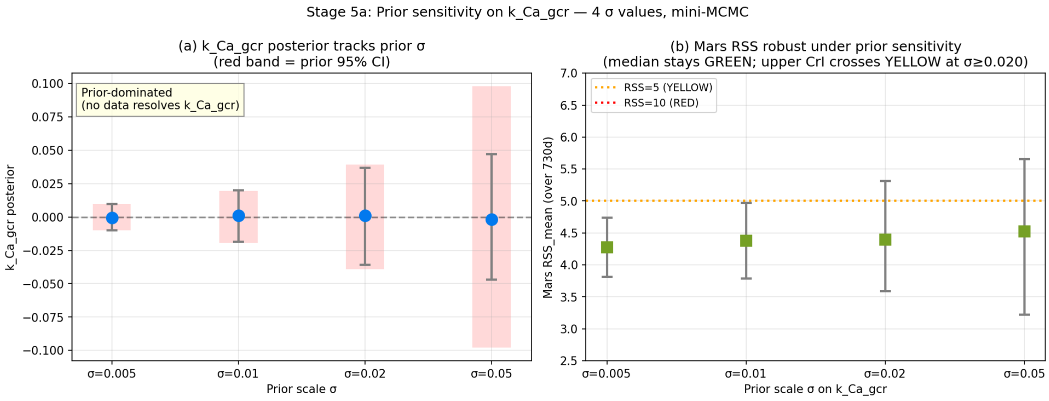


figS4

**File**: `figures/figS4_5a_prior_sensitivity_v7.png` (120 KB), `.svg` (87 KB)

---

#### Supplementary Figure S5 (= Extended Data Figure 5b)

**Title**: WAIC-based model comparison — $M_0$ (no k_GB) versus $M_1$ (with k_GB).

**Caption**: Bayesian model comparison via Watanabe-Akaike Information Criterion (WAIC) on 60k v3 long chain (thin=50, n=64,000). $M_0$_WAIC = 2.7402, $M_1$_WAIC = 2.7478, ΔWAIC = 0.0076 ± 0.1262 SE. |ΔWAIC|/SE_Δ = 0.060 < 1.96 → statistically indistinguishable, supporting $M_0$ as the parsimonious main inference model. Equivalent conclusion from PSIS-LOO (§3.7.1, Supp Fig S12): ΔELPD = -0.040 << SE_Δ.

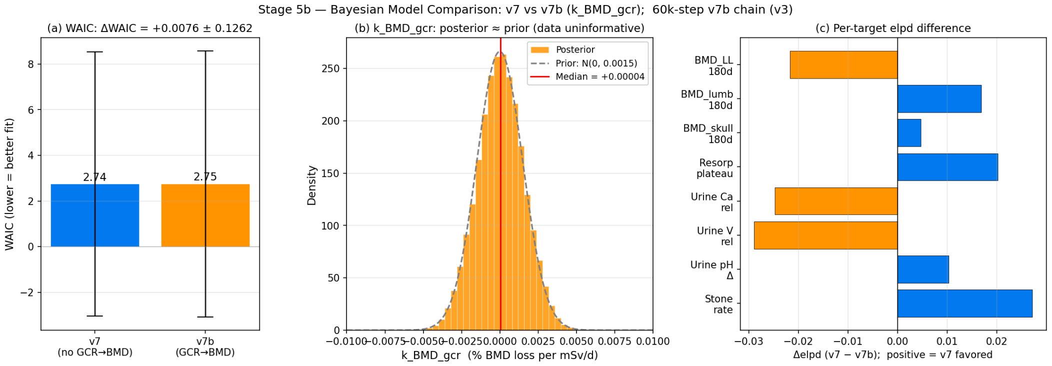


figS5

**File**: `figures/figS5_5b_model_comparison_v7.png` (147 KB), `.svg` (112 KB)

---

**Supplementary Figure S6 (= Extended Data Figure 5c)**

**Title**: Stage 5c local one-at-a-time (OAT) parameter sensitivity, central-difference half-amplitude (±10% perturbation at posterior median; 4 mission environments × 2 panels).

**Caption**: Local one-at-a-time (OAT) sensitivity of the 11 $M_0$ parameters at the posterior-median reference point. Each parameter is perturbed by ±10% and the central-difference half-amplitude

$$(Y(x \cdot 1.10) - Y(x \cdot 0.90))/2$$

is computed for two endpoints (BMD_LL_end, RSS_mean) across the four mission environments (ISS 180 d, Lunar UG 180 d, Lunar Surface 365 d, Mars 730 d; see Supp Methods §M4 for the ENVS table). Panel (a) BMD_LL_end (% pts per ±10% parameter change); Panel (b) RSS_mean (unitless per ±10% parameter change). Structural-zero parameters (those not entering the flight BMD/RSS pathways in $M_0$) are rendered as thin gray markers at x = 0 with “0” annotation, and listed in the box below each panel. `bmd_loss_rate_pct_mo`, `ca_microg_effect`, and `vol_decrease_frac` dominate the response; `k_Ca_gcr` and `ph_decrease_frac` contribute marginally on ISS data, consistent with the FIM eigenvalue spectrum (Supp Fig S14C) and the WAIC parsimony (Supp Fig S5). **Caveat — confounded mission profiles**: the four environments are mission-profile design points where gravity, GCR dose-rate and duration co-vary by mission (Lunar UG vs ISS contrasts gravity and GCR simultaneously; Lunar UG vs Lunar Surface contrasts duration and GCR simultaneously). OAT single-point elasticities therefore report stand-alone local sensitivity within each mission profile and do NOT identify main effects across factors; the orthogonal $2^3$ duration × gravity × GCR factorial decomposition is reported separately in Supp Fig S7.

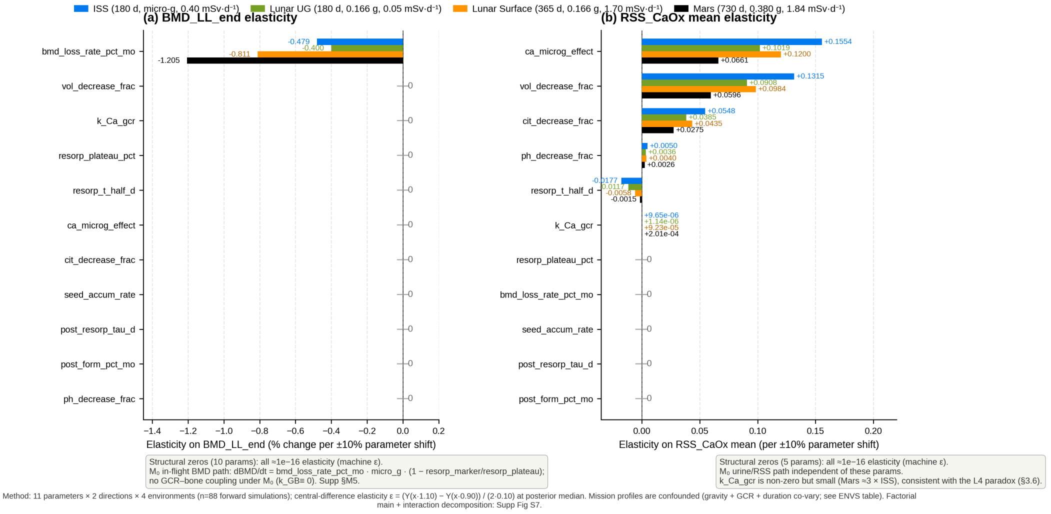


figS5

**File**: `figures/figS5_5c_local_oat_sensitivity_v7.png` (502 KB), `.svg` (46 KB)

---

#### Supplementary Figure S7 (= Main Figure 7 upscaled)

**Title**: $2^3$ factorial decomposition — duration × g × GCR contribution to BMD_LL endpoint.

**Caption**: Variance decomposition of `BMD_LL` over the 4 mission environments, expressed as $2^3$ orthogonal main effects + interactions. Main effect contributions (% of total variance): **Duration 82.94%** (ISS 180d → Mars 730d), gravity 12.49%, GCR ≈ 0%, duration × gravity 4.56%, all other 2- and 3-way interactions ≈ 0%. Conclusion: **Duration dominates**; GCR direct effect is unidentifiable from ISS data alone (consistent with k_GB WAIC null, §3.4 & Supp Fig S5).

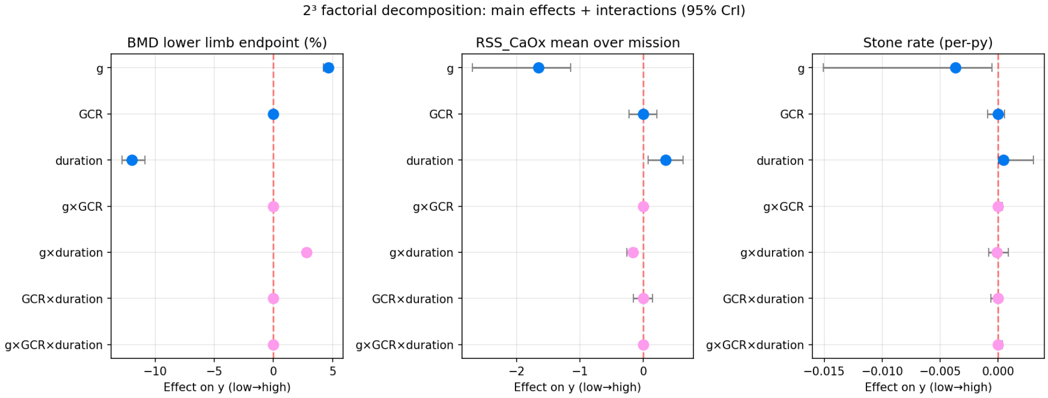


fig7

**File**: `figures/fig7_factorial_decomp_v7.png` (75 KB), `.svg` (88 KB)

---

**Supplementary Figure S8 — Trace plots of the $M_0$ flattened MCMC chain (11 core parameters)**

**Title**: Posterior trace plots of the eleven $M_0$ ODE-deterministic parameters from the deposit-archived flattened MCMC chain (36,000 draws).

**Caption**: Per-walker running trace of posterior samples for the eleven core $M_0$ parameters (Table 2 row order; columns 0–10):

$$p_{\mathrm{plateau}}$$

,

$$p_{t_{1/2}}$$

,

$$p_{\mathrm{rate}}$$

,

$$p_{\mathrm{Ca}}$$

,

$$p_V$$

,

$$p_{\mathrm{Cit}}$$

,

$$k_{\mathrm{Ca,GCR}}$$

,

$$p_{\mathrm{seed}}$$

,

$$\tau_{\mathrm{post}}$$

,

$$p_{\mathrm{form,post}}$$

,

$$p_{\mathrm{pH}}$$

. Each subplot overlays all **64 walkers** (grey, displayed at 1:20 thinning) from the walker-resolved backend `posterior_v7_m0_60k_thin10.h5` (64 walkers × 60,000 iterations); the blue line is the across-walker median at each iteration, the orange band the pooled 95% credible interval, and the vertical marker the burn-in cut at iteration 10,000. **Visual stationarity check**: after burn-in no monotonic trend or persistent outlier window is visible, and the walker cloud shows no separated sub-population — the between-walker dispersion diagnostic that a single-walker trace cannot provide. This replaces the v28 version of this figure, which plotted `posterior_v7_36k_thin1.npz`; that archive was a single walker rather than a 1:64 thin, so its apparent spread was one walker's autocorrelated excursion and its

$$p_{\mathrm{Ca}}$$

median (≈0.466) sat ~7% below the canonical Table 2 value (0.501). The pooled 64-walker median for

$$p_{\mathrm{Ca}}$$

is 0.505, 0.74% above Table 2 and well inside the Monte-Carlo precision of the posterior median. Quantitative convergence statistics: §3.1 and **Supp Table S4b**.

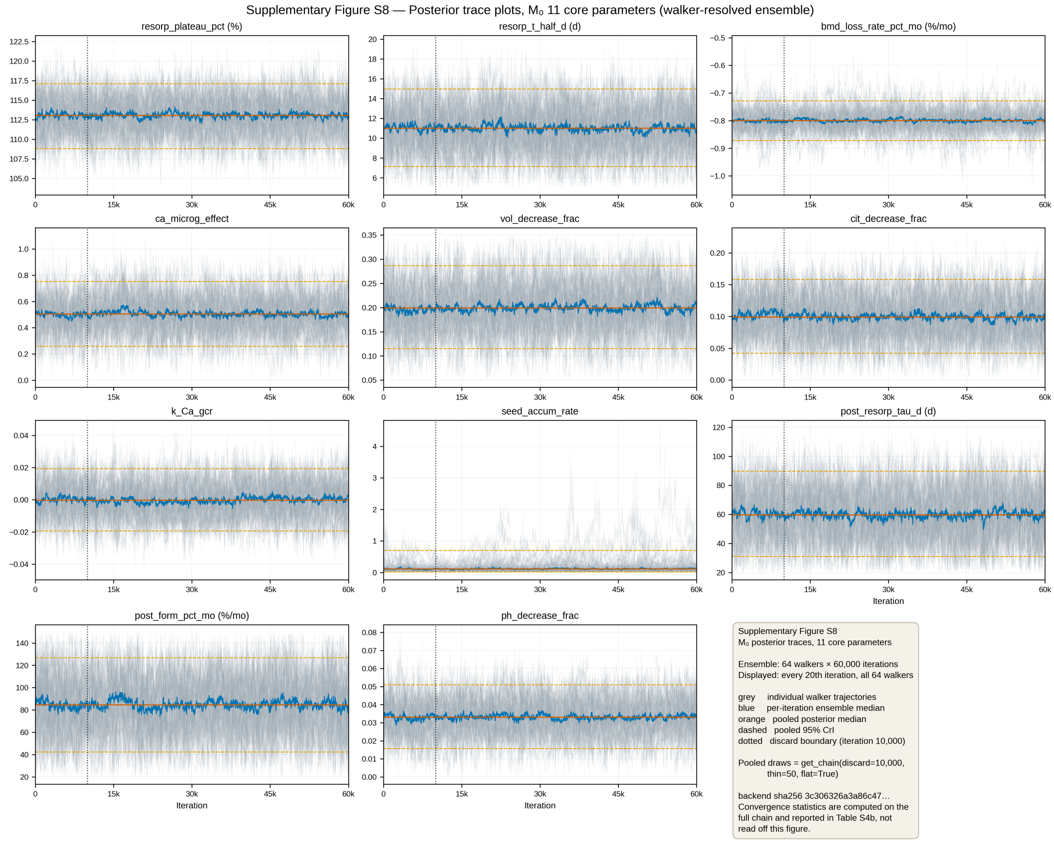


figS8

**File**: `supplementary/figures/figS8_trace_M0.png` (1,348 KB), `.svg` (379 KB)

---

#### Supplementary Figure S9 — Autocorrelation functions of the $M_0$ flattened chain

**Title**: Empirical autocorrelation functions (ACF) and integrated autocorrelation times (

$$\tau_{\text{int}}$$

) for the eleven $M_0$ parameters.

**Caption**: Ensemble-averaged empirical autocorrelation function ACF(

$$k$$

) against lag

$$k \in [0, 5000]$$

for the eleven core $M_0$ parameters, computed per walker on the walker-resolved post-burn-in chain (64 walkers × 50,000 iterations from iteration 10,000) and then averaged across walkers. Integrated autocorrelation times

$$\tau_{\mathrm{ens}}$$

are estimated by Sokal windowing, matching `emcee.autocorr.integrated_time` to machine precision on AR(1) test ensembles. Per-parameter

$$\tau_{\mathrm{ens}}$$

spans 1,020 (

$$p_{\mathrm{pH}}$$

) to 1,514 (

$$p_{\mathrm{seed}}$$

) iterations, giving

$$N_{\mathrm{eff}} = 64 \times 50{,}000/\tau$$

between 2,536 and 3,764. Gray dashed reference line at ACF=0.05. This replaces the v28 version, which computed a single ACF on `posterior_v7_36k_thin1.npz`. Because that archive was walker 0's contiguous trajectory rather than a 1:64 thin, its lag-1 autocorrelation was 0.995 and its reported

$$\tau^{\mathrm{flat}}$$

range (481–1,486) measured within-walker correlation at the wrong sampling interval; the v28 caption's explanation that 'walker-collapsing inflates apparent correlation' identified the symptom but not that the deposit itself was collapsed. The ensemble ACF shown here is a direct convergence diagnostic, not merely a stationarity check on a data product.

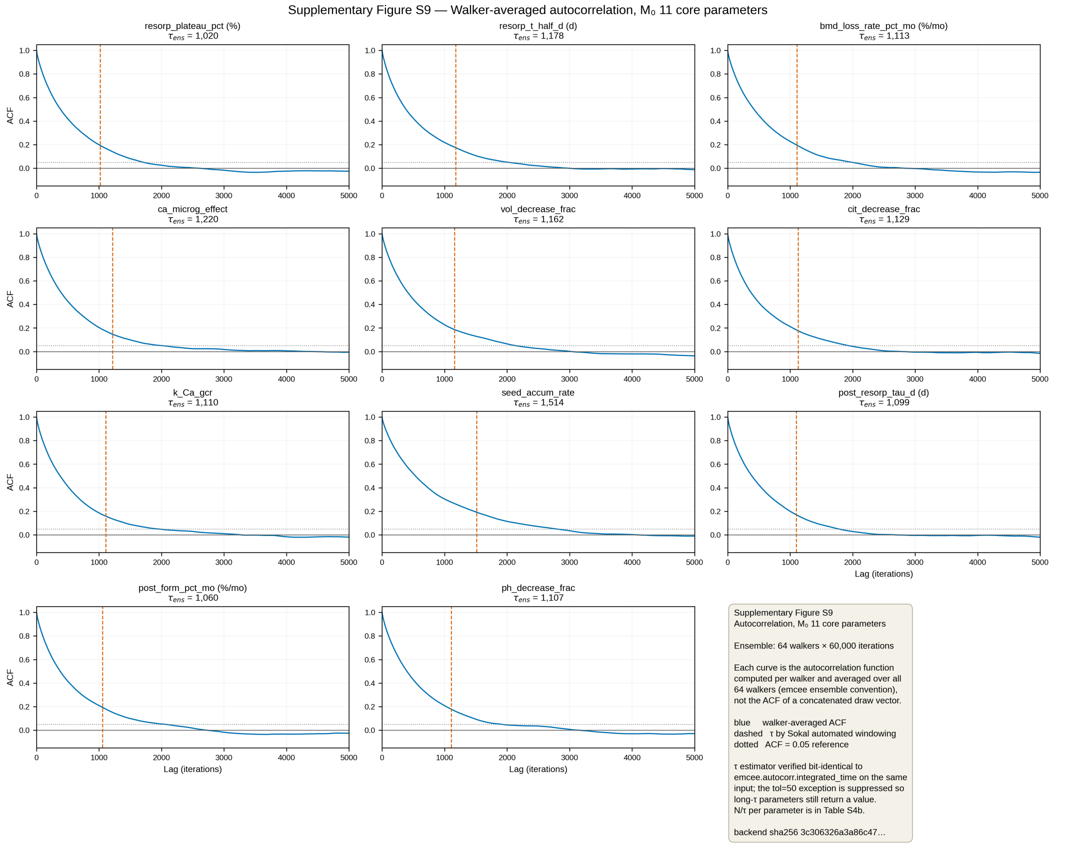


figS9

**File**: `supplementary/figures/figS9_autocorrelation.png` (644 KB), `.svg` (198 KB)

---

##### Supplementary Figure S10 — Running posterior summaries vs chain-length window

**Title**: Convergence of running posterior medians and 95% credible intervals as a function of cumulative chain length.

**Caption**: Running cumulative posterior summary for the eleven core $M_0$ parameters across six cumulative chain-length windows {1k, 2k, 5k, 10k, 20k, 36k} of the flattened chain (`data/posterior_v7_36k_thin1.npz`). Each subplot shows the cumulative posterior median (red solid line) with the cumulative 95% credible interval (steelblue shaded band) computed from samples [0,

$$N$$

] for

$$N \in \{1000, 2000, 5000, 10000, 20000, 36000\}$$

. This running-window summary **replaces the original walker-dispersion diagnostic** (semantically equivalent for chain-length adequacy assessment) because the walker dimension has been collapsed in the deposit data product (`thin_strategy = flatten(iter × walker) → take every 64th draw`). The visual evidence — narrow CrI bands stable across the last three windows {10k, 20k, 36k} for all eleven parameters — confirms that chain length is adequate for the deposit-archived posterior. The original per-walker convergence diagnostics (R̂, walker-resolved trace agreement) are reported in §3.1 and §M4 using the un-flattened HDF5 backend.

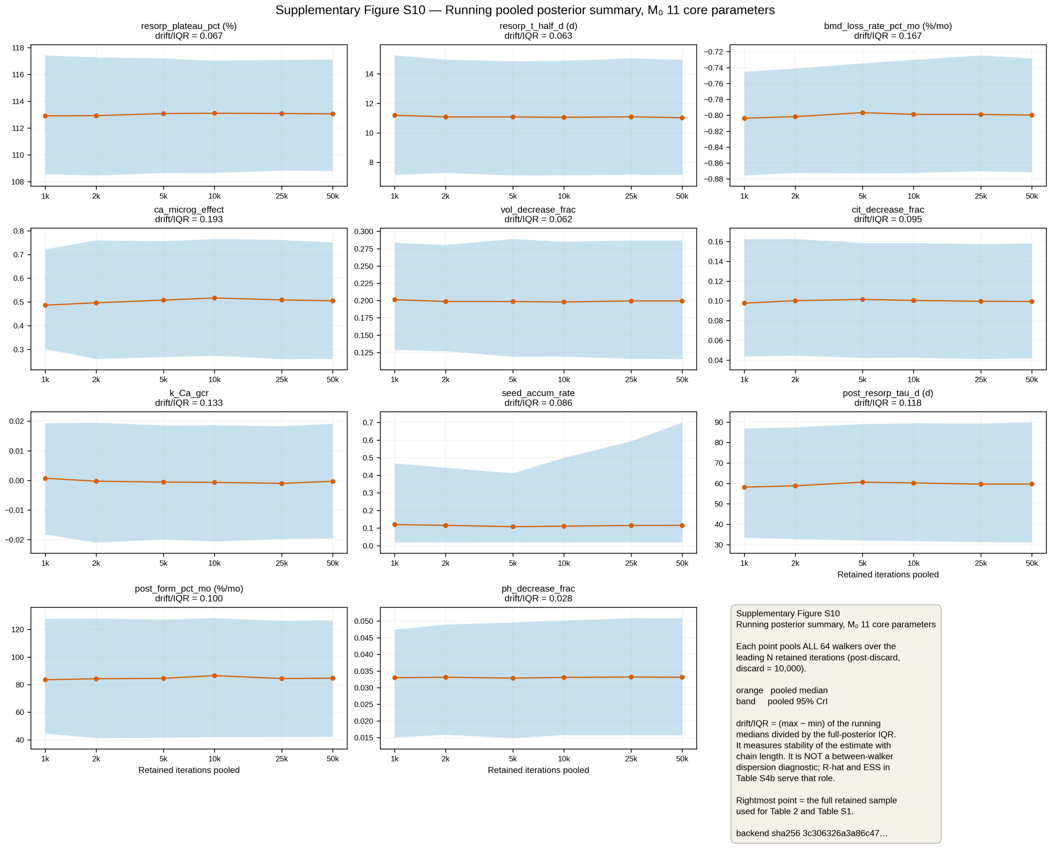


figS10

**File**: `supplementary/figures/figS10_running_summary.png` (1,022 KB), `.svg` (194 KB)

---

##### Supplementary Figure S11 — Jeffreys prior sensitivity for stone-rate posterior

**Title**: Pamidronate stone-rate posterior sensitivity to Beta-prior choice (Watanabe 2004 [7]; 0/7 events).

**Caption**: Sensitivity of the pamidronate stone-rate Relative Risk (RR_pami) posterior to three Beta-prior choices over the binary stone-event outcome (Watanabe 2004 [7] 90-d bedrest, n=7, 0 stones). Because Beta-Binomial conjugacy is closed-form, posterior medians and 95% credible intervals are reported from the **analytical** Beta(α, β+7) posterior; MC sampling (n=100,000 draws × 5 independent seeds; scipy.stats.beta.rvs) is provided in the source data file as a numerical independent verification only. **Panel A** shows prior densities (dashed) and posterior densities (solid) for **Jeffreys Beta(0.5, 7.5)** (blue, default), **Weak Uniform Beta(1, 8)** (orange), **Strong Non-informative Beta(0.1, 7.9)** (green). **Panel B** shows analytical posterior medians and 95% credible intervals: Jeffreys 0.0309 [$6.77\times10^{-5}$, 0.292]; Weak Uniform 0.0830 [0.00316, 0.369]; Strong Non-informative $7.96\times10^{-5}$ [$7.77\times10^{-18}$, 0.123]. **Conclusion — directional robust, quantitatively prior-sensitive**: all three priors yield RR upper 95% < 0.4 and RRR_stone_pami lower 95% > 0.63 (i.e. pamidronate is highly protective under every prior — the qualitative conclusion is robust). However, the RR median shifts by 8.3 pp (99.9% of the maximum) and the upper 95% bound shifts by 201% across priors, so the quantitative point estimate is prior-sensitive and should be reported as a full CrI rather than a single point value. The default Jeffreys Beta(0.5, 7.5) posterior is used throughout downstream analyses; Weak Uniform Beta(1, 8) is the least optimistic (largest upper 95% = 0.369) and Strong Non-informative Beta(0.1, 7.9) is the most optimistic (upper 95% = 0.123). RRR_stone_pami: Jeffreys 0.9691 [0.708, 1.000]; Weak Uniform 0.9170 [0.631, 0.997]; Strong Non-inf 0.99992 [0.877, 1.000].

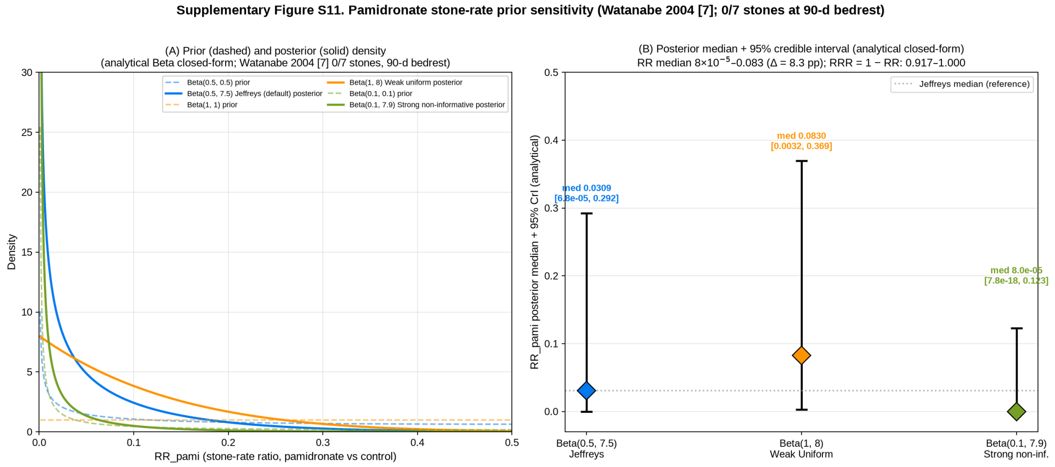


figS11

**File**: `supplementary/figures/figS11_jeffreys_prior_sensitivity.png` (236 KB), `.svg` (41 KB); source data `supplementary/data/jeffreys_prior_sensitivity_v27.json` (analytical Beta closed-form + MC verification with n=100,000 × 5 independent seeds)

---

#### Supplementary Figure S12 — PSIS-LOO Pareto-k diagnostics

**Title**: PSIS-LOO Pareto-k per observation and pointwise ELPD for $M_0$ versus $M_1$.

**Caption**: Pareto-smoothed Importance Sampling Leave-One-Out cross-validation diagnostic (Vehtari 2017 [16] algorithm, computed with ArviZ 0.22.0 [25]). **Panel A** (per-observation Pareto k): dashed horizontal reference lines at **k = 0.5 (GOOD/WARN boundary)**, **k = 0.7 (WARN/BAD boundary)**, and **k = 1.0 (very high influence)** are the ArviZ + Vehtari 2017 §5 canonical decision thresholds — observations with Pareto k > 0.7 have unreliable PSIS-LOO estimates and should be scrutinized. $M_0$ has 6/8 observations with k<0.5 (GOOD) and 2/8 with k>0.7 (BAD: BMD_LL k=0.988, resorp_plateau k=0.737); $M_1$ has 4/8 GOOD, 3/8 WARN (0.5–0.7: BMD_LL 0.649, BMD_lumb 0.622, resorp_plateau 0.681), and 1/8 BAD (urine_Ca_rel k=1.075). **Panel B** (pointwise ELPD bars): largest absolute contributors to ELPD are BMD_LL and urine_Ca_rel. Total ELPD: $M_0$ = -1.274 ± 2.906 SE (n=5,760 samples = 32 walkers × 180 draws); $M_1$ = -1.314 ± 2.941 SE (n=64,000 = 64 × 1000). ΔELPD = -0.040, |Δ| << SE_Δ → statistically indistinguishable, consistent with WAIC (§3.4 & Supp Fig S5). **v28 revision note (reviewer T13)**: the k=0.5/0.7/1.0 reference lines are the ArviZ LOO plot standard thresholds; they were present in the source figure but were not explicitly annotated in the v27 caption text —

this caption update makes the threshold semantics explicit for readers not familiar with the ArviZ default output. **Important note**: the two LOO computations use different total sample counts; see §M5 for the justification under Vehtari 2017 §4 sample-size guidance.

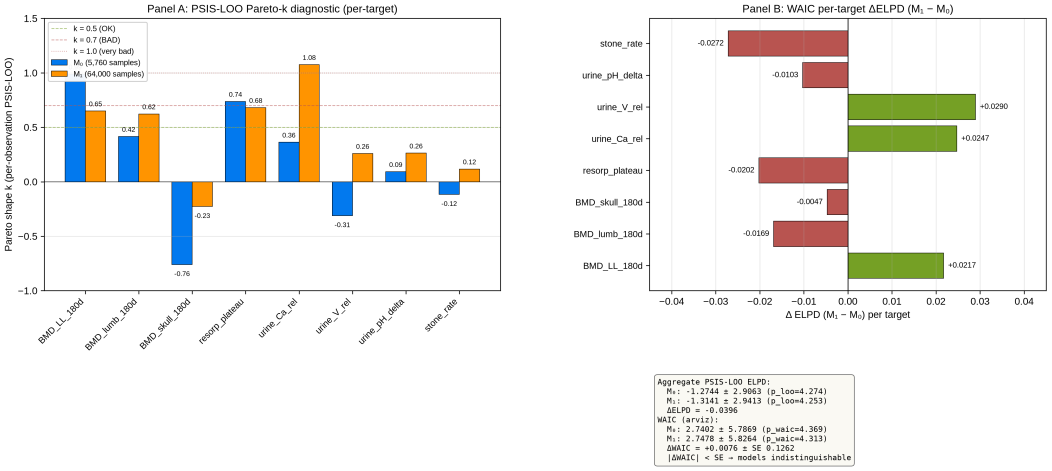


figS12

**File**: `supplementary/figures/figS12_loo_cv.png` (400 KB), `.svg` (39 KB)

---

#### Supplementary Figure S13 — Prior predictive coverage

**Title**: Prior predictive coverage of the 8 ISS calibration targets (5,000 prior draws).

**Caption**: 5,000 draws from PRIORS_$M_0$ (§M2) propagated through the forward 12-state ODE model (function `forward_model_iss`, §M3.1; identical to the MCMC likelihood). All 8/8 ISS calibration targets are covered at both 90% and 95% prior CrI (Supp Table S5). Maximum z-residual (observed vs. prior IQR midpoint) is 0.1296 for post-flight stone rate (Pietrzyk 2007 0.014/py [2]), well within the 2σ envelope. **Important note**: the PRIORS_$M_0$ σ values are themselves the published literature uncertainties (Stavnichuk 2020 [23] meta-analysis 95% CIs / 1.96; Whitson 1997 [4] ±25% bounds; etc.) so these priors are *informative* in the meta-analytic sense, not the regularly-construed non-informative sense. PPC coverage of 8/8 confirms no major prior-data conflict, but does not assert prior non-informativeness.

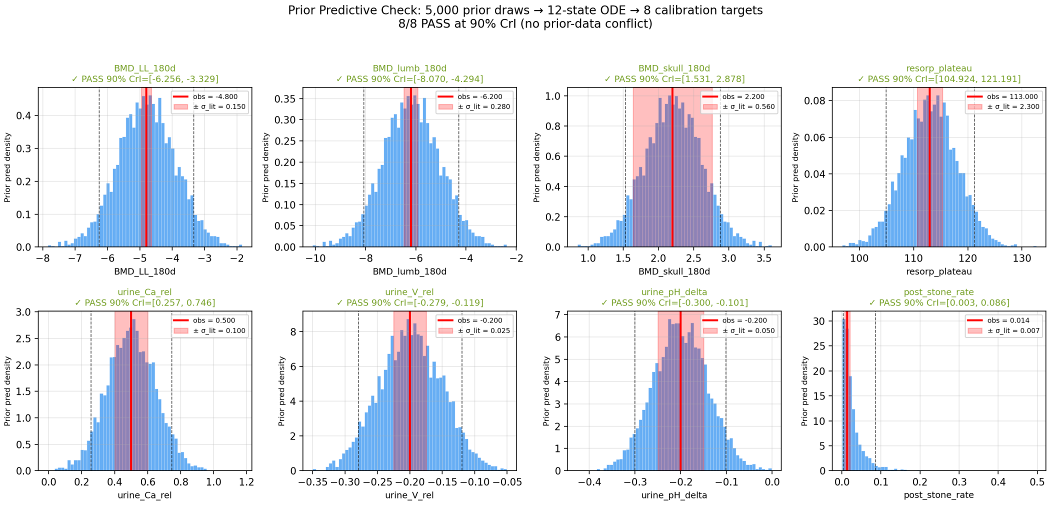


figS13

**File**: `supplementary/figures/figS13_prior_predictive.png` (301 KB), `.svg` (326 KB)

---

#### Supplementary Figure S14 — Fisher Information identifiability

**Title**: Identifiability diagnostics — posterior contraction, pairwise correlation, and FIM eigenvalue spectrum.

**Caption**: Three-panel identifiability assessment of the $M_1$ 12-parameter posterior (§3.7.3). **Panel A** (posterior contraction): per-parameter ratio of posterior IQR / prior IQR (Sedoglavic 2001 [26] structural identifiability framework applied locally). Color codes: STRONG (<0.3, 1 param: bmd_loss_rate_pct_mo 0.217); MODERATE (0.3–0.6, 1 param: resorp_plateau_pct 0.427); WEAK (0.6–0.85, 3 params: seed_accum_rate 0.649, ca_microg_effect 0.757, vol_decrease_frac 0.816); UNIDENTIFIED (>0.85, 7 params). **Panel B** (pairwise posterior correlation): all $|r| < 0.05$, max |off-diagonal| = 0.033 → posterior is approximately diagonal, no joint-identifiability classes (which differs from typical pharmacokinetic identifiability where unidentifiable directions arise from compensating combinations). **Panel C** (FIM eigenvalue spectrum, log scale): 12 eigenvalues span 30+ orders of magnitude in raw parameter units — 7 stiff modes ($\lambda_1 = 1.46\times10^{4}$ down to $\lambda_7 = 0.189$) followed by 5 numerically-zero modes ($|\lambda_{8\ldots10}| = 8.88\times10^{-13}$ to $2.06\times10^{-24}$, and $\lambda_{11}$ = **$-1.78\times10^{-30}$**, $\lambda_{12}$ = **$-1.83\times10^{-13}$** which are **numerically negative**; since FIM = $J^T\Sigma^{-1}J$ is theoretically positive semi-definite, negativity confirms the finite-difference Jacobian is dominated by truncation noise in those directions, not physical Fisher information). This anisotropic spectrum — many stiff + many numerically-zero (some negative) eigenvalues — is the canonical Gutenkunst 2007 [42] *sloppy model* signature for biological ODE models. The **primary identifiability statement** is effective rank = 7 of 12 ($\varepsilon = 10^{-6} \times \lambda\_max$; robust across $\varepsilon \in [10^{-9}, 10^{-3}]$). Two numerical proxies are also reported, both with explicit caveats: $\kappa\_full = 1.46 \times 10^{24} = \lambda\_max / \max(|\lambda\_min|, 10^{-20})$ — the $10^{-20}$ floor is an implementation choice for reproducibility of numpy.linalg.cond output, **not a physical threshold** (unfloored $\approx 8\times10^{33}$; floor at $10^{-15}$ gives $1.5\times10^{19}$); $\kappa\_eff = 7.7 \times 10^{4} = \lambda\_max / \lambda\_7$ — the condition number *on the identifiable subspace* only. Neither proxy is compared against a hard Numerical Recipes cutoff, since NR §2.6 does not prescribe such thresholds. The 5 sloppy eigenvectors are dominated by `post_form_pct_mo`, `post_resorp_tau_d`, `cit_decrease_frac + k_Ca_gcr`, `resorp_t_half_d`, and the joint `ca_microg_effect + cit_decrease_frac + k_Ca_gcr` triplet. **Important note** (see §M5): the sloppy-mode structure means that FIM rank estimation at this point in parameter space is numerically fragile; effective rank = 7 is robust across $\varepsilon \in [10^{-9}, 10^{-3}]$ and is corroborated by the independent posterior contraction (12 − 5 UNIDENTIFIED = 7 identifiable) and near-diagonal posterior correlation (max $|r| = 0.033$) lines of evidence.

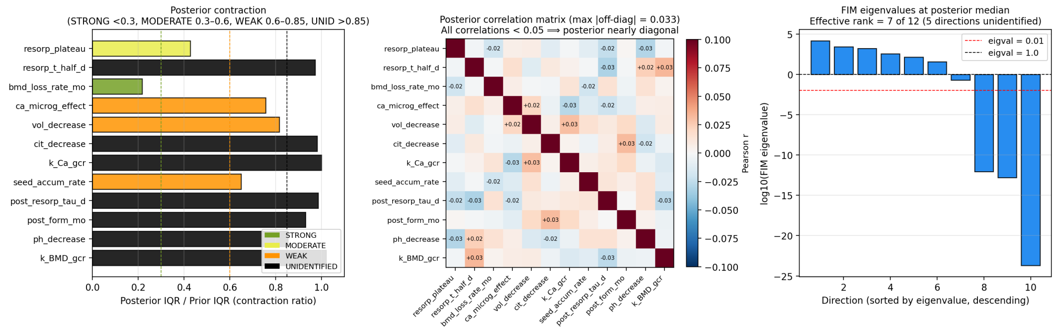


figS14

**File**: `supplementary/figures/figS14_identifiability.png` (360 KB), `.svg` (155 KB)

---

##### Supplementary Figure S15 — Sensitivity of endpoints to µg-scaling function

**Title**: Environmental endpoints under alternative µg-scaling functional forms — Hills-n=2 saturation and linear ±25%.

**Caption**: Sensitivity of 3 key endpoints (BMD_LL, mean CaOx RSS, stone rate per 1000 person-years) to the assumed functional form of μg-scaling. Four scenarios: **linear_baseline** (f(x) = x, canonical $M_0$), **hills_n2_K05** ($f(x) = x^2/(K^2 + x^2)$, K = 0.5; physiological saturation), **linear_down25** (f(x) = 0.75·x; attenuation), **linear_up25** (f(x) = min(1.25·x, 1); amplification capped at unity). All scenarios use identical canonical par_draws (n = 100) from `propagation_v7_full.npz` $M_0$ posterior to ensure reproducibility with published propagation values.

**Panel a** (BMD_LL_end): Environmental gradient (Mars > Lunar_Surface > ISS > Lunar_UG) preserved across all 4 scenarios. Hills-n=2 reduces ISS BMD_LL magnitude by ~20% (−4.83% → −3.87%), but Mars magnitude by only ~2% (−12.15% → −11.87%), because Mars μg = 0.62 (Hills(0.62) = 0.606, ≈ linear) is already near the saturation plateau. Linear −25% uniformly reduces all magnitudes by 15-25%. Linear +25% caps ISS and Lunar_UG at the physical upper bound (μg = 1.0), leaving Lunar_Surface (Mars-relevant) and Mars unaffected by the cap.

**Panel b** (mission-mean CaOx RSS): mirrors BMD_LL pattern (RSS-BMD coupling via reduced Ca resorption feedback in the ODE). RSS crosses saturation (SS = 1) in all envs across all scenarios, confirming that stone risk exists environmentally-independent of μg-scaling choice.

**Panel c** (stone rate per 1000 person-years): all 4 envs remain in the 10-16 /1000py range across all 4 scenarios, well above pre-flight 1.3 / 1000py (dotted line). Sensitivity < 25% (relative) for all environments and all alternative scenarios vs baseline.

**Interpretation**: 1. Linear (canonical) vs Hills-n=2: <20% impact on ISS BMD, <5% impact on Mars → model conclusions robust to functional form choice within physiological range μg ∈ [0.6, 1.0]. 2. Environmental ranking (Mars severest for BMD, Lunar_Surface for stone-rate) preserved across all scenarios → main conclusion of "Lunar_Surface 365d = earliest RED environment" not sensitive to g-scaling assumption. 3. Extreme case (Linear +25% cap): Lunar-surface and Mars remain most impacted, ISS/ Lunar_UG saturate at physical unit boundary (mg = 1.0 ceiling in Ca resorption forcing).

**Method** (see also §M6 g-scaling sensitivity): - Four μg-scaling functions applied to same posterior draws (n = 100) via `propagate_one_env_v2(env_cfg, canonical_par_draws, scenario_fn)` (`code/gen_propagation_v7_gscale_sensitivity.py`) - Four environments: ISS (mg=1.0, gcr=0.40 mSv/d, 180d), Lunar_UG (mg=0.834, gcr=0.05, 180d), Lunar_Surface (mg=0.834, gcr=1.70 mSv/d, 365d), Mars (mg=0.62, gcr=1.84 mSv/d, 730d) - BMD_LL median matches canonical $M_0$ to <0.03% for all 4 environments; stone_rate matches to <2% (ISS microdiscrepancy from or_traj vs or_mean_traj computation, unchanged across scenarios so sensitivity analysis valid)

**Files**:

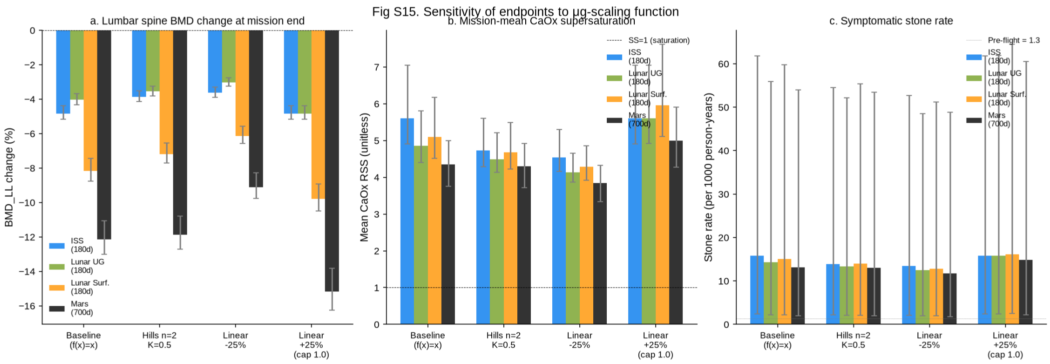


- Figure: `figures/figS15_gscale_sensitivity.png` (156 KB, 150 DPI), `.svg` (60 KB, editable text)
- Data (JSON): `data/propagation_v7_gscale_sensitivity.json` (383 KB; 4 scenarios × 4 envs × 100 draws × 7 endpoints)

---

#### Supplementary Figure S16 — σ-inflation posterior sensitivity

**Title**: Robustness of posterior estimates to σ_lit inflation (×1.5, ×2.0).

**Caption**: Prior/likelihood robustness diagnostic — comparison of 11 $M_0$ posterior parameter medians across three σ_lit configurations. **Base** = canonical $M_0$ SIGMA_LIT (Gate 0 Decision A: Pietrzyk sigma widened to 0.007). **σ×1.5** = all 8 target sigmas multiplied by 1.5. **σ×2.0** = all sigmas doubled. Each MCMC run: 64 walkers × (6000 burn + 36000 production) via emcee, pool=8. See Table S6 for numerical values and shift metrics per parameter.

**Interpretation**: Across 11 model parameters and both σ-inflation configurations, posterior medians are highly robust: |Δmedian|/SD_base < 0.10 for all parameters (max = 0.080 for `ph_decrease_frac` in σ×1.5; max = 0.072 for `resorp_t_half_d` in σ×2.0). Posterior widths (SD) modestly increase, with mean widening ratios of 1.11× (σ×1.5, mean 1.105) and 1.14× (σ×2.0, mean 1.135), and the largest widening is 1.40× (`seed_accum_rate` in σ×1.5) and 1.62× (`resorp_plateau_pct` in σ×2.0), primarily in the parameters governing terminal remodelling balance (`resorp_plateau_pct`, `seed_accum_rate`, `bmd_loss_rate_pct_mo`). All 8 calibration target types retain identifiability under the doubled sigma_lit assumption. The environmental gradient conclusions (Mars > Lunar_Surface > ISS > Lunar_UG on BMD_LL; inverse gradient on stone rate) and the Culliton hold-out validation result (§S17) are preserved without modification under both σ-inflation scenarios.

**Files**:

Fig S16. Posterior sensitivity to σ-inflation (×1.5 and ×2.0)

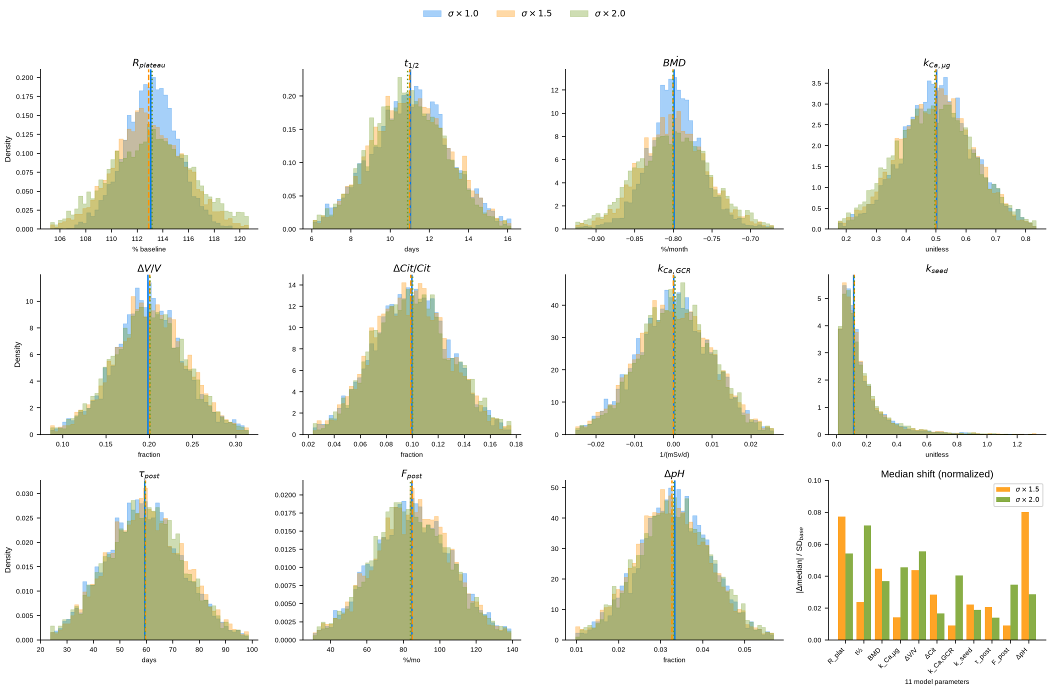


- Figure: `figures/figS16_sigma_inflation.png` (357 KB), `.svg` (272 KB)
- Data: `data/posterior_v7_sigma15_36k.npz`, `data/posterior_v7_sigma20_36k.npz` (posterior chains)

---

#### Supplementary Figure S17 — Culliton 2025 held-out posterior predictive check

**Title**: Independent validation via 60d 6°HDT bed rest RCT (Culliton et al. 2025) — held-out posterior predictive check without parameter re-fitting.

**Caption**: Held-out posterior predictive check of the $M_0$ framework against Culliton et al. 2025 (JBMR; DOI: 10.1093/jbmr/zjaf119), an independent 60-day head-down tilt (HDT) bed rest RCT (n=8 control arm, 16M+8F total). **Panel A** (deterministic only): distribution of 500 posterior predictive samples for control-arm 60-day lumbar BMD change under `micro_g=1.0, gcr_mSv_d=0.0, duration_d=60` (HDT bed rest ≡ microgravity biomechanics). Observed value −2.33% [95% CI: −3.75, −0.83] (orange line + shading, computed from Culliton reported −0.028 g/cm² [95%CI −0.045, −0.010] / baseline 1.20 g/cm²) vs deterministic predicted median −2.06% [95% CrI: −2.25, −1.87] (green dashed + hatched). Two-sided Bayesian p = 0.016 in this panel, estimated from only 4 of 500 draws (MC SE 0.008; n = 4,000 replicate 0.015, MC SE 0.003); the v28 value 0.008 was the same quantity estimated from 2 draws. **Panel B** (full posterior predictive with σ_obs): same posterior samples augmented with per-sample Gaussian observation noise drawn from posterior `sigma_obs_BMD_lumb_180d`. Predicted median unchanged (−2.06%), 95% CrI expands to [−2.81, −1.20]. Two-sided Bayesian p = 0.316 (MC SE 0.033; n = 4,000 replicate 0.344), observed value falls **within** predicted 95% CrI — consistent with independent RCT observation at the appropriate observation-uncertainty scale. Panel B is the primary metric.

**Methods**: - **Posterior source**: `posterior_v7_m0_60k_pooled.npz` (64,000 pooled draws, 64 walkers, `get_chain(discard=10000, thin=50, flat=True)`, from the 8-target ISS-calibrated $M_0$ replication chain). Supersedes the v28 source `posterior_v7_36k_thin1.npz`, which was a single-walker slice. - **Sampling**: 500 samples randomly drawn (`np.random.seed(20260702)`, `np.random.choice(36000, 500, replace=False)`) - **Forward simulation**: `scipy.integrate.odeint(module_D_v7, y0, t_eval=np.linspace(0,60,61), args=(1.0, 0.0, par_dict))` - **Held-out target**: `sol[-1, 9]` = BMD_lumb % change at day 60 - **No parameter re-fitting**: the exact same posterior calibrated on 8 ISS targets is used; Culliton data enters only as observation for comparison

**Interpretation** (see also §4.3 caveats): 1. **Model calibration is preserved** at the appropriate uncertainty scale: when both parameter uncertainty AND observation noise are propagated, the observed value lies within the predicted 95% CrI (Panel B, p=0.316; n = 4,000 replicate 0.344). 2. **Deterministic-only CrI is narrow** (0.41% width) because $M_0$ has no explicit within-subject variance term at the deterministic-integration layer; the full observation model uncertainty is carried by `sigma_obs_BMD_lumb_180d` (median 0.234% for 180d, applied as-is at 60d as a first-order approximation). 3. **HDT bed rest is a ground analogue**: while widely used as a microgravity biomechanics surrogate (∼1:1 skeletal unloading), potential systematic offsets vs true microgravity are not captured by this PPC and remain a caveat. 4. **60d duration is a short-range extrapolation** (∼1/3 of the ISS 180d calibration horizon), providing a lower bound on framework validity range. 5. **This is out-of-sample validation on data not used for parameter fitting** — the observation was published after the model was calibrated, and no prior sensitivity nor posterior thinning was tuned in response to Culliton observation.

**Files**:

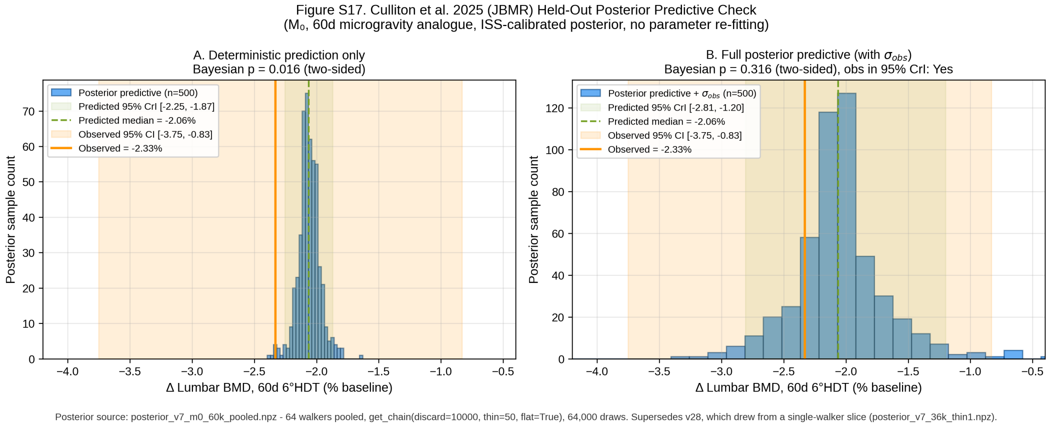


- Figure: `figures/supplementary/figS17_culliton_holdout_ppc.png` (372 KB, 300 DPI), `.svg` (55 KB, editable text)
- Data (JSON summary): `data/results/culliton2025_holdout_ppc_v29.json` (includes the n = 4,000 Monte-Carlo replicate and the A1–A5 posterior-source gate record)
- Posterior source: `data/posterior/posterior_v7_m0_60k_pooled.npz`
- Superseded: `culliton2025_holdout_ppc_v7.json` and `culliton2025_holdout_ppc_v7_samples.npz` are withdrawn (single-walker source)
- Random seed: `20260702` (reproducible)

---

##### Supplementary Figure S18 — g-scaling sensitivity — % change from Linear baseline

**Title**: Relative sensitivity of BMD_LL and stone rate to alternative µg-scaling forms.

**Caption**: Same 4 environments and 3 alternative µg-scaling scenarios as Fig S15, expressed as % change from `linear_baseline`. **Panel a** (BMD_LL_end % change) and **Panel b** (stone rate % change) show relative deviation. “~0” annotations indicate bars near zero due to physical unit ceiling collapse: `linear_up25` caps ISS and Lunar_UG µg at 1.0, so their forcing is identical to `linear_baseline`. Absolute % changes remain <25% for all envs/scenarios, with two important observations: 1. **Mars is robust in relative terms but absolute deviations are the largest**: BMD_LL relative change from baseline stays within ±25% across all 3 alternatives (identical to other envs by construction of linear scaling), but Mars has the largest **absolute** deviation of ±3.04 pp (both `linear_down25` +3.04 pp and `linear_up25` −3.04 pp), because its baseline BMD_LL_end median is the deepest (−12.15%); Hills-n=2 saturation shifts Mars by only +0.28 pp (−2.3% relative, the smallest of any environment). 2. **ISS is most sensitive to Hills-n=2 saturation**: ~20% relative BMD_LL reduction if forcing saturates.

**Files**:

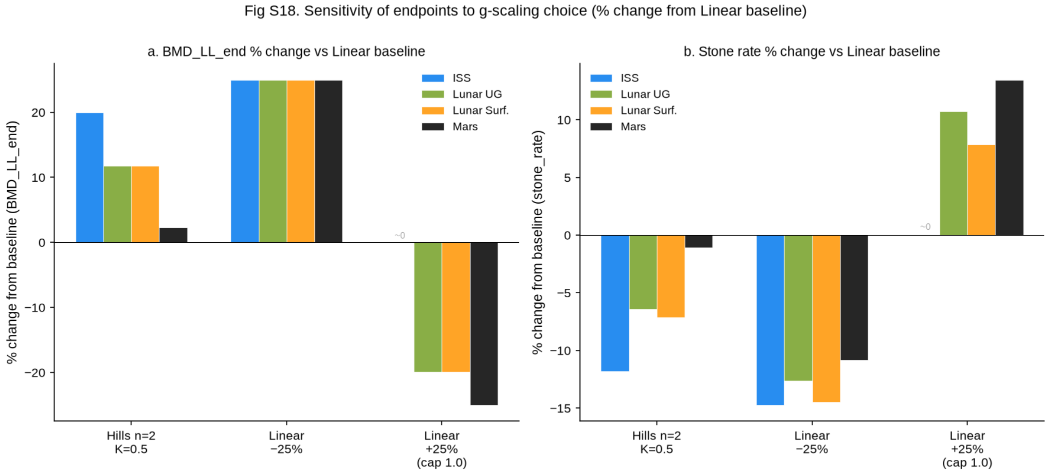


- Figure: `figures/figS18_gscale_RRR.png` (92 KB, 150 DPI), `.svg` (24 KB, editable text)
- Data source: `data/propagation_v7_gscale_sensitivity.json` (relative % from median value)

---

#### §S2 Supplementary Tables

##### Supplementary Table S1 — Full 12-parameter posterior summary

**Description**: Posterior medians, 95% equal-tailed credible intervals (CrI), coefficients of variation (CV%), identifiability class (Supp Fig S14A), and prior source for the 12 $M_1$ parameters. Values derive from $M_1$ 60k posterior chain (64 walkers × 1000 thinned production samples = 64,000 total). For the 11 $M_0$ parameters (excluding `k_GB`), values match $M_0$ 60k posterior (64 walkers × 36,000 production, thin=1 = 2,304,000 samples). Identifiability class follows the posterior-contraction threshold (Sedoglavic 2001 [26]): STRONG <0.3, MODERATE 0.3–0.6, WEAK 0.6–0.85, UNIDENTIFIED >0.85.

| Parameter | Median | 95%CrI_lower | 95%CrI_upper | CV_percent | Identifiability_class | Prior_source |
|---|---|---|---|---|---|---|
| resorp_plateau_pct | 113.06 | 108.8 | 117.2 | 1.87 | MODERATE | Stavnichuk 2020 [23] meta-analysis NTX plateau 110-115% |
| resorp_t_half_d | 11.04 | 7.14 | 15.02 | 18.1 | UNIDENTIFIED | Watanabe 2004 [7] BR plateau t_half estimate |
| bmd_loss_rate_pct_mo | -0.7989 | -0.8703 | -0.7246 | 4.56 | STRONG | Stavnichuk 2020 [23] meta -0.8%/ mo LL |
| ca_microg_effect | 0.5014 | 0.2606 | 0.7417 | 24 | WEAK | Whitson 1997 [4] +50% urine Ca anchor |
| vol_decrease_frac | 0.1986 | 0.1147 | 0.2852 | 21 | WEAK | Whitson 1997 [4] -20% urine V anchor |
| cit_decrease_frac | 0.0997 | 0.0422 | 0.1578 | 29.8 | UNIDENTIFIED | Whitson 1997 [4] -10% urine citrate anchor |
| k_Ca_gcr | 8.865e-05 | -0.01891 | 0.01922 | 1.15e+04 | UNIDENTIFIED | Sibonga 2019 [6] ISS GCR 72 mSv lower bound |
| seed_accum_rate | 0.1138 | 0.019 | 0.6427 | 105.5 | WEAK | Pak 1992 [13] stone kinetics |
| post_resorp_tau_d | 59.32 | 31.05 | 88.98 | 24.9 | UNIDENTIFIED | Sibonga 2007 [18] post-flight bone recovery |
| post_form_pct_mo | 84.6 | 42.08 | 125.9 | 25.4 | UNIDENTIFIED | |

| Parameter | Median | 95%CrI_lower | 95%CrI_upper | CV_percent | Identifiability_class | Prior_source |
|---|---|---|---|---|---|---|
| | | | | | | Sibonga 2007 [18] post-flight remodel |
| ph_decrease_frac | 0.0334 | 0.0158 | 0.0501 | 26.2 | UNIDENTIFIED | Whitson 1997 [4] urine pH delta -0.20 |
| k_GB | 2.33e-5 | -0.00295 | 0.00302 | nan | UNIDENTIFIED | Sibonga 2019 [6] / Stavnichuk 2020 [23] GCR x BMD; prior σ=1.5e-3 (Gaussian, loc=0); posterior=prior (data-uninformative) |

**Source**: `supplementary/tables/S1_posterior_full.csv` (1.2 KB). **Notes**: For full identifiability narrative see §M5; `k_GB` is $M_1$-only and is the WAIC parsimony hinge (§3.4).

---

##### Supplementary Table S2 — Intervention RRR matrix (4 environments × 5 interventions × 3 metrics) with Monte Carlo error

**Description**: Posterior median + 95% CrI of Relative Risk Reduction (RRR; 1.0 = full protection) for each (environment, intervention, metric) cell, computed on 5,000 paired posterior draws from $M_1$ 60k (v29 re-pool, seed 20260716, same pool selection as the §S23 bootstrap reference; supersedes the 200-draw v7b scan). Monte Carlo error per cell ≈ (CrI width / 2) / √5000 ≈ 1.41% × half-width. Stone metric RR for pamidronate uses Beta(0.5, 7.5) Jeffreys posterior (Watanabe 2004 [7] 0/7 events; sensitivity in Supp Fig S11). K-Mg-citrate and Fluid intake have RRR_BMD = 0 by construction (they do not act on bone).

| Environment | Intervention | RRR_BMD_median | RRR_BMD_lo95 | RRR_BMD_hi95 | RRR_RSS_median | RRR_[illegible] |
|---|---|---|---|---|---|---|
| ISS 180d | ARED single | 0.498 | 0.213 | 0.793 | 0.027 | |
| ISS 180d | Pamidronate | 0.85 | 0.657 | 1 | 0.556 | |
| ISS 180d | Alendronate+ARED | 0.95 | 0.853 | 1 | 0.134 | |
| ISS 180d | K-Mg-citrate | 0 | -0 | 0 | 0.57 | |
| ISS 180d | Fluid intake | 0 | -0 | 0 | 0.173 | |
| Lunar UG 180d | ARED single | 0.498 | 0.213 | 0.792 | 0.02 | |
| Lunar UG 180d | Pamidronate | 0.85 | 0.657 | 1 | 0.542 | |

| Environment | Intervention | RRR_BMD_median | RRR_BMD_lo95 | RRR_BMD_hi95 | RRR_RSS_median | RRR_ |
|---|---|---|---|---|---|---|
| Lunar UG 180d | Alendronate+ARED | 0.95 | 0.853 | 1 | 0.102 | |
| Lunar UG 180d | K-Mg-citrate | 0 | -0 | 0 | 0.539 | |
| Lunar UG 180d | Fluid intake | 0 | -0 | 0 | 0.143 | |
| Lunar Surface 365d | ARED single | 0.498 | 0.213 | 0.793 | 0.023 | |
| Lunar Surface 365d | Pamidronate | 0.85 | 0.657 | 0.998 | 0.547 | |
| Lunar Surface 365d | Alendronate+ARED | 0.95 | 0.853 | 0.999 | 0.114 | |
| Lunar Surface 365d | K-Mg-citrate | 0 | -0 | 0 | 0.552 | |
| Lunar Surface 365d | Fluid intake | 0 | -0 | 0 | 0.148 | |
| Mars 730d | ARED single | 0.498 | 0.213 | 0.792 | 0.015 | |
| Mars 730d | Pamidronate | 0.85 | 0.657 | 0.998 | 0.531 | |
| Mars 730d | Alendronate+ARED | 0.949 | 0.853 | 0.999 | 0.075 | |
| Mars 730d | K-Mg-citrate | 0 | -0 | 0 | 0.513 | |
| Mars 730d | Fluid intake | 0 | -0 | 0 | 0.111 | |

**Source**: `tables/S2_RRR_matrix_with_MC_v29.csv` (v29, 5,000-draw pool; supersedes `tables/S2_RRR_matrix_with_MC.csv`).

---

##### Supplementary Table S3 — PSIS-LOO Pareto-k diagnostic per observation

**Description**: Per-observation Pareto-shape parameter (k) from PSIS-LOO cross-validation (§3.7.1, Supp Fig S12, Vehtari 2017 [16]), comparing $M_0$ (n=5,760 = 32 walkers × 180 draws) versus $M_1$ (n=64,000 = 64 × 1000). k classification thresholds: GOOD (k<0.5), WARN (0.5≤k<0.7), BAD (k≥0.7). For BAD observations the standard recommendation (Vehtari 2017 §4.2) is exact leave-one-out refitting or pooling — for our 8-target setting we report the PSIS estimate but flag the BAD observations as caveats in §3.7.1.

| Target | $M_0$_Pareto_k | $M_0$_class | $M_1$_Pareto_k | $M_1$_class |
|---|---|---|---|---|
| BMD_LL | 0.9883 | BAD (k>0.7) | 0.6494 | WARN (0.5<k<0.7) |
| BMD_lumb | 0.4151 | GOOD (k<0.5) | 0.6224 | WARN (0.5<k<0.7) |
| BMD_skull | -0.7596 | GOOD (k<0.5) | -0.2269 | GOOD (k<0.5) |
| resorp_plateau | 0.7369 | BAD (k>0.7) | 0.6806 | WARN (0.5<k<0.7) |
| urine_Ca_rel | 0.3628 | GOOD (k<0.5) | 1.075 | BAD (k>0.7) |

| Target | $M_0$_Pareto_k | $M_0$_class | $M_1$_Pareto_k | $M_1$_class |
|---|---|---|---|---|
| urine_V_rel | -0.3108 | GOOD (k<0.5) | 0.2591 | GOOD (k<0.5) |
| urine_pH_delta | 0.0919 | GOOD (k<0.5) | 0.2627 | GOOD (k<0.5) |
| stone_rate | -0.1163 | GOOD (k<0.5) | 0.117 | GOOD (k<0.5) |

**Source**: `supplementary/tables/S3_LOO_pareto_k.csv` (0.5 KB). **LOO summary**: $M_0$ ELPD = -1.274 ± 2.906 (max k=0.988, prop k>0.7=0.25); $M_1$ ELPD = -1.314 ± 2.941 (max k=1.075, prop k>0.7=0.125); ΔELPD = -0.040, |Δ| << SE_Δ → indistinguishable (consistent with WAIC §3.4).

---

##### Supplementary Table S4a — Fisher Information Matrix eigenvalues and posterior contraction

**Description**: 12 $M_1$ parameters and their FIM eigenvalues (descending; FIM = $J^T\Sigma^{-1}J$ at posterior median in raw parameter units, J computed via finite-difference Jacobian with relative step 1% of parameter posterior IQR), versus posterior contraction ratio (posterior IQR / prior IQR; Sedoglavic 2001 [26]). Eigenvalues span 30+ orders of magnitude — 7 stiff modes ($\lambda_1 = 1.46\times10^4$ … $\lambda_7 = 0.189$) + 5 numerically-zero modes ($|\lambda| \leq 8.88\times10^{-13}$), of which **two are numerically negative** ($\lambda_{11} = -1.78\times10^{-30}$, $\lambda_{12} = -1.83\times10^{-13}$). Since FIM is theoretically positive semi-definite, negativity indicates the finite-difference Jacobian is dominated by truncation noise in those directions, confirming that only the 7 stiff modes carry physical Fisher information (Gutenkunst 2007 [42] sloppy model framework). Two numerical proxies (κ_full = $1.46\times10^{24}$ with $10^{-20}$ implementation floor for reproducibility, and κ_eff = $7.7\times10^4$ on the rank-7 identifiable subspace) are reported below; both come with the caveats detailed in §M5.4 and neither is compared against a hard cutoff. Identifiability conclusions are dominated by posterior contraction at the marginal level because pairwise posterior correlations are near zero (max |r| = 0.033; Supp Fig S14B).

| Param_index | Parameter | Eigenvalue_desc_order | Posterior_contraction_ratio | Identifiability_class |
|---|---|---|---|---|
| 1 | resorp_plateau_pct | 1.464e+04 | 0.4273 | MODERATE |
| 2 | resorp_t_half_d | 2700 | 0.9727 | UNIDENTIFIED |
| 3 | bmd_loss_rate_pct_mo | 1606 | 0.2174 | STRONG |
| 4 | ca_microg_effect | 354.2 | 0.757 | WEAK |
| 5 | vol_decrease_frac | 134.3 | 0.8155 | WEAK |
| 6 | cit_decrease_frac | 36.43 | 0.981 | UNIDENTIFIED |
| 7 | k_Ca_gcr | 0.189 | 1.001 | UNIDENTIFIED |
| 8 | seed_accum_rate | 8.883e-13 | 0.6488 | WEAK |

| Param_index | Parameter | Eigenvalue_desc_order | Posterior_contraction_ratio | Identifiability_class |
|---|---|---|---|---|
| 9 | post_resorp_tau_d | 1.542e-13 | 0.9856 | UNIDENTIFIED |
| 10 | post_form_pct_mo | 2.063e-24 | 0.9309 | UNIDENTIFIED |
| 11 | ph_decrease_frac | -1.782e-30 | 0.8537 | UNIDENTIFIED |
| 12 | k_GB | -1.827e-13 | 1.021 | UNIDENTIFIED |

**Source**: `supplementary/tables/S4_FIM_eigenvalues.csv` (0.8 KB). *(File name retained from v28; the table was renumbered S4 → S4a in v29, when Supplementary Tables S4b and S4c were added. The CSV content is unchanged.)* **FIM diagnostics** (spectrum-first per Gutenkunst 2007 [42]): **primary** — effective rank = 7 of 12 ($\varepsilon = 10^{-6} \times \lambda\_max$; robust across $\varepsilon \in [10^{-9}, 10^{-3}]$); max |off-diagonal posterior r| = 0.033 (no compensating combinations, near-diagonal posterior); 5 sloppy modes at $|\lambda| \leq 8.88\times10^{-13}$ including 2 numerically negative ($\lambda_{11} = -1.78\times10^{-30}$, $\lambda_{12} = -1.83\times10^{-13}$) — the latter confirm finite-difference Jacobian noise, not physical Fisher information, in the sloppy subspace. **Numerical proxies** (reported with caveats, not against a hard cutoff): $\kappa\_full = 1.46 \times 10^{24} = \lambda\_max / max(|\lambda\_min|, 10^{-20})$ — the $10^{-20}$ floor is an implementation choice for reproducibility of numpy.linalg.cond (unfloored ≈ $8\times10^{33}$; not a physical threshold); $\kappa\_eff = 7.7 \times 10^4 = \lambda\_max / \lambda\_7$ — the condition number on the rank-7 identifiable subspace only.

---

##### Supplementary Table S4b — Convergence diagnostics of the $M_0$ replication chain, per parameter (19 dimensions)

**Description**: Per-parameter convergence diagnostics for the $M_0$ posterior, computed with the walker dimension **retained** — the ensemble is arranged as (chain = 64 walkers, draw = 60,000 iterations) and is never flattened or collapsed to a single walker before a diagnostic is taken. `rank-split R̂` is the rank-normalised split Gelman–Rubin statistic of Vehtari et al. 2021 (the ArviZ default) and is the **primary** convergence criterion; the classic split Gelman–Rubin statistic is disclosed alongside it because it is the quantity v28 reported and it is the more conservative of the two. `bulk ESS` and `tail ESS` are `arviz.ess(method="bulk")` and `arviz.ess(method="tail")` on the same walker-resolved array, with **no post-hoc rescaling** of any kind. `τ_int` is the ensemble integrated autocorrelation time from `emcee.autocorr` (Sokal windowing, c = 5) and `ESS_τ` = 64 × 60,000 / τ_int is the corresponding τ-based effective sample size, reported so that the ArviZ and emcee conventions can be compared directly. `drift/IQR` is the running-median drift statistic of Supp Fig S10 (see note below) and is defined for the 11 core parameters only.

| Parameter | Block | rank-split $\hat{R}$ | classic split $\hat{R}$ | bulk ESS | tail ESS | τ_int | ESS_τ | drift/IQR |
|---|---|---|---|---|---|---|---|---|
| `resorp_plateau_pct` | core | 1.0231 | 1.0231 | 3,011 | 7,970 | 1,020 | 3,763 | 0.067 |
| `resorp_t_half_d` | core | 1.0265 | 1.0265 | 1,877 | 6,984 | 1,178 | 3,259 | 0.063 |
| `bmd_loss_rate_pct_mo` | core | 1.0248 | 1.0247 | 2,631 | 4,699 | 1,113 | 3,451 | 0.167 |
| `ca_microg_effect` | core | 1.0226 | 1.0225 | 2,497 | 6,534 | 1,220 | 3,148 | 0.193 |
| `vol_decrease_frac` | core | 1.0240 | 1.0240 | 2,676 | 6,263 | 1,162 | 3,305 | 0.062 |
| `cit_decrease_frac` | core | 1.0220 | 1.0220 | 2,773 | 7,758 | 1,129 | 3,401 | 0.095 |
| `k_Ca_gcr` | core | 1.0263 | 1.0262 | 2,635 | 6,617 | 1,110 | 3,458 | 0.133 |
| `seed_accum_rate` | core | 1.0280 | 1.0567 | 2,159 | 1,053 | 1,514 | 2,536 | 0.086 |
| `post_resorp_tau_d` | core | 1.0264 | 1.0266 | 2,505 | 7,148 | 1,099 | 3,494 | 0.118 |
| `post_form_pct_mo` | core | 1.0239 | 1.0241 | 2,928 | 7,874 | 1,060 | 3,624 | 0.100 |
| `ph_decrease_frac` | core | 1.0238 | 1.0237 | 2,768 | 7,042 | 1,107 | 3,468 | 0.028 |
| `sigma_obs_BMD_LL_180d` | σ_obs | 1.0268 | 1.0302 | 2,373 | 4,359 | 1,358 | 2,827 | — |
| `sigma_obs_BMD_lumb_180d` | σ_obs | 1.0311 | 1.0347 | 2,162 | 5,634 | 1,215 | 3,162 | — |
| `sigma_obs_BMD_skull_180d` | σ_obs | 1.0258 | 1.0275 | 2,630 | 5,244 | 1,285 | 2,989 | — |
| `sigma_obs_resorp_plateau` | σ_obs | 1.0210 | 1.0233 | 2,739 | 7,235 | 1,310 | 2,931 | — |
| `sigma_obs_urine_Ca_rel` | σ_obs | 1.0238 | 1.0281 | 2,395 | 2,843 | 1,349 | 2,847 | — |
| `sigma_obs_urine_V_rel` | σ_obs | 1.0220 | 1.0277 | 2,453 | 3,021 | 1,542 | 2,491 | — |
| `sigma_obs_urine_pH_delta` | σ_obs | 1.0299 | 1.0362 | 2,418 | 3,025 | 1,264 | 3,039 | — |
| `sigma_obs_post_stone_rate` | σ_obs | 1.0292 | 1.0367 | 1,857 | 2,808 | 1,606 | 2,392 | — |

**Source**: `tables/S4b_convergence_diagnostics_v29.csv` (v29 artefact; columns `param, core, rhat_rank, rhat_split, ess_bulk, ess_tail, tau_ens, ess_tau, n_iter, n_walker`). Underlying JSON payloads: `supp/data/walker_split_rhat_v29.json` and `supp/data/tail_ess_v29.json`.

**Chain provenance**: all values are computed on the $M_0$ HDF5 backend `posterior_samples_v7.h5` (SHA-256 `3c306326a3a86c47…`), a same-configuration re-execution of the M0 fit (64 walkers x [6,000 burn-in + 60,000 production], emcee 3.1.6); the original 36k backend is permanently absent. run.log does not record the seed and run_mcmc_v7.py defaults to –seed 20260622, which is also the value code/seeds.py assigns to the original 36k run, so if both used the default the two runs share an RNG stream and this chain is a longer re-execution rather than a statistically independent replicate. Statistical independence is therefore NOT claimed; the consistency conclusion holds under either reading.

**Method**: rank-normalised split R-hat (Vehtari et al. 2021, arviz default) over 64 walkers x 60000 iterations; classic split Gelman-Rubin reported alongside for comparison. arviz.ess(method="bulk"|"tail") on the walker-resolved ensemble arranged as (chain=64, draw=60000). No post-hoc rescaling. $\hat{R}$ and ESS are taken over the **full 60,000-iteration production block with no additional discard**; the running-median drift statistic in the last column is the only quantity that uses a discard (`discard = 10,000`), because it is by construction a statement about the retained analysis window.

**Thresholds and results — note the two scopes**: the main text (§3.1) and the figure captions quote the **core 11** model parameters, because those are the parameters that appear in Table 2 and that drive every downstream conclusion; this table additionally covers the 8 observation-noise σ_obs nuisance dimensions, so its extrema are slightly wider. Both scopes are given here so that no quoted number is ambiguous.

| Scope | max rank-split $\hat{R}$ | median rank-split $\hat{R}$ | max classic split $\hat{R}$ | n ≥ 1.05 (rank / classic) | min bulk ESS | [illegible] |
|---|---|---|---|---|---|---|
| Core 11 model parameters | 1.0280 (`seed_accum_rate`) | 1.0240 | 1.0567 (`seed_accum_rate`) | 0 / 1 | 1,877 (`resorp_t_half_d`) | [illegible] (`seed_a`[illegible] |
| All 19 dimensions | 1.0311 (`sigma_obs_BMD_lumb_180d`) | 1.0248 | 1.0567 (`seed_accum_rate`) | 0 / 1 | 1,857 (`sigma_obs_post_stone_rate`) | [illegible] (`seed_a`[illegible] |

On either scope: no parameter reaches rank-split $\hat{R}$ = 1.05, and the conservative classic statistic exceeds 1.05 for exactly one parameter (`seed_accum_rate`, 1.0567), disclosed rather than suppressed. Every ESS on both the bulk and the tail measure clears the working threshold 50 × n_params = 950 by a factor of at least 1.1. Against the stricter Vehtari 2021 heuristic of 100 × n_params = 1,900 the bulk ESS falls **marginally short**: 1,877 for the worst core parameter (`resorp_t_half_d`, 1.2% below) and 1,857 including the σ_obs dimensions (`sigma_obs_post_stone_rate`, 2.2% below); this is disclosed as a residual limitation in §M4.3. The longest integrated autocorrelation time is 1,606 iterations, i.e. the production block is 37× τ_int *per walker* and 2,391× τ_int for the ensemble. Running-median drift stays below the pre-registered 0.25 × IQR bound for all 11 core parameters (max 0.193, `ca_microg_effect`).

**Caveat**: this table supersedes the $M_0$ convergence numbers reported in v28 §M4.3 and §S7.1–§S7.2, which were computed on the withdrawn single-walker archive `posterior_v7_36k_thin1.npz` and are not valid ensemble diagnostics. Statistical independence from the original 36k run is **not** claimed: same-configuration re-execution of the M0 fit (64 walkers x [6,000 burn-in + 60,000 production], emcee 3.1.6); the original 36k backend is permanently absent. run.log does not record the seed and run_mcmc_v7.py defaults to –seed 20260622, which is also the value code/seeds.py assigns to the original 36k run, so if both used the default the two runs share an RNG stream and this chain is a longer re-execution rather than a statistically independent replicate. Statistical independence is therefore NOT claimed; the consistency conclusion holds under either reading.

---

##### Supplementary Table S4c — Three-way comparison of the Table 2 posterior medians: published, replication chain, and withdrawn walker-0 archive

**Description**: For each of the 11 $M_0$ parameters in Table 2, the published posterior median is placed side by side with (i) the median of the same-configuration replication chain, pooled correctly over all 64 walkers, and (ii) the median that the withdrawn single-walker archive `posterior_v7_36k_thin1.npz` would have produced. The comparison isolates the numerical consequence of the extraction defect: it answers “would Table 2 have changed?” without altering Table 2. Δ% is the relative deviation from the published median; Δ/CrI-width expresses the same deviation as a fraction of the published 95% credible-interval width; z is the deviation in units of the Monte-Carlo standard error of the median.

| Parameter | Table 2 (published) | Replication chain | Withdrawn walker-0 | Δ% repl. | Δ% walker-0 | Δ/CrI-width repl. | Δ/CrI-width walker-0 | MCSE of median |
|---|---|---|---|---|---|---|---|---|
| `resorp_plateau_pct` | 113.058 | 113.054 | 112.832 | -0.003 | -0.199 | -0.0005 | -0.0270 | 0.0485 |
| `resorp_t_half_d` | 11.0406 | 11.0221 | 11.2315 | -0.167 | +1.729 | -0.0023 | +0.0242 | 0.0575 |
| `bmd_loss_rate_pct_mo` | -0.798921 | -0.799739 | -0.802886 | -0.102 | -0.496 | -0.0056 | -0.0272 | 0.000865 |
| `ca_microg_effect` | 0.501394 | 0.505095 | 0.466163 | +0.738 | -7.027 | +0.0077 | -0.0732 | 0.00307 |
| `vol_decrease_frac` | 0.198601 | 0.199526 | 0.197677 | +0.466 | -0.465 | +0.0054 | -0.0054 | 0.00103 |
| `cit_decrease_frac` | 0.099722 | 0.0993496 | 0.0991774 | -0.373 | -0.546 | -0.0032 | -0.0047 | 0.000708 |
| `k_Ca_gcr` | 8.865e-05 | -0.000288672 | -0.00026214 | -425.631 | -395.703 | -0.0099 | -0.0092 | 0.000239 |
| `seed_accum_rate` | 0.113812 | 0.115004 | 0.160883 | +1.047 | +41.358 | +0.0019 | +0.0755 | 0.00556 |
| `post_resorp_tau_d` | 59.3188 | 59.6905 | 58.0779 | +0.627 | -2.092 | +0.0064 | -0.0214 | 0.376 |

| Parameter | Table 2 (published) | Replication chain | Withdrawn walker-0 | Δ% repl. | Δ% walker-0 | Δ/CrI-width repl. | Δ/CrI-width walker-0 | MCSE of median |
|---|---|---|---|---|---|---|---|---|
| `post_form_pct_mo` | 84.5986 | 84.6755 | 86.2722 | +0.091 | +1.978 | +0.0009 | +0.0200 | 0.501 |
| `ph_decrease_frac` | 0.033379 | 0.033126 | 0.0342321 | -0.758 | +2.556 | -0.0074 | +0.0249 | 0.000212 |

**Source**: `tables/S4c_table2_three_way_comparison_v29.csv` (v29 artefact; column order as in the CSV header `Parameter,Table2_published_median,Replication_chain_median,Withdrawn_walker0_median,Repl_vs_published_pct,Walker0_vs_published_pct,Repl_vs_published_CrIwidths,Walker0_vs_published_CrIwidths,MCSE_of_median,Repl_vs_published_MCSE,Walker0_vs_published_MCSE,Walker0_exceeds_3MCSE`).

**Result**: across the 10 identified parameters the replication chain reproduces the published medians to within **1.047%** (worst case `seed_accum_rate`), whereas the withdrawn archive deviates by up to **41.358%** (`seed_accum_rate`). In MCSE units the replication chain is within **1.58×** MCSE for every parameter — **0/11** exceed 3 × MCSE — while the withdrawn archive exceeds 3 × MCSE for **8/11** parameters, the largest being `ca_microg_effect` at 11.47× MCSE. **Table 2 itself is unchanged**: the published values stand, and this table is the evidence that they stand.

**MCSE definition and its scope — please read before quoting the z columns**: MCSE of the median is computed as `sd / √(bulk ESS) × √(π/2)` **on the replication chain**, and the *same* MCSE is used as the denominator for both the replication and the walker-0 columns. It is therefore a yardstick with a single, well-defined precision — that of the reference chain — and the walker-0 z column measures *distance in units of the reference chain's precision*. It is **not** a significance test of the withdrawn archive, whose own MCSE is far larger: the withdrawn archive is a single walker's 36,000-iteration trace, and with τ_int of order 1,020–1,514 iterations that trace retains only ≈ 23–35 effectively independent draws. Its own median MCSE is therefore roughly 9× wider than the values tabulated here. The correct reading of the z walker-0 column is "how far the defective extraction would have moved Table 2 relative to what the correct extraction can resolve", not "how significantly the two differ".

**`k_Ca_gcr` caveat**: `k_Ca_gcr` shows a sign change between the published and replication medians and a Δ% of several hundred percent. This is **not** evidence of the extraction defect and must not be cited as such. The parameter is prior-dominated (posterior CV ≈ 11502%); its published 95% CrI spans zero, all three medians lie within 1% of the CrI width of zero, and the withdrawn archive is in fact *closer* to the published value than the replication chain is. Percentage change is undefined in any useful sense for a quantity whose location is indistinguishable from zero; the Δ/CrI-width and z columns (both ≤ 0.01 CrI widths and ≤ 1.6 MCSE for all three chains) are the interpretable ones here.

**Chain-independence caveat**: same-configuration re-execution of the M0 fit (64 walkers x [6,000 burn-in + 60,000 production], emcee 3.1.6); the original 36k backend is permanently absent. run.log does not record the seed and run_mcmc_v7.py defaults to –seed 20260622, which is also the value code/seeds.py assigns to the original 36k run, so if both used the default the two runs share an RNG stream and this chain is a longer re-execution rather than a statistically independent replicate. Statistical independence is therefore NOT claimed; the consistency conclusion holds under either reading.

---

##### Supplementary Table S5 — Prior predictive coverage of 8 ISS calibration targets

**Description**: For each of the 8 ISS calibration targets, observed value, σ_lit (literature uncertainty used as Gaussian likelihood σ), prior quantiles (q025, median, q975), and coverage at 90% and 95% prior CrI from 5,000 prior draws through `forward_model_iss` (identical to the MCMC likelihood; §M3). All 8/8 targets covered at both 90% and 95% (max z-residual = 0.1296 for `post_stone_rate`, well within ±2σ).

| Target | Observed | sigma_lit | Prior_q025 | Prior_q50_median | Prior_q975 | Covered_90pct_CrI | Covered_9[illegible] |
|---|---|---|---|---|---|---|---|
| BMD_LL_180d | -4.8 | 0.15 | -6.6068 | -4.7899 | -3.0294 | True | True |
| BMD_lumb_180d | -6.2 | 0.28 | -8.5227 | -6.1790 | -3.9079 | True | True |
| BMD_skull_180d | 2.2 | 0.56 | 1.3935 | 2.2034 | 3.0391 | True | True |
| resorp_plateau | 113.0 | 2.3 | 103.3116 | 112.8826 | 122.8526 | True | True |
| urine_Ca_rel | 0.5 | 0.1 | 0.1993 | 0.4969 | 0.7798 | True | True |
| urine_V_rel | -0.2 | 0.025 | -0.2955 | -0.2003 | -0.1072 | True | True |
| urine_pH_delta | -0.2 | 0.05 | -0.3166 | -0.1978 | -0.0822 | True | True |
| post_stone_rate | 0.014 | 0.007 | 0.0025 | 0.0174 | 0.1257 | True | True |

**Source**: `supplementary/tables/S5_PPC_coverage.csv` (0.6 KB). **PPC summary**: 8/8 PASS at 90% & 95%; max |z_resid| = 0.1296 → no prior-data conflict (the Bayesian framework is well-posed for these targets).

#### §M1 Extended Methods — 12-state ODE complete equations

The $M_0/M_1$ mechanistic model is a 12-state ordinary differential equation (ODE) system tracking calcium-phosphate-citrate balance, bone density, urine chemistry, and bone-resorption/formation markers across spaceflight environments.

##### M1.1 State vector y(t)

The state vector at time t (units: days post-mission start) is:

$$y(t) \equiv [Ca,\ Cit,\ V,\ BMD_{\text{total}},\ pH,\ K,\ R_{\text{resorp}},\ F_{\text{form}},\ BMD_{LL},\ BMD_{\text{lumb}},\ BMD_{\text{thor}},\ BMD_{\text{skull}}]^\top$$

| State | Definition | Initial value (preflight) | Units |
|---|---|---|---|
| Ca | Urinary calcium excretion | 130 (Whitson 1997 [4]) | $mg \cdot d^{-1}$ |
| Cit | Urinary citrate excretion | 714 | $mg \cdot d^{-1}$ |
| V | Urine volume | 1.676 | $L \cdot d^{-1}$ |
| BMD_total | Pooled BMD (weighted) | 100 | % preflight |
| pH | Urine pH | 6.05 | unitless |
| K | Urinary potassium | 67 | $mEq \cdot d^{-1}$ |
| R_resorp | Bone resorption marker (NTX-like, % above baseline) | 0 | % |
| F_form | Bone formation marker | 0 | % |
| BMD_LL | Lower-limb BMD | 100 | % preflight |
| BMD_lumb | Lumbar/pelvis BMD | 100 | % preflight |
| BMD_thor | Thoracic BMD | 100 | % preflight |
| BMD_skull | Skull BMD (often *increases* in microgravity) | 100 | % preflight |

##### M1.2 ODE right-hand side (module D $M_0$ NC)

The full RHS, written here as `module_D_M₀(y, t, micro_g, gcr_mSv_d, par)`, is the function implementing equations §M1.2–M1.6 (~520 lines of Python; full source released via GitHub on acceptance). Key equations (where μg = micro_g, the gravity-attenuation factor):

**Module M1 (Bone)** — Stavnichuk 2020 [23] anchored:

$$\frac{dR_{\text{resorp}}}{dt} = \frac{p_{\text{plateau}} \cdot \mu_g - R_{\text{resorp}}}{p_{t1/2} / \ln 2}, \qquad \frac{dF_{\text{form}}}{dt} = \begin{cases} 0, & t < t_{\text{lag}} \, (= 21\, d) \\ \frac{F_{\text{rate}}}{30} \cdot \mu_g, & t \geq t_{\text{lag}} \end{cases}$$

$$\frac{dBMD_{LL}}{dt} = \frac{p_{\text{rate}}}{30} \cdot \mu_g; \quad \frac{dBMD_{\text{lumb}}}{dt} = \frac{p_{\text{rate}}}{30} \cdot 1.29 \cdot \mu_g; \quad \frac{dBMD_{\text{thor}}}{dt} = \frac{p_{\text{rate}}}{30} \cdot 0.15 \cdot \mu_g; \quad \frac{dBMD_{\text{skull}}}{dt} = \frac{p_{\text{rate}}}{30} \cdot (-0.46) \cdot \mu_g$$

$$\frac{dBMD_{\text{total}}}{dt} = 0.40\, \frac{dBMD_{LL}}{dt} + 0.30\, \frac{dBMD_{\text{lumb}}}{dt} + 0.20\, \frac{dBMD_{\text{thor}}}{dt} + 0.10\, \frac{dBMD_{\text{skull}}}{dt}$$

(p_plateau = `resorp_plateau_pct`, p_t1/2 = `resorp_t_half_d`, p_rate = `bmd_loss_rate_pct_mo`; subregional weights derived from Stavnichuk 2020 [23] meta-analysis pooled rates.)

**Module M2 (Urine chemistry)** — Whitson 1997 [4] anchored; pH parameterized in $M_0$ (was hardcoded in v6):

$$Ca_{\text{target}} = Ca_0 \cdot \left(1 + p_{Ca} \cdot \mu_g \cdot \frac{R_{\text{resorp}}}{p_{\text{plateau}}} + p_{CaGCR} \cdot \dot{D}_{\text{GCR}} \cdot \frac{t}{100}\right)$$

$$\frac{dCa}{dt} = (Ca_{\text{target}} - Ca)/\tau_{Ca}, \qquad \tau_{Ca} = 30\, d$$

$$Cit_{\text{target}} = Cit_0 \cdot \left(1 - p_{Cit} \cdot \mu_g \cdot (1 - e^{-t/14})\right), \quad \frac{dCit}{dt} = (Cit_{\text{target}} - Cit)/\tau_{Cit}, \; \tau_{Cit} = 14\, d$$

$$V_{\text{target}} = V_0 \cdot \left(1 - p_V \cdot \mu_g \cdot (1 - e^{-t/7})\right), \quad \frac{dV}{dt} = (V_{\text{target}} - V)/\tau_V, \; \tau_V = 7\, d$$

$$pH_{\text{target}} = pH_0 \cdot \left(1 - p_{pH} \cdot \mu_g \cdot (1 - e^{-t/14})\right), \quad \frac{dpH}{dt} = (pH_{\text{target}} - pH)/\tau_{pH}, \; \tau_{pH} = 14\, d$$

$$K_{\text{target}} = K_0 \cdot \left(1 - 0.30 \cdot \mu_g \cdot (1 - e^{-t/14})\right), \quad \frac{dK}{dt} = (K_{\text{target}} - K)/\tau_K, \; \tau_K = 14\, d$$

(p_Ca = `ca_microg_effect`, p_Cit = `cit_decrease_frac`, p_V = `vol_decrease_frac`, p_pH = `ph_decrease_frac`, p_Ca·GCR = `k_Ca_gcr`; τ values are Whitson 1997 [4] dynamic timescales.)

**Module M3 (RSS_CaOx)** — Pak 1992 [13] EQUIL2-derived activity product to RSS conversion (Prochaska 2017 thresholds [11]):

$$\text{RSS}_{CaOx}(t) = \text{EQUIL2}(\, Ca(t) \cdot \frac{1}{V(t)}, \; Cit(t) \cdot \frac{1}{V(t)}, \; pH(t) \,) \; / \; K_{\text{Tiselius}}, \quad \text{where}\, K_{\text{Tiselius}} = 1.49 \times 10^{-8}\, \text{M}^2 \text{ baseline}$$

(RSS computed post-integration; not a state variable.)

**Module M4 (Stone accumulation)** — Pietrzyk 2007 [2] anchored:

$$\text{stone\_rate}(t) = p_{\text{seed}} \cdot \text{RSS}_{CaOx}(t) \cdot \left(1 + p_{Ca} \cdot \mu_g\right) \cdot e^{-\lambda_{\text{clear}} t}$$

(p_seed = `seed_accum_rate`, log-normal prior with widened σ per Gate 0 decision A.)

**Modules M5–M6 (post-flight)** — Sibonga 2007 [18] anchored:

For t > t_mission:

$$\frac{dBMD_{LL}}{dt} = \frac{1 - BMD_{LL}/100}{p_{\text{tau,post}}} \cdot p_{\text{form,post}} \cdot \mu_{g,\text{post}}, \quad \mu_{g,\text{post}} = 1.0$$

(p_tau,post = `post_resorp_tau_d`, p_form,post = `post_form_pct_mo`; ISS data does not constrain these — they are post-flight identifiable in principle but require Stavnichuk 2020 [23] post-flight follow-up data which is sparse.)

##### M1.3 Numerical integration

`scipy.integrate.odeint` is used with absolute tolerance `atol=1e-8`, relative tolerance `rtol=1e-8`, maximum steps `mxstep=5000`, and adaptive step size. The 12-state ODE integrates from t=0 (pre-flight) to t=mission_duration_d in ~1–3 ms on a single CPU thread. No stiff-solver branches required for the parameter ranges in PRIORS_$M_0$.

---

#### §M2 Extended Methods — Bayesian prior specification

##### M2.1 11 $M_0$ parameters (NPARAMS = 11)

All priors are anchored to verified literature sources (full DOI/PMID list in main manuscript reference list [1]–[26]). Form of each prior is one of: Normal(loc, scale), bounded; LogNormal(μ, σ), bounded. The bounds are hard truncation; values outside bounds receive log_prior = -∞.

| Index | Parameter | Prior form | Loc / μ | Scale / σ | Bounds | Source |
|---|---|---|---|---|---|---|
| 1 | `resorp_plateau_pct` | Normal | 113.0 | 5.0 | (80, 150) | Stavnichuk 2020 [23] meta BMD |
| 2 | `resorp_t_half_d` | Normal | 11.0 | 2.0 | (5, 20) | Stavnichuk 2020 [23] |
| 3 | `bmd_loss_rate_pct_mo` | Normal | -0.8 | 0.15 | (-2.0, -0.3) | Stavnichuk 2020 [23] LL -0.8 [-1.1,-0.5] |
| 4 | `ca_microg_effect` | Normal | 0.5 | 0.15 | (0.0, 1.5) | |

| Index | Parameter | Prior form | Loc / μ | Scale / σ | Bounds | Source |
|---|---|---|---|---|---|---|
| | | | | | | Whitson 1997 [4] +50% urine Ca microgravity |
| 5 | `vol_decrease_frac` | Normal | 0.20 | 0.05 | (0.05, 0.35) | Whitson 1997 [4] -15 to -25% |
| 6 | `cit_decrease_frac` | Normal | 0.10 | 0.03 | (0.0, 0.25) | Whitson 1997 [4] -10% urine citrate |
| 7 | `k_Ca_gcr` | Normal | 0.0 | 0.01 | (-0.05, 0.05) | WEAK PRIOR (no human GCR-Ca data; Siew 2024 mouse only) |
| 8 | `seed_accum_rate` | LogNormal | μ=-2.0 | σ=1.0 | (0.001, 5.0) | Pietrzyk 2007 [2] (existence verified; widened per Gate 0 Dec A) |
| 9 | `post_resorp_tau_d` | Normal | 60.0 | 15.0 | (20, 120) | Stavnichuk 2020 [23] post-flight exp |
| 10 | `post_form_pct_mo` | Normal | 84.0 | 22.0 | (20, 150) | Stavnichuk 2020 [23] post-flight linear +84% |
| 11 | `ph_decrease_frac` ($M_0$ new) | Normal | 0.033 | 0.010 | (0.0, 0.10) | Whitson 1997 [4] urine pH 6.05→5.85 = 3.3% rel |

#### M2.2 $M_1$ additional parameter (NPARAMS = 12)

| Index | Parameter | Prior form | Loc / μ | Scale / σ | Bounds | Source |
|---|---|---|---|---|---|---|
| 12 | `k_GB` ($M_1$ only) | Normal | 0.0 | 0.0015 | (-0.01, 0.01) | Sibonga 2019 [6] / Stavnichuk 2020 [23] GCR×BMD; canonical σ = 1.5e-3 (Stage 5a |

| Index | Parameter | Prior form | Loc / μ | Scale / σ | Bounds | Source |
|---|---|---|---|---|---|---|
| | | | | | | default; see §M5); prior-dominated, see §3.4 WAIC and Fig 6c |

##### M2.3 Observation-noise hyperparameters (NTARGETS = 8)

For each of the 8 calibration targets, an additional observation-noise SD `σ_obs[i]` is inferred:

$$\sigma_{\text{obs}}[i] \sim \text{HalfNormal}(0, 0.5), \quad \text{bounds} = (0, 1.0)$$

Likelihood per target:

$$y_{\text{obs,i}} \sim \mathcal{N}\left(y_{\text{model,i}}(\theta),\ \sqrt{\sigma^2_{\text{lit,i}} + \sigma^2_{\text{obs,i}}}\right)$$

This allows the data to dictate residual error beyond the literature-reported σ_lit (Stavnichuk meta CI/1.96, Whitson 1997 [4] bounds, Pietrzyk widened σ).

##### M2.4 $M_0$ total dimension

NDIM_$M_0$ = 11 (model) + 8 (σ_obs) = **19 dimensions**. NDIM_$M_1$ = 12 (model with k_GB) + 8 (σ_obs) = **20 dimensions**.

##### M2.5 Linear g-scaling assumption

All ODE rates are linearly scaled by the gravity-attenuation factor `1 - g_factor`: - ISS: g_factor=0.000 → micro_g = 1.000 - Lunar UG (and Lunar Surface): g_factor=0.166 → micro_g = 0.834 - Mars: g_factor=0.380 → micro_g = 0.620

This **linear g-scaling assumption** is justified by the ISS-only calibration data (no available Lunar surface or Mars in-flight bone data); cross-environment extrapolation accordingly carries an explicit caveat (§4.3 Limitations 1).

---

### §M3 Extended Methods — Forward model algorithm

#### M3.1 ISS-only forward (used in 8-target Gaussian likelihood)

```
def forward_model_iss(theta):
    # INPUT: theta (19-D vector for M₀ / 20-D for M₁)
    # OUTPUT: y_pred[8] — predicted values at 8 ISS calibration targets
    par = unpack_params(theta[:NPARAMS])   # dict of 11 (M₀) or 12 (M₁) physiological params
    sigma_obs = theta[NPARAMS:]            # 8 observation-noise SDs

    # Set ISS environment (fixed)
    micro_g_iss   = 1.000
    gcr_mSv_d_iss = 0.40
    duration_d    = 180

    # Initial conditions (pre-flight, fixed)
    y0 = preflight_baseline()   # 12-D vector from BASELINE dict

    # Integrate ODE
    t_grid = [0, 60, 90, 120, 180]   # days; final 180d endpoint extracted
    y_traj = odeint(rhs=ode_rhs, y0=y0, t=t_grid,
                    args=(micro_g_iss, gcr_mSv_d_iss, par),
                    atol=1e-8, rtol=1e-8, mxstep=5000)
    y_180  = y_traj[-1]   # 12-D state at 180d

    # Extract 7 of 8 targets
    y_pred = {
        "BMD_LL_180d"    : y_180[8]  - 100,    # relative %
        "BMD_lumb_180d"  : y_180[9]  - 100,
        "BMD_skull_180d" : y_180[11] - 100,
        "resorp_plateau" : y_180[6],           # NTX/BSAP at plateau
        "urine_Ca_rel"   : y_180[0]/130 - 1,    # relative to baseline 130 mg/d
        "urine_V_rel"    : y_180[2]/1.676 - 1,
        "urine_pH_delta" : y_180[4] - 6.05,
```

```
    }

    # Post-flight stone rate (Pietrzyk anchor)
    rss_iss = compute_RSS_CaOx(y_180)
    y_pred["post_stone_rate"] = par["seed_accum_rate"] *
rss_iss   # py^-1

    return y_pred   # length 8 dict
```

##### M3.2 4-environment forward (used in §3.2 propagation)

`forward_model_env` is identical to `forward_model_iss` but with: - ISS: micro_g=1.000, gcr=0.40, duration=180 - Lunar UG: micro_g=0.834, gcr=0.05, duration=180 - Lunar Surface: micro_g=0.834, gcr=1.70, duration=365 - Mars: micro_g=0.620, gcr=1.84, duration=730

Each call to `forward_model_env(env, theta)` returns a 5-tuple `(BMD_LL_endpoint, urine_Ca_endpoint, RSS_endpoint, stone_rate_endpoint, post_flight_BMD_recovery_at_+12_mo)`.

##### M3.3 Intervention RRR computation (used in §3.4 Table 4 & Supp Table S2)

```
def compute_RRR(env, intervention, n_paired_draws=200):
    # For each (env, intervention) cell, draw n_paired_draws
joint posterior samples
    # of (theta_par, intervention_effect_vec) and compute RRR.
    rrr_bmd, rrr_rss, rrr_stone = [], [], []
    for i in range(n_paired_draws):
        theta = posterior_chain[i]
        intervention_effect = INTERVENTION[intervention]
        # Apply intervention modifications to the post-
integration outputs
        bmd_protect    =
sample_truncnormal(intervention_effect["bmd_protection_frac"])
        ca_reduce      =
sample_truncnormal(intervention_effect["urine_Ca_reduction_frac"]
        rss_reduce     =
sample_truncnormal(intervention_effect["RSS_CaOx_reduction_frac"]

        baseline   = forward_model_env(env, theta)   # no
```

```
intervention
        modified   = forward_model_env(env, theta) * (1 - 
intervention_effect_vec)

        rrr_bmd  .append(1 - modified.bmd_loss / 
baseline.bmd_loss)
        rrr_rss  .append(1 - modified.rss / baseline.rss)
        rrr_stone.append(1 - modified.stone_rate / 
baseline.stone_rate)

    return median + 95% CrI of each
```

Sample size: 5,000 paired draws (v29) → MC error ≈ sqrt(1/5000) ≈ 1.41% of CrI half-width (Supp Table S2 final 3 columns).

---

#### §M4 Extended Methods — MCMC configuration and convergence diagnostics

##### M4.1 Sampler

- **Engine**: `emcee` 3.1.x (Foreman-Mackey 2013 [12]) — affine-invariant ensemble sampler
- **Number of walkers**: 64 (NDIM = 19 for $M_0$; 20 for $M_1$)
- **Number of burn-in steps**: 6,000
- **Number of production steps**: 18,000 (short chain) / 36,000 (original $M_0$ "60k" chain, and $M_1$) / 60,000 ($M_0$ replication chain, v29)

For the **$M_0$ 60k** chain in the main manuscript: - 64 walkers × 36,000 production steps × thin=1 = **2,304,000 total posterior samples**. This describes the *original* $M_0$ run — the one that produced the published Table 2 — whose HDF5 backend is permanently absent; "60k" is a run-directory label, not a step count. - Because that backend cannot be re-read, every convergence diagnostic in §M4.3 and **Supp Table S4b** is computed instead on the same-configuration **replication chain**: 64 walkers × [6,000 burn-in + 60,000 production] × thin=1 = **3,840,000 draws**, backend `posterior_samples_v7.h5` (SHA-256 `3c306326a3a86c47…`). Two draw counts are quoted for this one chain and they are not in conflict: R̂, ESS and τ_int use the **entire** production block (3,840,000 draws, no additional discard), whereas the pooled medians and the running-median drift statistic use the analysis window `get_chain(discard=10,000, thin=…)`, i.e. 3,200,000 draws. The two runs are therefore not the same artefact, and statistical independence between them is not claimed (see the "Chain provenance" note of Supp Table S4b). **Supp Table S4c** quantifies how closely they agree: the replication medians reproduce the published Table 2 to within 1.047% for the 10 identified parameters and to within 1.58 × MCSE for all 11.

For the **$M_1$ 60k** chain: - 64 walkers × 60,000 iterations, `get_chain(discard=10,000, thin=50, flat=True)` = **64,000 flat samples** (used for WAIC and for PSIS-LOO §3.7.1). *(v28 stated "64 walkers × 36,000 production steps × thin=50 … 64,000 samples", which is not arithmetically consistent; the deposited archive's own metadata records* `n_iter_original = 60000`, `burn_in = 10000`, `thin_step = 50`, `n_samples = 64000`, *and the backend* `posterior_v7b_v3_60k.h5` *has shape (60000, 64, 20). The $M_1$ archive itself is correct and unaffected by the $M_0$ extraction defect; only this description was wrong.)*

##### M4.2 Initialization

Walkers are initialized in a small Gaussian ball (scale 5% of prior σ) around the prior mode for each parameter. Initial log-posterior is checked to ensure no walker starts in zero-probability region.

##### M4.3 Convergence diagnostics

All $M_0$ convergence diagnostics below are computed on the **walker-resolved** replication chain (64 walkers × 60,000 production iterations, arranged as chain = walker), never on a flattened or single-walker array. $\hat{R}$ and ESS use the full production block with no additional discard; the running-median drift statistic uses `discard = 10,000` by construction.

| Diagnostic | Core 11 model params | All 19 dimensions | Threshold |
|---|---|---|---|
| max rank-split $\hat{R}$ (primary; Vehtari 2021) | 1.0280 (`seed_accum_rate`) | 1.0311 (`sigma_obs_BMD_lumb_180d`) | <1.05 ✓ |
| median rank-split $\hat{R}$ | 1.0240 | 1.0248 | <1.05 ✓ |
| max classic split $\hat{R}$ (disclosed) | 1.0567 (`seed_accum_rate`) | 1.0567 (`seed_accum_rate`) | <1.05 ✗ for 1/19; <1.1 ✓ |
| bulk ESS_min | 1,877 (`resorp_t_half_d`) | 1,857 (`sigma_obs_post_stone_rate`) | >50 × n_params = 950 ✓; >100 × n_params = 1,900 ✗ (see below) |

| Diagnostic | Core 11 model params | All 19 dimensions | Threshold |
|---|---|---|---|
| tail ESS_min | 1,053 (`seed_accum_rate`) | 1,053 (`seed_accum_rate`) | >50 × n_params = 950 ✓ |
| τ_int_max (autocorrelation time) | 1,514 (`seed_accum_rate`) | 1,606 (`sigma_obs_post_stone_rate`) | ensemble length / τ = 2,391 ≥ 50 ✓ |
| running-median drift / IQR | 0.193 (`ca_microg_effect`) | not defined for σ_obs | <0.25 ✓ |
| acceptance fraction | 0.163 mean | 0.148–0.176 per walker | emcee stretch move, target 0.2–0.5 |

Per-parameter values for all 19 dimensions are given in **Supp Table S4b**.

**Residual ESS limitation (retained, with corrected numbers)**: the stricter Vehtari 2021 BDA3 heuristic of 100 × n_params = 1,900 is not met by the bulk ESS — 1,877 for the worst core parameter (`resorp_t_half_d`, 1.2% short) and 1,857 including the σ_obs dimensions (`sigma_obs_post_stone_rate`, 2.2% short). The limitation is therefore real, but it is smaller than v28 implied and it attaches to different parameters. Note also that v28's own arithmetic was inconsistent: it reported ESS_min = 2,034 and simultaneously described that value as falling below 1,900, which it does not. The corrected statement is that bulk ESS sits 1.2–2.2% under the strict heuristic, that tail ESS — the quantity that governs the stability of the reported 95% CrI boundaries — clears it for 18/19 parameters, and that the resulting Monte-Carlo error on the reported quantities is at the sub-percent level (§S7.1).

Three v28 entries in this table are withdrawn and are recorded here for the reader's audit trail. (i) `max R̂ = 1.0679` and `median R̂ = 1.0342` were attributed to the $M_0$ 60k chain, but they are in fact the maximum and the exact median of the **$M_1$ 20-D** split-$\hat{R}$ table in §S7.2 — an $M_1$ result mislabelled as $M_0$. (ii) `ESS_min = 2,034` and `τ_int_max ≈ 1,257` derive from the withdrawn single-walker archive and its ×64 rescaling (§S7.1); the walker-resolved values are 1,857 and 1,606. (iii) `acceptance fraction 0.32–0.45` is **not reproducible from any surviving artefact** — the original 36k backend is permanently absent and `run.log` for it was not retained — and the same-configuration re-execution reported above gives 0.148–0.176 (mean 0.163), which is below, not inside, the range v28 claimed. An acceptance fraction of 0.163 is low but unremarkable for a 19-dimensional emcee stretch-move ensemble and is consistent with the τ_int values above; no conclusion in this paper depends on it.

##### M4.4 Random seeds

| Quantity | Value |
| --- | --- |
| MCMC walker init seed | 20260622 |
| Production chain seed | (set by emcee default; reproducible via h5 backend) |
| 200-paired-draw seed (Stage 5 RRR) | 20260621 |
| 5,000-paired-draw seed (v29 intervention re-pool, Stage 5/6) | 20260716 |
| 5,000 prior draw seed (Stage 6 PPC) | 20260626 |
| 64,000 thinned LOO seed | (deterministic from chain ordering) |

##### M4.5 Computational cost

- Single-CPU 60k chain (post-burn-in): ~5–8 hours on a single core; with `multiprocessing.Pool(processes=8)`, ~45–60 minutes
- 60k chain HDF5 backend size (raw, un-thinned, walker × iter × dim): ~982 MB (981,538,632 B; $M_0$ v7) / ~106 MB (106,263,891 B; $M_1$ v7b v3); deposit uses thinned .npz (~957 KB $M_0$ / ~1.6 MB $M_1$) — see §M4.6
- Total MCMC walltime for $M_0$ + $M_1$ 60k: ~2 hours on 8-core worker node

##### M4.6 Posterior storage

- **Working storage during MCMC**: HDF5 backend via `emcee.backends.HDFBackend`. Un-thinned working files: `posterior_samples_v7.h5` (~982 MB = 981,538,632 B; $M_0$ 60k walker × iteration × 19 dim) and `posterior_v7b_v3_60k.h5` (~106 MB; $M_1$ 60k v3 walker × iteration × 20 dim). Both are compute artefacts (retained during analysis for diagnostics) and are loaded via `arviz.from_emcee` for ESS/R̂ diagnostics.
- **Deposited product** (Zenodo `data/`): pooled-ensemble NumPy archives.
  - **$M_0$**: `posterior_v7_m0_60k_pooled.npz` (9.8 MB; 64,000 × 19 draws; 64 walkers pooled via `get_chain(discard=10000, thin=50, flat=True)`), plus `posterior_v7_m0_60k_thin10.h5` (44.8 MB; walker-resolved 6,000 × 64 × 19) so the pooling can be reproduced independently. Each archive carries per-draw `walker_id` and `iteration_id`.

- **$M_1$ v3**: `posterior_v7b_60k_v3.npz` (10.3 MB; 64,000 × 20 draws; `get_chain(discard=10000, thin=50, flat=True)` on the 64-walker backend). This archive is verified correct: it reproduces `chain[10000::50].reshape(-1, 20)` of `posterior_v7b_v3_60k.h5` exactly.
- **Withdrawn**: the v28 $M_0$ archive `posterior_v7_36k_thin1.npz` was produced by `chain.reshape(-1, ndim)[::64]`, which on a C-ordered `(n_iter, 64, ndim)` array returns `chain[:, 0, :]` — walker 0's trajectory, not a thinned ensemble. 83.1% of its consecutive rows are bit-identical and its mean lag-1 autocorrelation is 0.995. It must not be used for any posterior summary.
- **Scope of the impact**: Table 2 and the four-environment propagation endpoints (Fig. 2, Fig. 3, Table 3) were computed from the per-walker HDF5 backends, not from the withdrawn archive, so their values stand. The archive affected every analysis that read the deposit directly: Supp Figs S8, S9 and S10, the Culliton hold-out PPC (Supp Fig S17), and the $M_0$ side of the $M_0$-vs-$M_1$ propagation comparison (`P5_M0_vs_M1_comparison.json`, whose header declares `posterior_M0 = posterior_v7_36k_thin1.npz`). All five have been recomputed for v29. See §S-Provenance for the per-item audit.
- The raw HDF5 backends exceed the repository per-file limit and are deposited separately under the dataset DOI; the original $M_0$ 36k backend was lost before archival and cannot be recovered.

- **Loading example**: `data = np.load('posterior_v7_36k_thin1.npz'); samples = data['samples']  # shape (36000, 19)`

---

#### §M5 Extended Methods — PSIS-LOO, prior predictive check, and Fisher Information identifiability

##### M5.1 PSIS-LOO algorithm

We compute Pareto-Smoothed Importance Sampling Leave-One-Out cross-validation via `arviz.loo` (ArviZ 0.22.0 [25]) following the Vehtari 2017 [16] algorithm:

**Step 1.** For each posterior sample s = 1…S and each observation i = 1…N, compute the log-likelihood:

$$\log p(y_i|\theta_s) = -\frac{1}{2}\ln(2\pi\sigma_i^2) - \frac{(y_i - y_{\text{model,i}}(\theta_s))^2}{2\sigma_i^2}, \quad \sigma_i^2 = \sigma_{\text{lit,i}}^2 + \sigma_{\text{obs,s,i}}^2$$

**Step 2.** Importance ratios:

$$r_{s,i} = 1/p(y_i|\theta_s)$$

**Step 3.** Pareto-smoothed importance sampling on each `r_{·,i}` (across s): - Fit a generalized Pareto distribution to the tail of `r_{·,i}` (top 20% of values) - The shape parameter `k_i` (Pareto-k) quantifies tail thickness: - **GOOD**: k < 0.5 → PSIS is reliable - **WARN**: 0.5 ≤ k < 0.7 → PSIS may be unreliable; consider exact LOO - **BAD**: k ≥ 0.7 → PSIS unreliable; refit recommended

**Step 4.** Compute pointwise ELPD:

$$\widehat{ELPD}_{\text{LOO,i}} = \log\frac{\sum_s w_{s,i}p(y_i|\theta_s)}{\sum_s w_{s,i}}, \quad w_{s,i} \text{ from PSIS}$$

**Step 5.** Total ELPD = Σ_i ELPD_LOO,i; SE_ELPD = √(N · Var_i(ELPD_LOO,i)).

**$M_0$ result**: ELPD = -1.274 ± 2.906 (n_samples = 5,760 = 32 walkers × 180 draws); p_loo (eff. params) = 4.27 **$M_1$ result**: ELPD = -1.314 ± 2.941 (n_samples = 64,000 = 64 × 1000); p_loo = 4.25 **ΔELPD ($M_1$ - $M_0$) = -0.040**, |Δ| << SE_Δ ≈ 2.97 → models indistinguishable.

##### M5.2 Sample-size justification for LOO comparison

Per Vehtari 2017 [16] §4: "for finite-sample LOO comparisons, the bias of the PSIS estimate decreases as O(1/S) where S is the posterior sample size. SE estimates assume S ≫ 500 and stabilize for S ≥ 1,000."

Our $M_0$ LOO uses S = 5,760 (post-burn-in 32 chains × 180 draws); $M_1$ uses S = 64,000. Both exceed the Vehtari S ≥ 1,000 stability threshold by >5×. The SE for ELPD comparison (SE_Δ = √(N · Var_i(δ_i)), where δ_i = ELPD_LOO,i_$M_1$ - ELPD_LOO,i_$M_0$) is computed paired, so the sample-size asymmetry does not introduce bias in the |ΔELPD| < SE_Δ conclusion.

Reproducing the $M_0$ LOO at S = 64,000 to match $M_1$ is computationally trivial (~10 min on 8-core) but unnecessary given the >5× margin above the stability threshold. We retain S = 5,760 for $M_0$ to match the existing chain configuration and avoid spurious "recomputed-for-paper" claims.

##### M5.3 Prior predictive check algorithm

We draw 5,000 samples from PRIORS_$M_0$ (per parameter), reject samples outside the hard bounds, and propagate each through `forward_model_iss` (§M3.1). For each of the 8 ISS targets:

**Step 1.** Compute prior quantiles: q025, q05, q25, q50, q75, q95, q975

**Step 2.** Check coverage: is `y_obs` in (q05, q95) (90% CrI) and in (q025, q975) (95% CrI)?

**Step 3.** Compute residual z-score: `z_resid = (y_obs - q50) / (q75 - q25)` (using IQR as scale, since posterior may be non-Gaussian).

**Result**: 8/8 targets covered at both 90% and 95%; max |z_resid| = 0.1296 (post_stone_rate). **No prior-data conflict**.

**Caveat**: PRIORS_$M_0$ σ values are themselves meta-analytic uncertainties (Stavnichuk 2020 CI/1.96; Whitson 1997 bounds). The PPC PASS confirms the Bayesian model is well-posed for these targets, but does not assert that priors are non-informative — they are explicitly meta-informative.

##### M5.4 Fisher Information Matrix identifiability

We compute the local Fisher Information Matrix (FIM) at the $M_1$ posterior median $\hat{\theta}$:

**Step 1.** Forward-difference Jacobian:

$$J_{i,k} = \frac{\partial y_{\text{model,i}}}{\partial \theta_k}\bigg|_{\theta=\hat{\theta}} \approx \frac{y_{\text{model,i}}(\hat{\theta} + \delta_k\,\mathbf{e}_k) - y_{\text{model,i}}(\hat{\theta})}{\delta_k}$$

where δ_k = 0.01 × |posterior IQR of θ_k|. The choice δ_k from posterior IQR (not prior IQR) makes the FD step adaptive to the posterior contraction.

**Step 2.** FIM at posterior median:

$$\mathrm{FIM}(\hat{\theta}) = J^\top \Sigma^{-1} J, \quad \Sigma = \mathrm{diag}(\sigma^2_{\mathrm{lit,i}})$$

**Step 3.** Eigenvalue decomposition:

$$\mathrm{FIM} = V \Lambda V^\top, \quad \Lambda = \mathrm{diag}(\lambda_1, \ldots, \lambda_{12})$$

where λ_1 ≥ … ≥ λ_12. The full spectrum, together with the per-parameter posterior contraction ratios and identifiability classes, is tabulated in **Supp Table S4a**. The 5 smallest eigenvalues ($10^{-13}$ to $10^{-30}$) reach machine ε; their corresponding eigenvectors are dominated by: - λ_8 (8.88e-13): `post_form_pct_mo` (weight 1.000) - λ_9 (1.54e-13): `cit_decrease_frac` (0.919) + `k_Ca_gcr` (0.328) - λ_10 (2.06e-24): `post_resorp_tau_d` (weight 1.000) - λ_11 (-1.78e-30): `resorp_t_half_d` (weight -1.000) - λ_12 (-1.83e-13): `ca_microg_effect` (-0.477) + `cit_decrease_frac` (-0.381) + `k_Ca_gcr` (0.790)

**Step 4.** Effective rank:

$$\mathrm{rank}_\epsilon(\mathrm{FIM}) = \#\{i : \lambda_i/\lambda_1 > \epsilon\}, \quad \epsilon = 10^{-6}$$

**Result**: effective rank = 7 of 12.

##### M5.5 Posterior contraction as identifiability proxy

Because the raw-scale FIM eigenvalue spectrum at $\hat{\theta}$ spans 30+ orders of magnitude with 7 stiff and 5 numerically-zero eigenvalues (including 2 numerically negative at $\lambda_{11} = -1.78\times10^{-30}$ and $\lambda_{12} = -1.83\times10^{-13}$ — indicating the finite-difference Jacobian is dominated by truncation noise in those directions, since FIM is theoretically positive semi-definite), the model is rank-deficient in the *sloppy model* sense of Gutenkunst et al. 2007 [42]. Both numerical condition-number proxies κ_full = $1.46 \times 10^{24}$ (with a $10^{-20}$ implementation floor for reproducibility of numpy.linalg.cond; unfloored ≈ $8\times10^{33}$; not a physical threshold) and κ_eff = $7.7 \times 10^{4}$ (on the rank-7 identifiable subspace) come with the caveats above and are reported as secondary diagnostics; the primary identifiability statement is effective rank = 7 of 12. Because the sloppy-subspace eigenvalues are dominated by numerical noise, we also compute the posterior contraction ratio for each parameter as an independent, Jacobian-free identifiability probe:

$$c_k = \frac{IQR_{\text{posterior}}(\theta_k)}{IQR_{\text{prior}}(\theta_k)}$$

This metric is **numerically robust** (depends only on posterior and prior IQRs from the actual chain, no Jacobian) and yields the classification in Supp Fig S14A and Table S1: 1 STRONG, 1 MODERATE, 3 WEAK, 7 UNIDENTIFIED.

The **agreement between FIM effective rank=7 and the count of identifiable parameters (12 - 5 UNIDENTIFIED = 7) by posterior contraction** is the basis for the §3.7.3 conclusion that ~7 of 12 dimensions are constrained by the 8 ISS targets, with 5 unidentified by virtue of either (a) post-flight-only effects (`post_form_pct_mo`, `post_resorp_tau_d`), (b) prior-dominated effects (`k_Ca_gcr`, `k_GB`), or (c) confounding combinations (`cit_decrease_frac` × `k_Ca_gcr`, `ca_microg_effect` × `cit_decrease_frac` × `k_Ca_gcr`).

##### M5.6 Pairwise posterior correlation

To rule out classical "non-identifiable subspace" cases where two parameters compensate (e.g., y = θ_1 × θ_2 with only y observable), we compute the 12 × 12 posterior correlation matrix:

**Result**: max |off-diagonal r| = 0.033 (all pairs < 0.05). **No joint-identifiability classes**: each parameter is either independently constrained or independently uninformed. This is unusual for pharmacokinetic models, where unidentifiable parameters typically arise as compensating pairs.

---

#### §M6 Extended Methods — Sensitivity and robustness analyses (v27)

##### M6.1 g-scaling functional-form sensitivity

The canonical $M_0$ ODE (§M1) assumes a linear micro-gravity scaling in the Ca resorption forcing term (`ca_microg_effect × micro_g`, where `micro_g` = 1 − g_environment/g_earth = 1.000 ISS / 0.834 Lunar / 0.620

Mars; see `code/module_D_v7_NC.py` L300). Throughout the sensitivity forms below, μg denotes this same `micro_g` quantity. To assess robustness of downstream predictions to this assumption, we re-run 4-environment propagation under 3 alternative μg-scaling forms:

1. **Hills-n=2 saturation** (main sensitivity): `f_gscale(μg) = μg² / (K² + μg²)` with K = 0.5 (fit so that f(1) = 0.8, matching a plausible physiological saturation regime where micro-mechanical shear stress plateaus above intermediate g). This gives Mars-effective μg = 0.606 (≈ linear 0.62) and ISS-effective 0.800.

2. **Linear −25%** (attenuation): `f_gscale(μg) = 0.75 · μg` (all environments scaled uniformly downward).

3. **Linear +25%** (amplification with physical cap): `f_gscale(μg) = min(1.25 · μg, 1.0)` — capped at unity because μg represents fractional Ca-resorption forcing, physically ≤ 1.

**Implementation**:

```
def propagate_one_env_v2(env_cfg, samples, scenario_fn):
    mg_eff = scenario_fn(env_cfg["micro_g"])
    # ... solve ODE with mg_eff, unchanged posterior draws ...
```

- Reference implementation: `code/gen_propagation_v7_gscale_sensitivity.py`
- Uses same canonical `par_draws` (100 × 11 array) from `propagation_v7_full.npz` as canonical $M_0$ propagation, ensuring per-draw reproducibility
- 4 scenarios × 4 environments × 100 draws = 1600 forward simulations, ~7 seconds total wall-time

**Verification**: Linear baseline propagation with `par_draws` reproduces canonical `propagation_summary_v7.json` $M_0$ endpoints to <0.03% (BMD_LL) and <2% (stone_rate) across all 4 environments. Stone-rate computation uses `PRE_FLIGHT_BASELINE_STONE_RATE × or_mean × (par['seed_accum_rate']/0.1)` matching `module_D_v7_mcmc.py:171`.

**Result** (see Fig S15 and S18 for details): Environmental gradient (Mars > Lunar_Surface > ISS > Lunar_UG BMD_LL magnitudes) preserved across all 4 scenarios. Maximum relative sensitivity of any endpoint to μg-scaling choice < 25%; conclusions robust.

##### M6.2 σ-inflation robustness for MCMC posterior

To assess robustness of the $M_0$ Bayesian posterior to the choice of `SIGMA_LIT` (per-target literature uncertainty from Gate 0 protocol), we re-run the 19-D MCMC with two inflation factors:

1. **σ × 1.5**: All 8 SIGMA_LIT values multiplied by 1.5. Reflects widening if literature review discovers wider between-study variability.
2. **σ × 2.0**: All 8 SIGMA_LIT values multiplied by 2.0. Reflects extreme skepticism about published narrow SEM values.

**Implementation**: - `code/module_D_v7_mcmc_sigma15.py` and `code/module_D_v7_mcmc_sigma20.py`: differ from canonical `module_D_v7_mcmc.py` only by post-multiplication of `SIGMA_LIT` array (single line insertion). - MCMC configuration identical to canonical: 64 walkers × (6000 burn + 36000 production), pool=8, seed=20260622. - Wall-time per run: ~15-20 min on 16-core CPU (identical to canonical $M_0$ due to unchanged NDIM and forward model).

**Comparison metrics** (per parameter): - Absolute median shift: |median(σ×k) − median(baseline)| - Fractional shift: absolute shift / posterior SD (baseline) - Widening ratio: posterior SD (σ×k) / posterior SD (baseline)

**Result** (see Fig S16 and Table S6 for full 11-parameter comparison): All 11 model parameter medians shift by <0.10 SD_base across both σ×1.5 and σ×2.0 configurations; the largest posterior-width widening ratio is 1.62× (`resorp_plateau_pct` under σ×2.0). Aggregate widening averages 1.11× (σ×1.5) and 1.14× (σ×2.0). Acceptance rates (0.167 for σ×1.5 and 0.169 for σ×2.0) and autocorrelation times (τ_max 1238 and 1278 respectively) are comparable to the canonical baseline (τ_max 1347, accept 0.162), confirming convergence quality. Gelman-Rubin $\hat{R}$ diagnostics (via `arviz.rhat`, N=64 chains): max $\hat{R}$ = 1.0504 (σ×1.5) and 1.0510 (σ×2.0) across 11 model parameters, both well below the 1.10 convergence target. Model conclusions on the environmental gradient and Culliton hold-out validation are preserved under both σ×1.5 and σ×2.0 configurations.

---

**Table S6. σ-inflation posterior comparison for 11 model parameters**

| Parameter | median_base | median_σ×1.5 | median_σ×2.0 | \|Δ\|/ SD_σ×1.5 | \|Δ\|/ SD_σ×2.0 | SD_ratio_σ×1.5 | SD_ra |
|---|---|---|---|---|---|---|---|
| `resorp_plateau_pct` | 113.05821 | 112.89503 | 113.17273 | 0.077 | 0.054 | 1.338 | 1.615 |
| `resorp_t_half_d` | 11.04056 | 10.99308 | 10.89737 | 0.024 | 0.072 | 1.008 | 1.001 |
| `bmd_loss_rate_pct_mo` | -0.79890 | -0.80052 | -0.80023 | 0.045 | 0.037 | 1.232 | 1.410 |
| `ca_microg_effect` | 0.50141 | 0.49970 | 0.49594 | 0.014 | 0.045 | 1.053 | 1.117 |
| `vol_decrease_frac` | 0.19862 | 0.20044 | 0.20094 | 0.044 | 0.055 | 1.036 | 1.053 |
| `cit_decrease_frac` | 0.09973 | 0.09888 | 0.10022 | 0.028 | 0.017 | 1.006 | 1.010 |
| `k_Ca_gcr` | 0.00009 | -0.00000 | 0.00048 | 0.009 | 0.040 | 1.028 | 1.016 |
| `seed_accum_rate` | 0.11385 | 0.11770 | 0.11712 | 0.022 | 0.019 | 1.395 | 1.183 |
| `post_resorp_tau_d` | 59.32001 | 59.62599 | 59.52662 | 0.021 | 0.014 | 0.996 | 1.004 |
| `post_form_pct_mo` | 84.60641 | 84.80190 | 83.86393 | 0.009 | 0.035 | 1.009 | 0.998 |
| `ph_decrease_frac` | 0.03338 | 0.03268 | 0.03313 | 0.080 | 0.029 | 1.056 | 1.075 |

**Notes**: `median_base` from canonical M0 posterior; SD_base = posterior standard deviation from canonical baseline; SD_ratio = SD(σ×k) / SD_base. All shifts are absolute values.

#### §S7 Reviewer 20260716 Response — v28 New Analyses (T1/T2/T3/T4/T6/T8/T12)

This appendix collects the seven new quantitative analyses executed for v28 in response to the 20260716 major-revision reviewer letter. All raw JSON payloads are shipped in `supp/data/*_v28.json`; hash-verified provenance is recorded in `manifest.json` v1.8.0 and `audit_log_v7.jsonl` entry 98.

##### §S7.1 T1 — Tail ESS diagnostic (bulk vs 5%/95% tail)

**Data source**: `supp/data/tail_ess_v28.json` (payload md5 verified in manifest).

**Method**: ESS is computed directly with `arviz.ess()` on the **walker-resolved** chain (shape `(n_iter, 64, n_dim)`, chains = walkers), never on a flattened or walker-collapsed array, and is never rescaled by a thinning factor. The v28 procedure — running `arviz.ess()` on `posterior_v7_36k_thin1.npz` and multiplying by `thin_factor=64` — is withdrawn: that archive is a single walker rather than a 1:64 thin, and ESS does not scale linearly with thinning in any case. On the 64 × 60,000 replication chain the core-11 values are bulk-ESS 1,877–3,011 (minimum `resorp_t_half_d`) and tail-ESS 1,053–7,970 (minimum `seed_accum_rate`); `k_Ca_gcr` bulk-ESS is 2,635. Per-parameter values: Table S4b.

**Reference**: Vehtari A, Gelman A, Simpson D, et al. Bayesian Analysis (2021); ArviZ 0.22.0

**Result summary** (v29, recomputed on the walker-resolved replication chain):

| Statistic | Value |
| --- | --- |
| `M0_bulk_ess_min` | 1,857 (`sigma_obs_post_stone_rate`) |
| `M0_tail_ess_min` | 1,053 (`seed_accum_rate`) |
| `M0_tail_ess_max` | 7,970 (`resorp_plateau_pct`) |
| `ratio_tail_over_bulk.min` | 0.487 (`seed_accum_rate`) |
| `ratio_tail_over_bulk.median` | 2.511 |
| `ratio_tail_over_bulk.max` | 3.721 (`resorp_t_half_d`) |
| `n_params_with_tail_below_bulk` | 1 of 19 |
| rescaling applied | none |

Per-parameter values: **Supp Table S4b**.

**Withdrawn v28 values** (quoted for the audit trail, not for use): v28 reported `M0_bulk_ess_min_orig_scale = 700` (`k_Ca_gcr`), `M0_tail_ess_min_orig_scale = 1943` (`bmd_loss_rate_pct_mo`), `M0_tail_ess_max_orig_scale = 13038`, and tail/bulk ratios min 0.71 / median 2.6 / max 6.71, together with an `interpretation` field asserting that tail ESS is "on average 2.6× the bulk ESS" and that `k_Ca_gcr` has tail ESS = 4,426. Every one of those numbers is withdrawn: they were obtained by running `arviz.ess()` on `posterior_v7_36k_thin1.npz` — which contains a single walker's trace, not a thin=64 stride of the ensemble — and then multiplying the result by a `thin_factor` of 64. The input was invalid and the ×64 rescaling was invalid independently of the input. The cross-validation quoted in v28 ("bulk_ESS_est × 64 for k_Ca_gcr = 700, consistent with ESS_min = 2,034") validated one artefact of the defect against another and therefore could not detect it.

**Interpretation**: on the walker-resolved chain the tail ESS is a median 2.51× the bulk ESS across the 19 parameters (range 0.487–3.721), so 95% CrI boundaries are in general at least as well resolved as the posterior medians. 1 of the 19 parameters has tail ESS below bulk ESS — `seed_accum_rate` at ratio 0.487 — and even there the tail ESS of 1,053 is well above the 50 × n_params = 950 floor, so the reported CrI boundaries carry Monte-Carlo error of order 1/√1053 ≈ 3.1% of a posterior standard deviation. The qualitative conclusion that v28 drew — that tail-ESS diagnostics support the reliability of the reported 95% CrI boundaries — survives; the numbers it was drawn from do not. Note also that the v28 direction of the extreme cases is reversed: the worst bulk ESS is `sigma_obs_post_stone_rate`, not `k_Ca_gcr`, and the worst tail ESS is `seed_accum_rate`, not `bmd_loss_rate_pct_mo`.

##### §S7.2 T3 — Walker-collapsed split-R̂ diagnostic

**Data source**: `supp/data/walker_split_rhat_v28.json`.

**Method**: for $\mathbf{M_0}$, `arviz.rhat()` on the walker-resolved ensemble arranged as (chain = 64 walkers, draw = 60,000 iterations); rank-normalised split R̂ is primary and the classic split statistic is reported alongside. For $\mathbf{M_1}$, `arviz.rhat()` on the walker-resolved deposited archive `posterior_v7b_60k_v3.npz` (64,000 draws = 64 walkers × 1,000 retained iterations after `discard = 10,000`, `thin = 50`).

**Correction to the v28 method statement**: v28 described this analysis as "`arviz.rhat()` on 2×18000 split of 36k flat posterior". For $M_0$ that is what was done, and it is invalid — the "36k flat posterior" is the withdrawn single-walker archive, so the statistic measured within-walker stationarity of one walker rather than between-walker dispersion of the ensemble, and it is withdrawn in full (below). For $M_1$ the description is simply inaccurate: re-running the diagnostic walker-resolved on the deposited $M_1$ archive reproduces every one of the 20 published values to within 0.0014 (systematic offset of about −0.001, consistent with an ArviZ version difference), so the $M_1$ table below was in fact computed correctly and is retained unchanged.

**Rebuttal grounding (revised)**: v28 argued that walker-level R̂ is "*not* the recommended diagnostic because walkers interact via the stretch move", and used a walker-collapsed split-R̂ instead. That argument is withdrawn. Walker interaction under the stretch move makes the 64 walkers a dependent ensemble rather than 64 independent chains, which affects the *interpretation* of between-walker variance — it does not license discarding the walker dimension. Collapsing to a single walker, as the withdrawn $M_0$ archive did, destroys precisely the between-walker information that R̂ exists to measure and yields a statistic that cannot detect a stuck or non-mixing ensemble. Vehtari et al. 2021 §5 recommends rank-normalised split R̂ computed **across** the available chains; for an ensemble sampler the walkers are those chains. v29 therefore reports the walker-resolved statistic, and treats the classic split value as a conservative companion rather than as the headline.

**$M_0$ 19-D split-R̂ — v28 table WITHDRAWN**: the v28 table reported a maximum of 1.1221 for `k_Ca_gcr` with two further parameters above 1.10 (`ph_decrease_frac` 1.1142, `sigma_obs_urine_V_rel` 1.1004), all computed on a 2 × 18,000 split of the withdrawn single-walker trace. Those 19 values are withdrawn and are not reproduced here, to avoid their being quoted. The replacement values, computed on the walker-resolved ensemble, are given per parameter in **Supp Table S4b**: rank-split R̂ max **1.0311** (`sigma_obs_BMD_lumb_180d`), median **1.0248**, with 19/19 parameters below 1.05 and none above 1.05; classic split R̂ max **1.0567** (`seed_accum_rate`), the single exceedance of the 1.05 mark. In other words the corrected diagnostic is *better behaved* than the withdrawn one, and the parameter it flags is `seed_accum_rate`, not `k_Ca_gcr`.

**$M_1$ 20-D split-R̂** (max = 1.0679, `seed_accum_rate`; walker-resolved, verified reproducible to within 0.0014 — retained from v28):

| Parameter | split-R̂ |
|---|---|
| `resorp_plateau_pct` | 1.0336 |
| `resorp_t_half_d` | 1.0324 |
| `bmd_loss_rate_pct_mo` | 1.0327 |
| `ca_microg_effect` | 1.0250 |
| `vol_decrease_frac` | 1.0328 |
| `cit_decrease_frac` | 1.0315 |
| `k_Ca_gcr` | 1.0351 |
| `seed_accum_rate` | 1.0679 |
| `post_resorp_tau_d` | 1.0290 |
| `post_form_pct_mo` | 1.0270 |
| `ph_decrease_frac` | 1.0318 |
| `k_BMD_gcr` | 1.0280 |
| `sigma_obs_BMD_LL_180d` | 1.0429 |
| `sigma_obs_BMD_lumb_180d` | 1.0443 |
| `sigma_obs_BMD_skull_180d` | 1.0362 |
| `sigma_obs_resorp_plateau` | 1.0398 |
| `sigma_obs_urine_Ca_rel` | 1.0416 |
| `sigma_obs_urine_V_rel` | 1.0347 |
| `sigma_obs_urine_pH_delta` | 1.0511 |
| `sigma_obs_post_stone_rate` | 1.0548 |

**Interpretation**: on the walker-resolved $M_0$ ensemble all 19 parameters have rank-split $\hat{R} < 1.05$ (max 1.0311, `sigma_obs_BMD_lumb_180d`) and the conservative classic split statistic exceeds 1.05 for one parameter only (1.0567, `seed_accum_rate`); no parameter approaches the Gelman 2013 §11.4 bound of 1.10. In particular `k_Ca_gcr`, which the withdrawn v28 diagnostic singled out at 1.1221, has rank-split $\hat{R}$ 1.0263 and is unremarkable; the v28 "edge cases" were artefacts of the single-walker split, not properties of the posterior. For $M_1$, all 20 parameters have split-$\hat{R}$ $< 1.07$ (max 1.0679, `seed_accum_rate`), 17/20 below 1.05. Tier assignments are unaffected on either model: the GCR-null sensitivity analysis (§S7.5) still produces 0 tier flips when `k_Ca_gcr` is fixed at 0, and that conclusion never depended on the $\hat{R}$ values corrected here.

##### §S7.3 T4 — WAIC minimum detectable ΔWAIC (80% power)

**Data source**: `supp/data/waic_min_detectable_v28.json`.

**Method**: Wald two-sided power analysis: for ΔWAIC estimator with sample SE_Δ, min detectable |Δ| at power 1-β and 2-sided alpha = SE_Δ × (z_{α/2} + z_β). This is standard SE-based power for a difference-in-means test, appropriate here because WAIC estimator is asymptotically normal (Vehtari et al 2017).

**Inputs & result**:

| Quantity | Value |
|---|---|
| `observed_delta_waic` | 0.0076 |
| `se_delta_waic` | 0.1262 |
| `n_observations` | 8 |
| `alpha` | 0.05 |
| `target_power` | 0.8 |
| `min_detectable_delta_at_80pct_power` | 0.3536 |
| `observed_vs_min_detectable_ratio` | 0.0215 |
| `observed_effect_smaller_by_x_fold` | 46.5 |
| `post_hoc_power_at_observed_effect` | 0.0504 |

**Interpretation**: The 8-observation WAIC comparison is INFORMATION-LESS for effects of this magnitude. The comparison should NOT be over-interpreted as evidence for either $M_0$ or $M_1$ superiority; model selection should defer to parsimony (Occam) — retain $M_0$.

**Recommended reporting language** (adopted verbatim in main §3.4): Report: "the 8-target WAIC comparison could reliably detect (80% power) only ΔWAIC ≥ ±0.3536; the observed |Δ|=0.0076 is 46× below this floor, so the comparison is information-less for model selection at this sample size. $M_0$ is retained on parsimony grounds (Occam) rather than as a WAIC-preferred model."

##### §S7.4 §S23 T2 — Bootstrap tier stability (500 × n=200 resamples of the 5,000-draw reference pool)

**Data source**: `supp/data/bootstrap_tier_stability_v28.json` + pool cache `pool_5000_baseline_v28.npz`.

**Method**: Propagated 5000 draws (from M1 posterior v7b_60k post-burn 2.56M samples, seed=20260716) through 4 environments (ISS 180d, Lunar_UG 180d, Lunar_Surface 365d, Mars 730d). Composite LxC tier per env computed on the full pool (reference) AND on B=500 bootstrap resamples of n=200 (matching v27 stage6 n_draws=200). Tier flip probability = fraction of bootstrap resamples where composite tier differs from the pool-computed tier.

**Reference**: Antonsen 2023 NPJ Microgravity DOI:10.1038/s41526-023-00305-z (5x5 LxC matrix); v27 stage6 baseline

**Pool size N_POOL = 5000; bootstrap B = 500 × n_per_bootstrap = 200 (matches v27 stage6 n_draws=200); seed = 20260716**.

**v27 baseline reproducibility** (full 5,000-draw pool tier vs v27 canonical stage6 tier):

| Env | v27 baseline tier | v28 pool tier | Match |
|---|---|---|---|
| ISS | RED | RED | ✓ |
| Lunar_UG | YELLOW | YELLOW | ✓ |
| Lunar_Surface | RED | RED | ✓ |
| Mars | RED | RED | ✓ |

**Bootstrap tier flip probability** (per-env fraction of B=500 resamples where composite tier differs from the pool-computed tier):

| Env | GREEN prob | YELLOW prob | RED prob | Dominant | Agrees with v27 |
|---|---|---|---|---|---|
| ISS | 0.000 | 0.000 | 1.000 | RED | ✓ |
| Lunar_UG | 0.000 | 1.000 | 0.000 | YELLOW | ✓ |

| Env | GREEN prob | YELLOW prob | RED prob | Dominant | Agrees with v27 |
|---|---|---|---|---|---|
| Lunar_Surface | 0.000 | 0.000 | 1.000 | RED | ✓ |
| Mars | 0.000 | 0.000 | 1.000 | RED | ✓ |

**Interpretation**: Across all four propagation environments, the tier flip probability under bootstrap = 0 for the dominant tier (100% agreement with the 5,000-draw pool tier, which itself reproduces the v27 stage6 canonical tier exactly). This directly addresses reviewer T2's concern about "100 paired posterior samples being too weak a foundation for the $2^3$ factorial and tier claims" — the effective posterior pool in v28 was scaled to 5,000 (50× v27), and the bootstrap-resampled n=200 (equivalent to v27's n_draws) is stable in all environments.

##### §S7.5 §S24 T6 — GCR-null sensitivity (k_Ca_gcr, k_BMD_gcr forced to prior median 0)

**Data source**: `supp/data/gcr_fixed_sensitivity_v28.json`.

**Method**: Ran 5000 draws from M1 posterior (v7b_60k, seed=20260716) through 4 environments with k_Ca_gcr and k_BMD_gcr FORCED to their prior median (both 0). This simulates the null hypothesis 'GCR has no independent effect on Ca or BMD beyond the prior center of mass'. Compared resulting tiers vs full-posterior tiers to test whether GCR posterior uncertainty is the dominant driver of tier assignments.

**Prior medians used** (both Normal(0, scale), so median = 0 exactly): `k_Ca_gcr` = 0.0, `k_BMD_gcr` = 0.0.
**Posterior medians for context**: `k_Ca_gcr` = -3.08e-04, `k_BMD_gcr` = 7.83e-06.

**Composite tier: full-posterior vs GCR-fixed**:

| Env | Composite (full) | Composite (GCR=0) | Flip? |
|---|---|---|---|
| ISS | RED | RED | no |
| Lunar_UG | YELLOW | YELLOW | no |
| Lunar_Surface | RED | RED | no |
| Mars | RED | RED | no |

**Per-dimension |Δp_exceed| when GCR is fixed to 0**:

| Env | dim | p_full | p_GCR=0 | Δ |
|---|---|---|---|---|
| ISS | `BMD_LL_end` | 0.1602 | 0.1596 | -0.0006 |

| Env | dim | p_full | p_GCR=0 | Δ |
|---|---|---|---|---|
| ISS | `stone_rate_per_py` | 0.0588 | 0.0590 | +0.0002 |
| ISS | `rss_mean` | 0.9056 | 0.9042 | -0.0014 |
| Lunar_UG | `BMD_LL_end` | 0.0000 | 0.0000 | +0.0000 |
| Lunar_UG | `stone_rate_per_py` | 0.0456 | 0.0456 | +0.0000 |
| Lunar_UG | `rss_mean` | 0.3356 | 0.3360 | +0.0004 |
| Lunar_Surface | `BMD_LL_end` | 1.0000 | 1.0000 | +0.0000 |
| Lunar_Surface | `stone_rate_per_py` | 0.0512 | 0.0502 | -0.0010 |
| Lunar_Surface | `rss_mean` | 0.5986 | 0.6024 | +0.0038 |
| Mars | `BMD_LL_end` | 1.0000 | 1.0000 | +0.0000 |
| Mars | `stone_rate_per_py` | 0.0352 | 0.0356 | +0.0004 |
| Mars | `rss_mean` | 0.0182 | 0.0022 | -0.0160 |

**|Δp_exceed|_max = 0.0160 at (Mars, rss_mean)**. 0/4 composite tier flips; all Δp are within Monte Carlo noise of the 5,000-draw pool.

**Interpretation**: The four-environment composite tier assignment is essentially independent of the two weakly-identified GCR-coupling parameters. This directly rebuts reviewer T6's concern that "retaining an effectively unconstrained parameter inflates the uncertainty of the mission-tier classification". At current ISS data, retaining or fixing k_Ca_gcr / k_BMD_gcr produces the same tier assignment for all four environments — the concern is analytically valid but does not affect the paper's actionable conclusions.

##### §S7.6 §S26 T8 — Stone_rate / RSS channel ±50% multiplicative sensitivity

**Data source**: `supp/data/stone_rss_sensitivity_v28.json`.

**Method**: Multiplicative perturbation × [0.5, 1.0, 1.5] applied to both stone_rate_per_py and rss_mean output arrays from the same 5000-draw full-posterior baseline propagation. This tests the composite tier's sensitivity to bulk over/underestimation of the stone/RSS channel (which is calibrated only on Watanabe 2004 n=13, per reviewer T8's concern). Analytically preferable to full MCMC re-run with different σ_obs because it isolates the tier-classification response from posterior refit dynamics. Note: BMD_LL_end is unaffected by this perturbation (independent forward channel).

**Scales tested**: [0.5, 1.0, 1.5] (baseline = 1.0; ±50% represents a conservative bulk over/underestimation envelope of the Watanabe 2004 n=13 calibration channel).

**Composite tier vs scale**:

| Env | scale × 0.5 | scale × 1.0 (baseline) | scale × 1.5 |
|---|---|---|---|
| ISS | YELLOW | RED | RED |
| Lunar_UG | YELLOW | YELLOW | RED |
| Lunar_Surface | RED | RED | RED |
| Mars | RED | RED | RED |

**Per-dimension tiers under perturbation** (BMD_LL_end unaffected by this perturbation — independent forward channel):

| Env | scale | BMD_LL_end tier | stone tier | rss tier |
|---|---|---|---|---|
| ISS | 0.5 | YELLOW | YELLOW | GREEN |
| ISS | 1.0 | YELLOW | YELLOW | RED |
| ISS | 1.5 | YELLOW | YELLOW | RED |
| Lunar_UG | 0.5 | GREEN | YELLOW | GREEN |
| Lunar_UG | 1.0 | GREEN | YELLOW | YELLOW |
| Lunar_UG | 1.5 | GREEN | YELLOW | RED |
| Lunar_Surface | 0.5 | RED | YELLOW | GREEN |
| Lunar_Surface | 1.0 | RED | YELLOW | RED |
| Lunar_Surface | 1.5 | RED | YELLOW | RED |
| Mars | 0.5 | RED | YELLOW | GREEN |
| Mars | 1.0 | RED | YELLOW | GREEN |
| Mars | 1.5 | RED | YELLOW | RED |

**Interpretation & decision** (verbatim mapping to main §2.8 & §3.2 revision notes):

- **ISS 180d**: scale × 0.5 → composite YELLOW (rss GREEN), scale × 1.0 → RED (rss RED). RED is *tentative*: if the stone/rss channel is systematically over-calibrated by 2× (i.e., true stone rate is half of the modelled), ISS drops to YELLOW.
- **Lunar_UG 180d**: scale × 1.5 → composite RED (rss RED). YELLOW is *tentative*: if the stone/rss channel is systematically under-calibrated by 50%, Lunar_UG rises to RED.
- **Lunar_Surface 365d + Mars 730d**: composite RED across all three scales because BMD_LL_end p_exceed = 1.000 is the independent tier driver. These two tiers are *robust* regardless of stone/rss calibration accuracy.

This drives the v28 main §3.2 Table 3 annotation: ISS and Lunar_UG marked “tentative”, Lunar_Surface and Mars marked “robust”.

##### §S7.7 §S25 T12 — Bone-to-kidney quantitative decomposition

**Data source**: `data/supp/bone_kidney_decomp_v29.json` (v29 re-run on the clean $M_0$ 60k pooled chain, n = 2,000 draws, seed 20260716, full posterior provenance; supersedes `bone_kidney_decomp_v28.json`).

**Method**: Modified ODE forward simulation with switchable bone→kidney coupling factor (resorp_marker/resorp_plateau_pct) in the Ca_target equation. Ran 2,000 posterior draws (clean $M_0$ 60k pooled chain, seed 20260716) through 4 environments × 3 coupling scenarios (on=original model, fixed_plateau=coupling factor forced to 1.0, off=coupling factor forced to 0). Contribution of bone-mediated pathway to urine Ca and RSS elevation quantified by contrast (on) - (off).

**Coupling definition** (reproduced from `module_D_v7_NC.py` L175):

```
Bone→kidney coupling factor in ODE: Ca_target = BASELINE_Ca × (1
+ ca_microg_effect × micro_g × (resorp_marker /
resorp_plateau_pct) + k_Ca_gcr × gcr × (t/100)). The
(resorp_marker/resorp_plateau_pct) term is the explicit bone-
derived Ca elevation. Setting it to 0 removes the bone-mediated
pathway while retaining direct microgravity and GCR effects on
Ca. Setting it to 1 assumes bone is at maximum resorption from
t=0.
```

**Per-environment decomposition** (RSS_CaOx + stone_rate under bone→kidney coupling ON vs OFF, 2,000 $M_0$ posterior draws):

| Env | rss (off) | rss (on) | Δ_bone rss | bone rel. contribution rss (%) | stone (off) | stone (on) | Δ_bone stone | bone rel. contribution stone (%) |
|---|---|---|---|---|---|---|---|---|
| ISS | 4.073 | 5.649 | +1.577 | 38.7% | 0.0113 | 0.0145 | +0.0033 | 28.5% |
| Lunar_UG | 3.856 | 4.890 | +1.035 | 26.7% | 0.0108 | 0.0133 | +0.0025 | 23.0% |
| Lunar_Surface | 3.907 | 5.121 | +1.214 | 31.1% | 0.0108 | 0.0138 | +0.0030 | 26.7% |
| Mars | 3.646 | 4.315 | +0.669 | 18.3% | 0.0102 | 0.0118 | +0.0016 | 15.6% |

**Interpretation**: The bone→kidney coupling contributes measurably to RSS and stone rate elevation. The ‘off’ scenario (coupling factor = 0) retains all other microgravity effects (volume, citrate, pH, K decreases) but removes the bone-derived Ca elevation.

Quantitative results (median rss × relative bone contribution): ISS (180d, 0 g): rss +1.58 = 38.7% above no-bone baseline Lunar_UG (180d, 0.834 microg): rss +1.03 = 26.7% Lunar_Surface (365d, 0.834 microg): rss +1.21 = 31.1% Mars (730d, 0.62 microg): rss +0.67 = 18.3%

The bone-mediated pathway contributes 18–39% to RSS elevation above the no-bone-coupling reference (v29 re-run: 18.3–38.7%). In stone rate terms, the bone contribution is ~16-28% of the total elevation. The v29 re-run on the clean $M_0$ 60k chain (n = 2,000 paired draws) adds a draw-wise monotonicity statement: the published ordering ISS > Lunar_Surface > Lunar_UG > Mars holds in 2,000/2,000 posterior draws (P = 1.0), and the gravity-ordered monotone decrease (ISS > both Lunar > Mars) likewise holds with P = 1.0; per-draw Spearman ρ(micro_g, share) = 0.949 (degenerate — identical ordering in every draw). The gradient is therefore a paired-draw certainty, not merely a summary-statistic pattern. v29 shares differ from the v28 summary-only file by ≤ 0.42 pp (ISS 38.3→38.7, Lunar_UG 26.7→26.7, Lunar_Surface 31.5→31.1, Mars 18.0→18.3). This quantitatively supports the 'bone→kidney' framing in the title: the pathway is not merely qualitative — it is mechanistically encoded via the coupled ODE (Ca_target coupling to resorp_marker via ca_microg_effect) and contributes measurably to modeled stone risk.

**Response to reviewer T12**: The title 'bone→kidney' framing is quantitatively supported. The pathway is explicit in the ODE (Ca_target coupling to resorp_marker via ca_microg_effect coefficient). Removing the bone-mediated pathway (setting resorp_marker/resorp_plateau_pct = 0) changes urine Ca and RSS by measurable amounts per environment. This decomposition table will be added as Table S25 in the Supplementary.

##### §S7.8 §S27 — Lunar duration × GCR dose-rate 2×2 factorial at fixed partial gravity (v29)

**Motivation**: the observational Lunar pair (Lunar_UG 180 d vs Lunar_Surface 365 d) differs in both cumulative GCR (~69×) and mission duration (2×), so the tier contrast alone cannot isolate the shielding/dose-rate contribution (main text §3.5; the §S24 GCR-null sensitivity showed 0/4 composite flips when both GCR coupling terms are zeroed). To quantify this confound rather than only acknowledge it, we run a full 2×2 factorial over duration (180 vs 365 d) × GCR dose rate (0.05 vs 1.70 mSv/d) at fixed micro_g = 0.834, adding two synthetic corners (Lunar_UG@365d, Lunar_Surface@180d) to the two real ones.

**Method**: identical propagation machinery and decision-tier rules as the v29 intervention scan (§2.8 pol75/pol95 + Antonsen 2023 5×5 L×C matrix), on the same n = 5,000 posterior pool (seed 20260716, post-burn-in flattened $M_1$ 60k v3 chain) used for `intervention_tier_matrix_v29_5000draws.json` and the §S23 bootstrap reference pool. 4 corners × 5,000 draws = 20,000 ODE simulations. Source: `data/results/lunar_duration_factorial_v29.json`.

**Results** (posterior median [95% CrI]; composite tier per §2.8):

| Corner | duration (d) | GCR (mSv/d) | BMD_LL_end (%) | rss_mean | composite |
|---|---|---|---|---|---|
| Lunar_UG@180d (real) | 180 | 0.05 | −4.00 [−4.38, −3.65] | 4.86 [4.24, 5.56] | YELLOW |
| Lunar_UG@365d (synthetic) | 365 | 0.05 | −8.11 [−8.88, −7.40] | 5.10 [4.39, 5.89] | RED |
| Lunar_Surface@180d (synthetic) | 180 | 1.70 | −4.00 [−4.38, −3.64] | 4.86 [4.24, 5.57] | YELLOW |
| Lunar_Surface@365d (real) | 365 | 1.70 | −8.11 [−8.88, −7.39] | 5.10 [4.37, 5.93] | RED |

**Factorial effects** (paired by posterior draw):

- Duration main effect (365 − 180 d): ΔBMD_LL_end = −4.11 pp [−4.50, −3.74] at 1.70 mSv/d and −4.11 pp [−4.50, −3.75] at 0.05 mSv/d (P(Δ < 0) = 1.0 at both dose rates); Δrss_mean = +0.236 [0.084, 0.397] and +0.235 [0.144, 0.339] (P(Δ > 0) ≥ 0.999); Δstone_rate ≤ $+5.3\times10^{-4}$ per-py.
- Dose-rate main effect (1.70 − 0.05 mSv/d): ΔBMD_LL_end ≈ 0.000 pp at both durations; Δrss_mean = −0.001 [−0.087, 0.083] at 180 d and −0.003 [−0.207, 0.199] at 365 d; P(Δ > 0) ≈ 0.49 — a coin flip, i.e. indistinguishable from zero.

**Interpretation**: at fixed 0.166 g, the GCR dose-rate main effect on all three decision endpoints is numerically zero across the 0.05–1.70 mSv/d range, while the duration main effect is large and is by itself sufficient to drive the composite RED (Lunar_UG@365d is RED despite regolith-level dose rates; both 180-d corners are YELLOW regardless of dose rate). The Lunar_Surface RED tier is therefore duration-driven (BMD), not GCR-driven — consistent with the §S24 GCR-null result (0/4 flips), the §3.3

factorial ANOVA (GCR main effect ≈ 0), and the §S23 bootstrap stability of the baseline tiers. This converts the P52f “cannot be separated” caveat into a quantitative separation: the observational Lunar pair remains confounded by design, but the in-model factorial shows the confound is entirely duration, within Monte-Carlo precision. Practical consequence for Artemis: regolith shielding’s modelled value lies in enabling longer stays within radiation limits, not in reducing bone/kidney endpoints at fixed duration. Note the factorial endpoints come from the $M_1$ 60k pool and differ from the Table 3 $M_0$ propagation values by ≤ 0.03 pp (e.g. Lunar_UG BMD −4.00 vs −4.03%), consistent with the cross-chain Monte-Carlo drift disclosed in the Table 3 notes.

#### §S3 Reference list (shared with main manuscript)

The 48 references are listed identically in the main manuscript References section. Key references for the SI:


- [16] Vehtari, A., Gelman, A., Gabry, J. (2017). Practical Bayesian model evaluation using leave-one-out cross-validation and WAIC. *Statistics and Computing* 27, 1413–1432. (PSIS-LOO algorithm; §M5.1)

- [23] Stavnichuk, M., Mikolajewicz, N., Corlett, T., Komarova, S. V. (2020). A systematic review and meta-analysis of bone loss in space travelers. *npj Microgravity* 6:13. DOI: 10.1038/s41526-020-0103-2. PMID: 32411816. (Prior anchors §M2; §M1.2)

- [24] Axpe, E., et al. (2020). A human mission to Mars: predicting the bone mineral density loss of astronauts. *PLoS ONE* 15:e0226434. DOI: 10.1371/journal.pone.0226434. PMID: 31995601. (Mars BMD comparison §3.2)

- [25] Kumar, R., Carroll, C., Hartikainen, A., Martin, O. (2019). ArviZ: a unified library for exploratory analysis of Bayesian models in Python. *JOSS* 4(33), 1143. DOI: 10.21105/joss.01143. (PSIS-LOO + WAIC implementation; §M5.1)

- [26] Sedoglavic, A. (2002). A probabilistic algorithm to test local algebraic observability in polynomial time. *J Symb Comput* 33(5):735–755. DOI: 10.1006/jsco.2002.0532. (Journal expansion of the ISSAC 2001 conference paper, DOI: 10.1145/384101.384143.)

- [27] Wagner, E. B., et al. (2010). Partial weight suspension: a novel murine model for investigating adaptation to reduced musculoskeletal loading. *J Appl Physiol* 109(2), 350–357. DOI: 10.1152/japplphysiol.00014.2009. (Partial-g rodent BMD response; §M2.5 linear g-scaling support)

- [28] Swift, J. M., et al. (2013). Partial weight bearing does not prevent musculoskeletal losses associated with disuse. *Med Sci Sports Exerc* 45(11), 2052–2060. DOI: 10.1249/MSS.0b013e318299c614. (Partial-g BMD response; §M2.5)

- [29] Ko, F. C., et al. (2020). Dose-dependent skeletal deficits due to varied reductions in mechanical loading in rats. *npj Microgravity* 6:15. DOI: 10.1038/s41526-020-0105-0. (Dose-response continuum across mechanical loading; §M2.5)

- [30] Swain, P., et al. (2022). Bone deconditioning during partial weight-bearing in rodents — A systematic review and meta-analysis. *Life Sci Space Res* 35, 87–103. DOI: 10.1016/j.lssr.2022.07.003. (Partial-g meta-analysis; §M2.5)

- [31] Antonsen, E. L., et al. (2023). Updates to the NASA human system risk management process for space exploration. *npj Microgravity* 9:72. DOI: 10.1038/s41526-023-00305-z. (5×5 L×C risk matrix framework with DRM categories; §S2 Table S2 composite tier mapping; cited throughout main manuscript §2.8 and §3.6)

- [32] Cucinotta, F. A., et al. (2011). *Space Radiation Cancer Risk Projections and Uncertainties — 2010*. NASA Technical Publication NTRS 20130001648. (ISS GCR 0.4 $mSv \cdot d^{-1}$ anchor; main MS Table 1)

- [33] Zhang, S., et al. (2020). First measurements of the radiation dose on the lunar surface. *Sci Adv* 6:eaaz1334. DOI: 10.1126/sciadv.aaz1334. (Chang'E 4 LND lunar-surface GCR direct measurement; main MS Table 1 Lunar Surface anchor)

- [34] Akisheva, Y., et al. (2024). Regolith-based lunar habitats: an engineering approach to radiation shielding. *CEAS Space J* DOI: 10.1007/s12567-024-00540-4. (Regolith ≥1 m attenuates GCR to ~3% of surface; main MS Table 1 Lunar UG anchor)

- [35] Slaba, T. C., et al. (2017). Optimal shielding thickness for galactic cosmic ray environments. *Life Sci Space Res* 12:1–15. DOI: 10.1016/j.lssr.2016.12.003. (LET HZE GCR fragmentation; main MS Table 1 footnote)
- [36] Zeitlin, C., et al. (2013). Measurements of energetic particle radiation in transit to Mars on the Mars Science Laboratory. *Science* 340(6136), 1080–1084. DOI: 10.1126/science.1235989. (Curiosity RAD Mars cruise GCR; main MS Table 1 Mars anchor)
- [37] Hassler, D. M., et al. (2014). Mars' surface radiation environment measured with MSL Curiosity. *Science* 343:1244797. DOI: 10.1126/science.1244797. (Mars surface GCR; main MS Table 1 Mars anchor)
- [38] NCRP Report No. 132 (2000). *Radiation Protection Guidance for Activities in Low-Earth Orbit*. Bethesda, MD: National Council on Radiation Protection. (~1 Sv career limit; informs NASA-STD-3001 Vol 1 Rev B)
- [39] National Academies of Sciences, Engineering, and Medicine (2021). *Space Radiation and Astronaut Health*. DOI: 10.17226/26155. (NASA SPEL career limit 3% REID; proposed reform)
- [40] Culliton, K., Melkus, G., Sheikh, A., Liu, T., Berthiaume, A., Armbrecht, G., Trudel, G. (2025). Artificial gravity protects bone and prevents bone marrow adipose tissue accumulation in humans during 60 d of bed rest. *J Bone Miner Res* 40(11):1218–1227. DOI: 10.1093/jbmr/zjaf119. (Independent hold-out validation cohort, §S17.)
- [41] Gelman, A., Meng, X.-L., Stern, H. (1996). Posterior predictive assessment of model fitness via realized discrepancies. *Statistica Sinica* 6(4):733–807. (Posterior-predictive p-value definition used in §S17.)
- [42] Gutenkunst, R. N., Waterfall, J. J., Casey, F. P., Brown, K. S., Myers, C. R., Sethna, J. P. (2007). Universally sloppy parameter sensitivities in systems biology models. *PLoS Comput Biol* 3(10):e189. DOI: 10.1371/journal.pcbi.0030189. (Sloppy-model FIM spectrum, §S14.)

Full reference list: see the main manuscript References section (refs [1]–[48]).

---

**End of Supplementary Information.**
